# Microbial biomarkers of tuberculosis infection and disease in blood: systematic review and meta-analysis

**DOI:** 10.64898/2026.03.09.26347934

**Authors:** Shruthi Chandran, Edward Cruz Cervera, David A. Jolliffe, Divya Tiwari, David A. Barr, Graeme Meintjes, Rishi K. Gupta, Donald G. Catanzaro, Timothy C. Rodwell, Adrian R. Martineau

## Abstract

**Background:** Numerous studies reporting utility of microbial blood biomarkers for diagnosis and treatment monitoring of *M. tuberculosis* (Mtb) infection and tuberculosis disease have been conducted, but up-to-date systematic reviews and meta-analyses of these data are lacking. We aimed to evaluate diagnostic accuracy of microbial blood biomarkers for detection of tuberculosis disease and to characterise their responses to antimicrobial therapy.

**Methods:** For this aggregate data meta-analysis, we searched MEDLINE, EMBASE and Scopus from 1st January, 1990, to 22 October, 2025, using terms for “tuberculosis”, “microbial biomarker”, and “human blood” to identify studies in which participants underwent blood sampling for detection of cell-free Mtb DNA, cell-associated Mtb DNA, protein/peptide Mtb antigens and lipid/glycolipid Mtb antigens before ± after initiation of antimicrobial therapy. For bivariate analyses we used hierarchical summary receiver operating characteristic (HSROC) models to calculate areas under HSROC curves (AUC) for each analyte class to evaluate diagnostic accuracy for tuberculosis disease. For longitudinal analyses, we calculated risk differences to evaluate changes in proportions of biomarker-positive individuals after vs. before initiation of antimicrobial therapy, and pooled them using random-effects meta-analysis.

**Findings:** 137 eligible articles were identified in the search, of which 109 vs. 13 contributed data to bivariate vs. longitudinal analyses, respectively. For cell-free Mtb DNA targets, AUC was 0.87 (95% CI 0.84 to 0.89), with sensitivity 61.5% (51.0 to 71.0) and specificity 93.0% (88.1 to 96.1); 4,878 samples from 34 unique study/setting/biomarker combinations (investigations). For cell-associated Mtb DNA targets, AUC was 0.93 (0.90 to 0.95), with sensitivity 43.9% (29.4 to 59.4) and specificity 97.1% (94.5 to 98.5); 3,589 samples, 32 investigations. For protein/peptide targets, AUC was 0.94 (0.92 to 0.96), with sensitivity 78.9% (73.2 to 83.6) and specificity 92.9% (90.7 to 94.5%); 10,260 samples, 61 investigations. For lipid/glycolipid targets, AUC was 0.96 (0.94 to 0.97), with sensitivity 68.6% (54.1 to 80.3) and specificity 97.0% (94.0 to 98.5); 3,287 samples, 22 investigations. Pooled risk differences for proportions of individuals biomarker-positive after vs. before initiation of antimicrobial therapy were -0.44 (-0.89 to 0.01; 68 samples, 5 investigations) for cell-free Mtb DNA; -0.46 (-0.88 to -0.03; 89 samples, 5 investigations) for cell-associated Mtb DNA, and -0.24 (-0.75 to 0.28; 160 samples, 4 investigations) for protein/peptide antigens. No studies investigating responses of lipid/glycolipid antigens to antimicrobial therapy were identified. Heterogeneity was moderate (I^2^ 25-50%) for the majority of studies. 98/109 and 11/13 studies contributing data to bivariate vs. longitudinal analyses, respectively, were assessed as being at high risk of bias.

**Interpretation:** Molecular and biochemical microbial blood biomarkers exhibit similar accuracy for detection of tuberculosis disease, with specificity consistently exceeding sensitivity. Cell-associated Mtb DNA biomarkers exhibited a statistically significant response to antimicrobial therapy, with similar trends observed for cell-free Mtb DNA and protein/peptide antigens. These findings should be interpreted cautiously in the light of high risk of bias for many of the primary studies contributing data. Higher quality studies are needed to evaluate this emerging class of tuberculosis biomarkers.

**Funding:** Barts Charity, The Medical College of Saint Bartholomew’s Hospital Trust, Wellcome HARP Doctoral Fellowship Scheme.

**RESEARCH IN CONTEXT:** 

**Evidence before this study:** The World Health Organisation (WHO) has identified high-priority biomarker needs for screening, diagnosis, evaluating likelihood of disease progression and treatment monitoring for tuberculosis. Numerous studies reporting utility of microbial blood biomarkers for diagnosis and treatment monitoring of *M. tuberculosis* (Mtb) infection and tuberculosis disease have been conducted, but up-to-date systematic reviews and meta-analyses of these data are lacking. We performed a systematic search of MEDLINE, EMBASE and Scopus from 1st January, 1990, to 22 October, 2025, using terms for “tuberculosis”, “microbial biomarker”, and “human blood” to identify studies in which participants underwent blood sampling for detection of cell-free Mtb DNA, cell-associated Mtb DNA, protein/peptide Mtb antigens and lipid/glycolipid Mtb antigens before ± after initiation of antimicrobial therapy. Multiple studies have investigated utility of microbial blood biomarkers for detection of tuberculosis disease and monitoring responses to antimicrobial therapy, but only three relevant systematic reviews have been conducted to date, of which two (2007, 2021) report on detection of cell-free Mtb DNA, and one (2011) reports on antigen detection tests. Numerous primary studies have been published since these meta-analyses were conducted, but up-to-date syntheses incorporating the latest data for all classes of microbial blood biomarker for tuberculosis are lacking.

**Added value of this study:** To our knowledge, this study is the most comprehensive systematic review and meta-analysis of data from studies of microbial blood biomarkers of Mtb infection and tuberculosis disease conducted to date. It is also the first meta-analysis to synthesise data from studies investigating detection of cell-associated Mtb DNA in blood. Bivariate analysis of data from 109 studies revealed AUC values of 0.87 to 0.96 for the four microbial biomarker classes investigated, with sensitivity vs. specificity ranging from 43.9% to 80.2% vs. 87.9% to 97.1%, respectively. Cell-associated Mtb DNA biomarkers exhibited a statistically significant response to antimicrobial therapy, with similar trends observed for cell-free Mtb DNA and protein/peptide antigens. However, most primary studies were assessed as being at high risk of bias.

**Implications of all the available evidence:** Molecular and biochemical microbial blood biomarkers exhibit similar accuracy for detection of tuberculosis disease, with specificity consistently exceeding sensitivity. Cell-associated Mtb DNA biomarkers exhibited a statistically significant response to antimicrobial therapy, with similar trends observed for cell-free Mtb DNA and protein/peptide antigens. These findings should be interpreted cautiously in the light of high risk of bias for many of the primary studies contributing data. Higher quality studies are needed to evaluate this emerging class of tuberculosis biomarkers.

## INTRODUCTION

Tuberculosis is the world’s leading infectious cause of mortality, responsible for an estimated 1.23 million deaths in 2024.^1^ Biomarkers play a key role in clinical management and tuberculosis control, and the World Health Organisation (WHO) has released a range of target product profiles (TPP) outlining high-priority biomarker needs for screening, diagnosis, evaluating likelihood of disease progression and treatment monitoring.^2–5^ Historically, sputum has been the most widely used sample for tuberculosis biomarker testing, but its application is limited to assessment of individuals with suspected or proven pulmonary disease who can expectorate, and challenges include collection difficulties, variable sample quality, infection control issues necessitating decontamination and adverse impacts of viscous sample matrix on assay performance. By contrast, peripheral blood testing offers fewer infection control issues and easier standardization, fulfilling WHO’s prioritization of tuberculosis biomarkers that can be measured in easy-to-obtain samples.^6^

Most existing blood tests for *M. tuberculosis* (Mtb) infection detect host responses to the organism rather than bacillary targets. These indirect measures of Mtb infection may be impaired in sensitivity (false-negative interferon-gamma release assay [IGRA] and tuberculin skin test [TST] results have been documented in young children and HIV-infected individuals with tuberculosis)^7^ and specificity (IGRA may remain positive in individuals who have remote, potentially resolved, Mtb infection but retain memory responses,^8,9^ while host blood transcriptomic biomarkers may yield false-positive results in individuals with intercurrent viral respiratory infections).^10^ Consequently, immunoassays have low positive predictive value for progression to active tuberculosis^11^ and limited utility as biomarkers of response to preventive or full antimicrobial therapy.^12,13^

By contrast, detection of mycobacterial components in the circulation (hereafter termed microbial blood biomarkers) represents a direct measure of Mtb infection, with potential to overcome limitations of immunoassays and address the unmet need for a gold standard test of Mtb infection.^14^ Specific targets investigated include cell-free Mtb DNA (which circulates freely in serum or plasma rather than being contained within host cells), cell-associated Mtb DNA (which is contained within host cells) and Mtb antigens whose composition may broadly be classified as primarily constituting mycobacterial proteins/peptides and/or lipids/glycolipids. To our knowledge, three relevant systematic reviews have been conducted to date, of which two (2007, 2021) report on detection of cell-free Mtb DNA,^15,16^ and one (2011) reports on antigen detection tests.^17^ Multiple primary studies have been published since these meta-analyses were conducted, but up-to-date syntheses incorporating the latest data for all classes of microbial biomarker are lacking. We therefore conducted a systematic review and meta-analysis to evaluate: (1) the prevalence of detection of different classes of microbial blood biomarker among individuals being screened for Mtb infection and tuberculosis disease and healthy controls; (2) diagnostic accuracy of different biomarker classes for detection of tuberculosis disease; (3) biomarker responses to antimicrobial therapy; and (4) positive and negative predictive values of different biomarker classes for progression to tuberculosis disease in asymptomatic individuals, and for relapse following completion of treatment of tuberculosis disease.

## METHODS

### Protocol and Registration

The protocol for the study was registered with the PROSPERO International Prospective Register of Systematic Reviews (CRD420261308367).

### Eligibility Criteria

Studies were eligible for inclusion if they met the following criteria: (1) primary research articles involving human participants of any age, gender, or ethnicity; (2) inclusion of individuals with confirmed or probable active tuberculosis (microbiological and/or clinical diagnosis, pulmonary or extrapulmonary disease), tuberculosis contacts or other asymptomatic individuals, or symptomatic individuals with non-tuberculosis illness. (3) use of blood-derived samples, including whole blood, plasma, serum, buffy coats, leukocytes, or other blood constituents; (4) direct detection of Mtb complex–derived microbial biomarkers in blood, including bacillary DNA, proteins, peptides, lipids, glycolipids, or lipoproteins; and (5) study designs comprising clinical studies, diagnostic accuracy studies, or biomarker studies. Only peer-reviewed articles published between 1^st^ January 1990 and 22^nd^ October 2025, in English were included.

Studies were excluded if they met any of the following criteria: (1) non-human studies, including animal models; (2) *in vitro* studies not involving human-derived samples; (3) studies analysing non-blood samples only; (4) studies investigating host-response assays only; (5) studies exclusively involving detection of intact Mtb organisms through microscopy or culture; (6) assays not specifically targeted to the Mtb complex, such as metagenomic next-generation sequencing; (7) studies detecting Mtb-specific RNA only; (8) case reports or single-patient studies, case series with less than three eligible participants, conference abstracts, narrative reviews, commentaries, or editorials; (9) studies that did not report outcomes in terms of the proportion or number of test-positive individuals studied; and (10) studies published in languages other than English.

### Information sources and search strategy

MEDLINE, EMBASE and Scopus databases were searched using the terms reported in Supplementary Material. Each database was searched from 1st January 1990 until 22 October 2025. To identify additional eligible studies, the reference lists of relevant review articles and eligible publications were also screened.

### Study selection

All records identified through the searches were exported and de-duplicated using Rayyan software^18^ prior to screening. Titles and abstracts were manually screened by two authors (SC, ECC) on Rayyan. Discrepancies were resolved through discussion between the reviewers.

### Data collection and data items

Data extraction was performed independently and manually by three authors (SC, ECC, ARM) using a structured data extraction form implemented in Qualtrics XM (Qualtrics LLC, Provo, UT). Any discrepancies were resolved by discussion to achieve consensus. Extracted data included study site(s), study design, target type (e.g. cell-free Mtb DNA, cell-associated Mtb DNA, antigens), specific target (e.g. IS*6110* DNA, CFP-10 peptide), clinical setting, cohort type, sample processing or extraction method, detection method, cohort size, and the proportion of participants of each pre-defined clinical phenotype (e.g. active tuberculosis, non-tuberculosis illness, IGRA/TST-positive or -negative asymptomatic individuals) testing positive for the target biomarker. For longitudinal studies of individuals being treated for Mtb infection or tuberculosis disease, paired proportions of test-positive participants were recorded at baseline and at completion of antimicrobial treatment, unless follow-up did not extend to this point, in which case data from the the latest reported follow-up time point were utilised. Where required, study investigators were contacted by email to obtain additional information or clarify reported data. To avoid data duplication, studies by the same authors reporting identical detection approaches applied to samples from the same participant cohort were identified, and only the most complete or relevant report was included.

### Assessments of risk of bias and applicability

Risk of bias and applicability were assessed using a modified version of the QUADAS-3 tool^19^ (details presented in Supplementary Material). Artificial intelligence software (Gemini 3 pro) was used to make judgements for each domain, a subset of which were then manually verified by two authors (SC, ECC), working independently. The likelihood of publication bias was investigated through the construction of a Deeks’ funnel plot assessing study precision against performance.^20^

### Synthesis of results

All statistical analyses were performed using Stata software v.19 (StataCorp, College Station, TX). Meta-analyses were conducted in three phases: an estimation of pooled prevalence using cross-sectional data, a bivariate analysis of diagnostic accuracy, and a risk difference analysis of longitudinal data from studies that evaluated response to antimicrobial therapy.

### Estimation of pooled prevalence of biomarker positivity across sub-groups at baseline

To maximise generalisability of our findings, we first estimated the pooled prevalence of target detection (positivity rates) across varying study populations at baseline for all eligible studies (i.e. including studies that only investigated individuals with active tuberculosis or without active tuberculosis). This was done by performing a one-proportion meta-analysis of cross-sectional data. Because proportions can suffer from variance instability—particularly when close to 0 or 1—we applied the Freeman-Tukey double arcsine transformation^21^ to the raw proportions prior to pooling. To account for the hierarchical structure of the data, where multiple targets and/or locations were reported within a single study (resulting in non-independent effect sizes), we employed multilevel random-effects models nested by study and location. Parameters were estimated using Restricted Maximum Likelihood (REML). Following the analysis, the pooled estimates and their 95% confidence intervals (CIs) were back-transformed to conventional proportions for interpretation and presentation in forest plots.^22^ Analyses were stratified by subgroups including HIV status (positive, negative, and not reported) and participant classification (active tuberculosis, non-tuberculosis illness, and IGRA/TST status).

### Diagnostic accuracy and bivariate analysis

The principal diagnostic accuracy measures reported were sensitivity and specificity for detection of active tuberculosis disease, based on the requirement for initiation of full anti-tuberculous treatment rather than microbiological reference standards: the latter (e.g. sputum culture) were not universally available across study sites, and we wished to maximise generalisability by including individuals with a clinical diagnosis of active tuberculosis. The unit of assessment was per-participant. For the assessment of diagnostic performance, we constructed 2×2 contingency tables (True Positive, False Positive, False Negative, True Negative) for the subset of studies that reported prevalence of biomarker positivity for individuals with and without active tuberculosis. Data from the latter group (comprising those with non-tuberculosis illness and asymptomatic individuals irrespective of Mtb infection status) were pooled for this analysis. We utilised a Hierarchical Summary Receiver Operating Characteristic (HSROC) model^23^ implemented via the metandi and midas packages^24^ to jointly synthesise sensitivity and specificity estimates while accounting for the correlation between them and the heterogeneity across studies. This approach yielded pooled estimates for sensitivity and specificity and facilitated the generation of HSROC curves. The proportion of variation in sensitivity and specificity attributable to between-study heterogeneity was calculated from intraclass correlation coefficients and presented as I^2^ values for each estimate separately. The area under the curve (AUC) was calculated using numerical integration (trapezoidal rule) based on the HSROC model parameters.

### Longitudinal analysis of data from studies evaluating response to antimicrobial therapy

To evaluate changes in proportions of individuals who were biomarker-positive following initiation of antimicrobial therapy (follow-up vs. baseline), we calculated the Risk Difference (RD) for longitudinal cohorts. Standard errors for the risk difference were computed adjusting for the paired nature of the data (repeated measures on the same subjects). As the within-subject correlation coefficient was not reported in the primary studies, a conservative correlation (*r*) of 0.5 was assumed for the calculation of paired variances.^25^ A random-effects meta-analysis was then performed to pool the risk differences, stratified by analyte class, with results presented in forest plots. A weighted linear regression was performed to evaluate the relationship between risk difference and post-treatment sampling timepoint, using study cohort sizes as analytic weights. The results are presented in a bubble plot, stratified by biomarker class, along with a fitted trend line and accompanying R-squared value.

### Additional analyses

As an extension of the primary analysis, we conducted a bivariate meta-regression for each analyte class to investigate the following potential sources of heterogeneity: age (adults vs. children), decade of publication (1990s vs. 2000s vs. 2010s vs. 2020s), WHO region (African Region vs. Region of the Americas vs. Eastern Mediterranean Region vs. South-East Asian Region vs. Western Pacific Region), study design (multi-gate vs. single-gate), laboratory detection method (for Mtb DNA targets: endpoint polymerase chain reaction [PCR] vs. quantitative PCR [qPCR] vs. digital PCR [dPCR] vs. clustered regularly interspaced short palindromic repeats [CRISPR]-driven vs. sequencing approaches; for antigen targets, enzyme-linked immunosorbent assay [ELISA] vs. electrochemiluminescence assay [ECLIA] vs. mass spectrometry vs. nanoparticle-mediated detection), sample type (for cell-associated Mtb DNA, PBMC vs. whole blood vs. undifferentiated leucocytes; for other targets, serum vs. plasma); and, for antigen targets only, detection of free vs. antibody-bound antigen. For each analysis, subgroups were excluded where they did not meet the minimum requirement of four studies for bivariate modelling. Within this framework, generalised linear mixed-effects models were used to generate a global Wald p-value to assess the statistical difference in diagnostic accuracy between subgroups with >2 levels, or pairwise Wald p-values where subgroups had only two levels. For bivariate analyses of each analyte class, we also attempted to conduct sensitivity analyses excluding studies assessed to be of high risk of bias, where sufficient studies (≥5) remained.

### Role of the funding source

The study was funded by Barts Charity (ref G-002700), The Medical College of Saint Bartholomew’s Hospital Trust and a Wellcome HARP Doctoral Fellowship (ref 227532/Z/23/Z). No funder had any role in writing the manuscript or in the decision to submit it for publication. The authors were not paid to write this article by a pharmaceutical company or other agency. The authors were not precluded from accessing data in the study, and they accept responsibility to submit for publication.

## RESULTS

### Study selection

The study selection process is summarised in Figure 1. Our search identified 10,994 records, including 1,813 from PubMed, 2,825 from EMBASE, 6,347 from Scopus and nine from reference lists of relevant review articles and other publications. After removal of 3,542 duplicate records, 7,452 unique records were screened at the title and abstract level. Of these, 7,284 were excluded based on title and abstract review. The most common reasons for exclusion were studies investigating host response biomarkers (as opposed to microbial biomarkers), studies investigating diseases other than tuberculosis, case reports, studies detecting Mtb targets in samples other than blood, review articles, genetic studies, animal studies and studies not presented in English.

**Figure 1.**
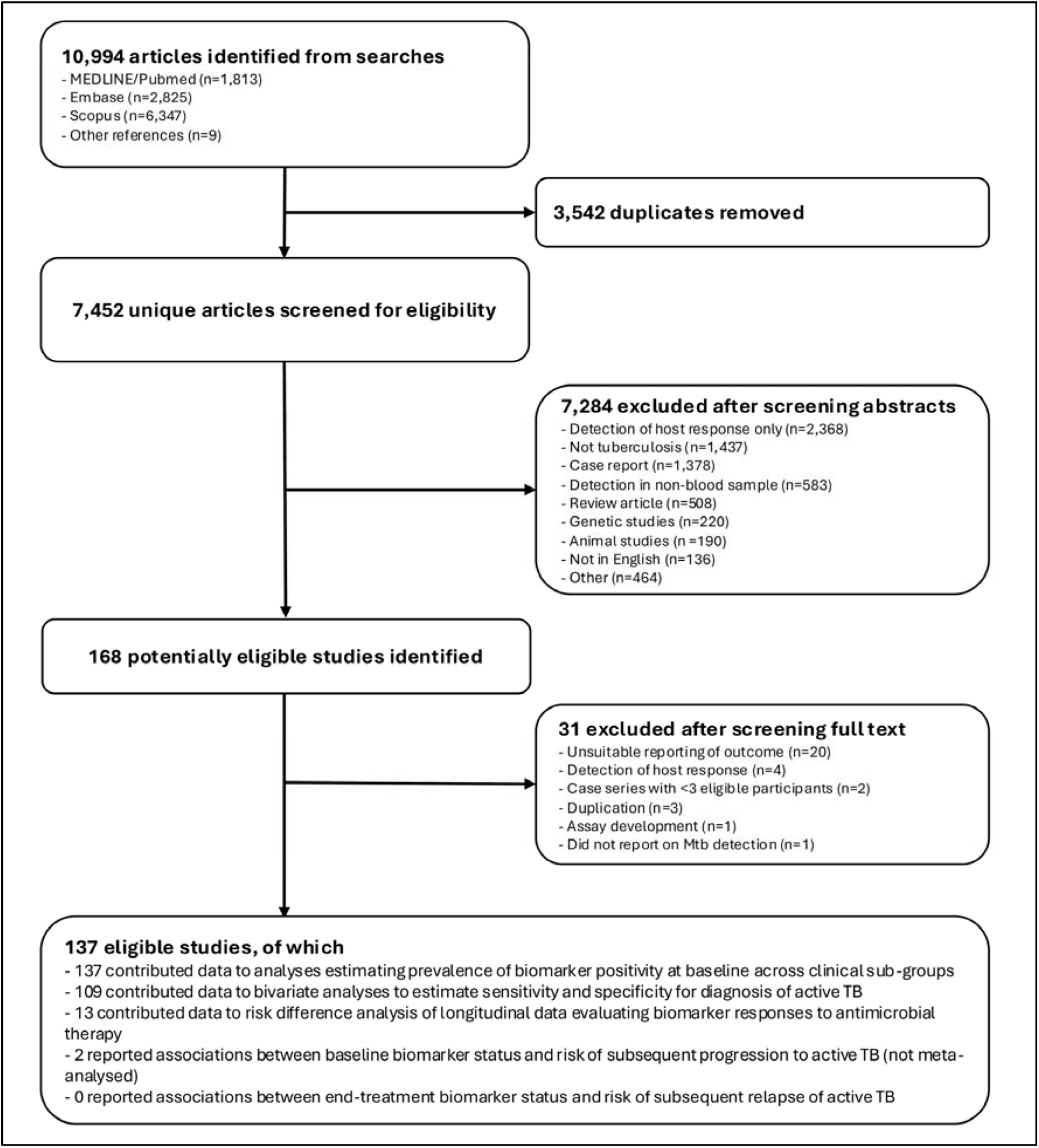
Flow chart of study selection

A total of 168 full-text articles were retrieved and assessed for eligibility. Of these, 31 studies were excluded after full-text review; reasons for their exclusion are listed in Table S1, Supplementary Material. The remaining 137 studies met eligibility criteria for this review, of which all contributed data to analyses estimating prevalence of biomarker positivity at baseline across clinical sub-groups. Additionally, 109 of these studies contributed data to bivariate analyses estimating sensitivity and specificity for diagnosis of active tuberculosis; 13 contributed data to risk difference analysis of longitudinal data evaluating biomarker responses to antimicrobial therapy; and 2 reported associations between baseline biomarker status and risk of subsequent progression to active tuberculosis. No studies investigating associations between end-treatment biomarker status and risk of subsequent relapse of active tuberculosis were identified.

### Characteristics of studies, participants, microbial blood biomarkers and detection methods

Characteristics of the 137 studies contributing data to this meta-analysis are presented in Table 1. All included a cross-sectional component reporting biomarker status at baseline; a subset of 13 studies also included a nested longitudinal component. Studies were conducted in 37 different countries on 5 continents and enrolled children and adults of both sexes with a wide range of clinical phenotypes, including symptomatic individuals with active tuberculosis disease or non-tuberculosis illness as well as asymptomatic individuals, among whom both IGRA/TST-positive and -negative individuals were represented.

**Table 1.**
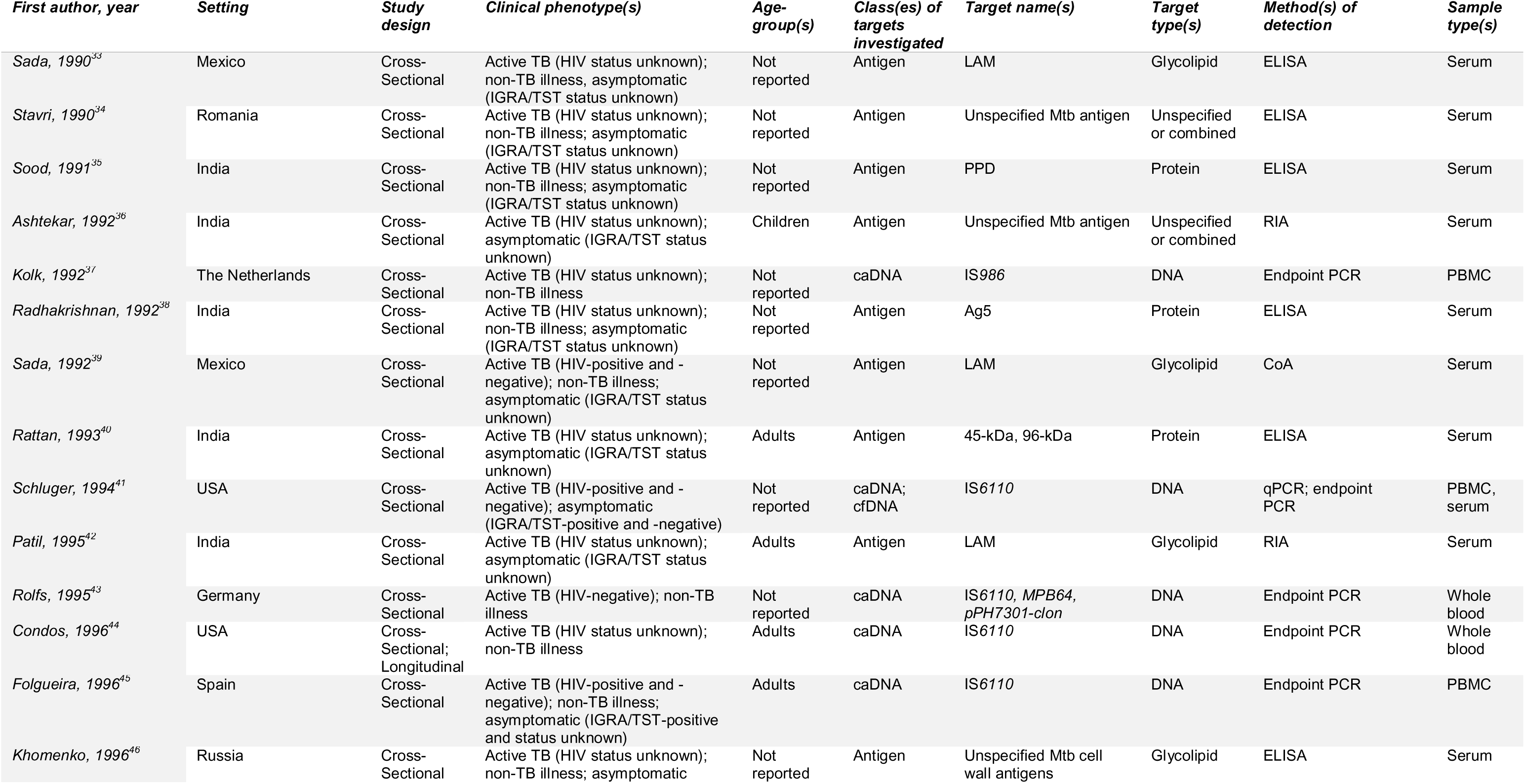
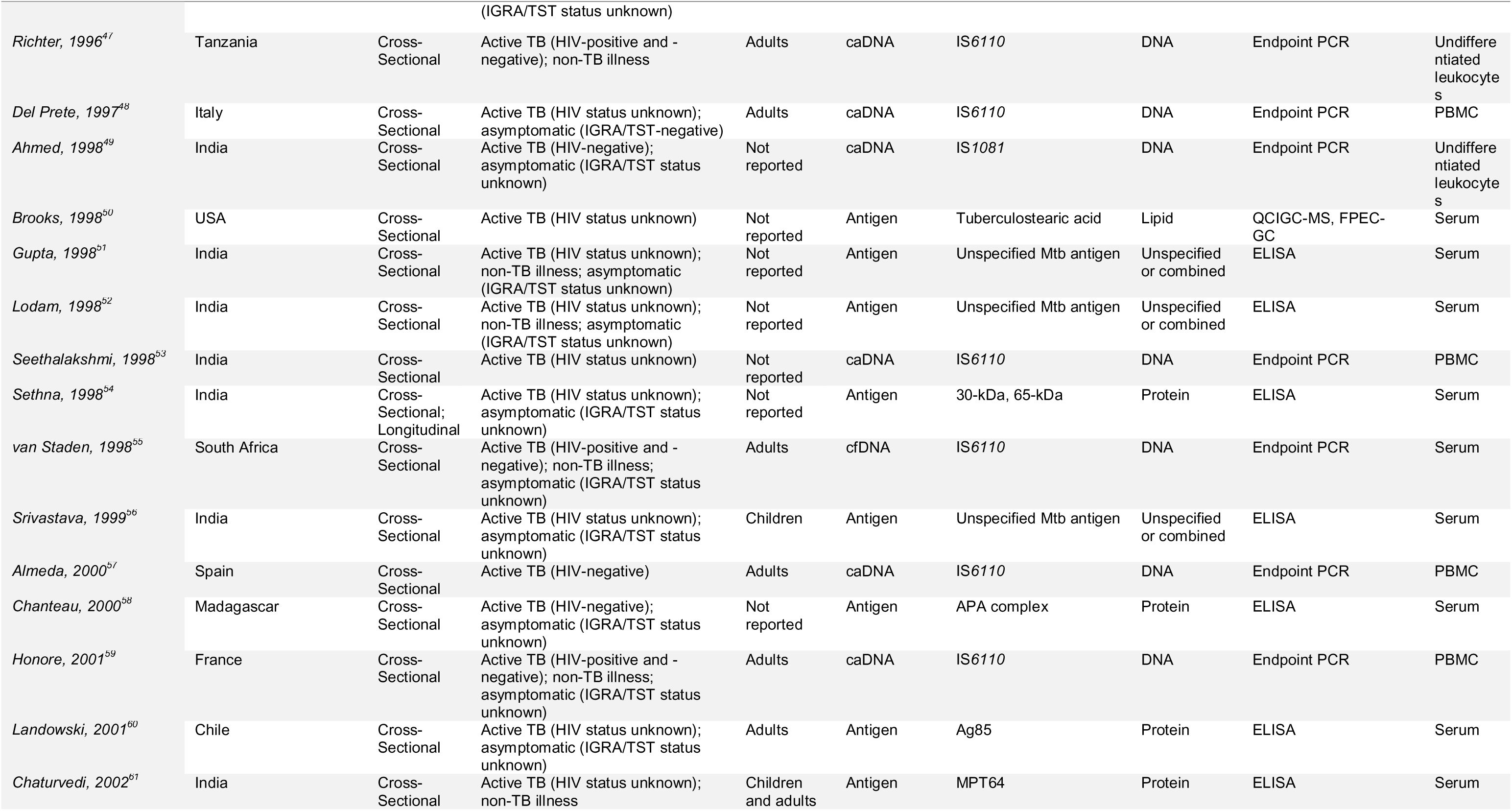
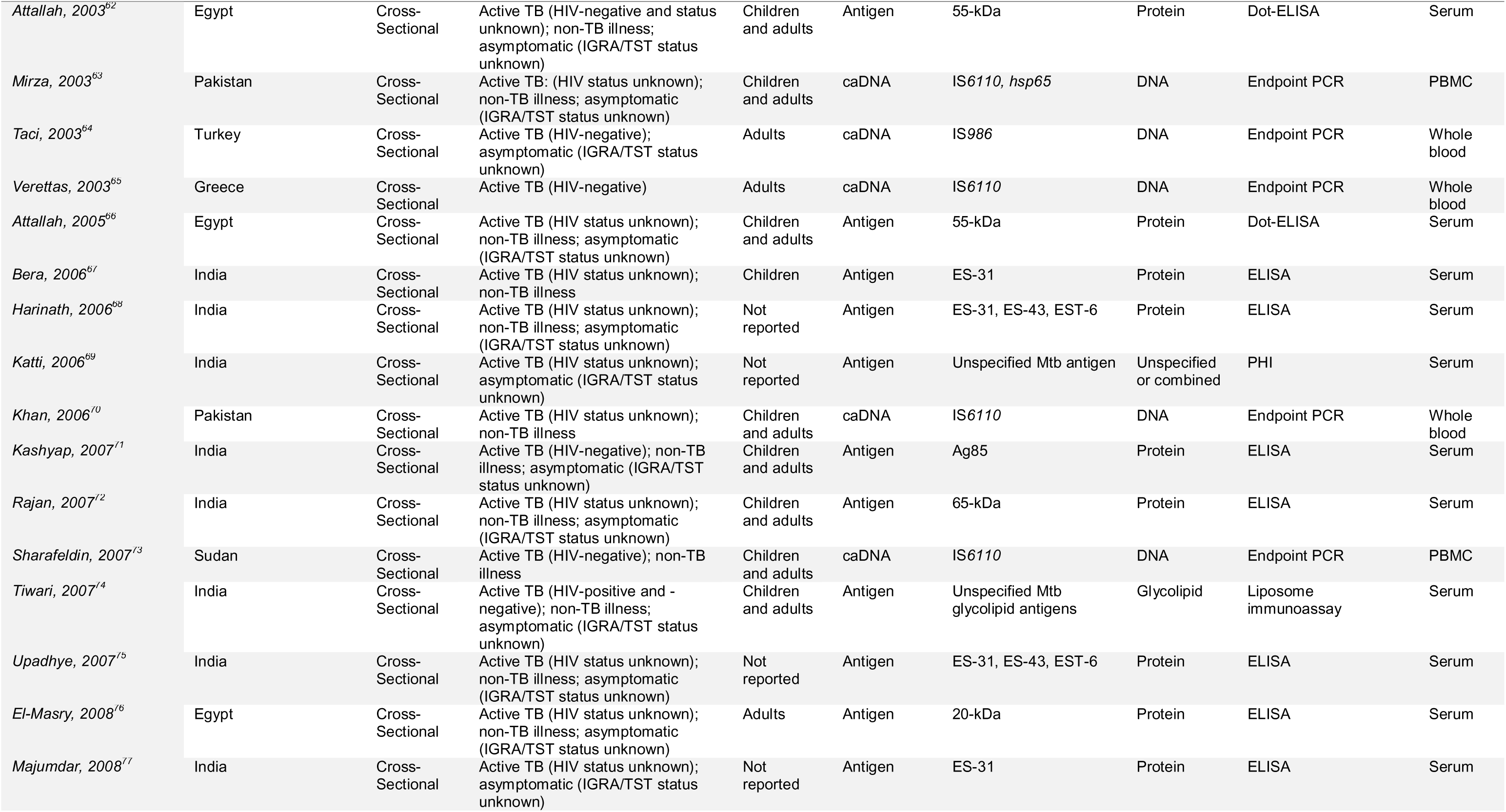
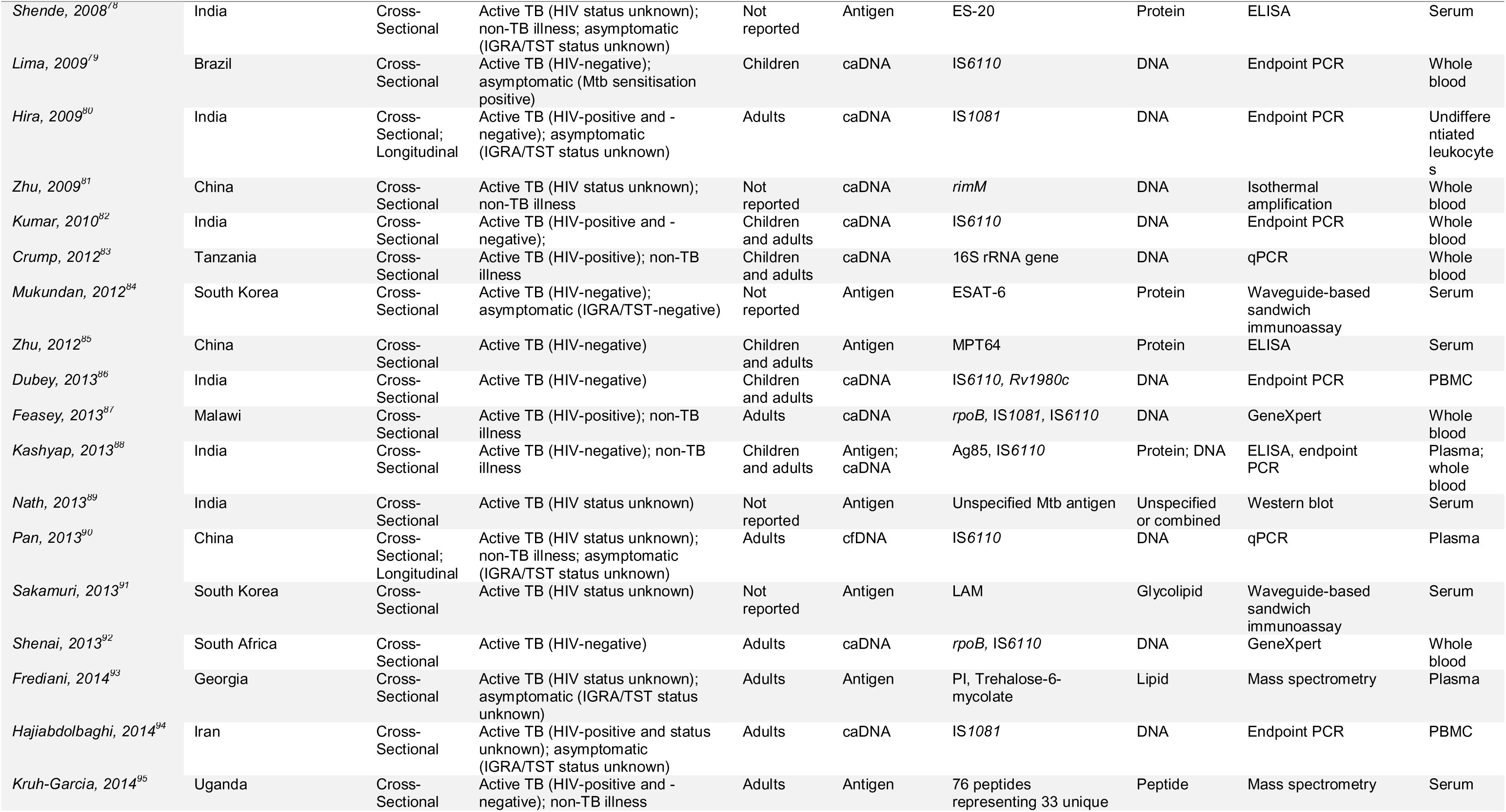
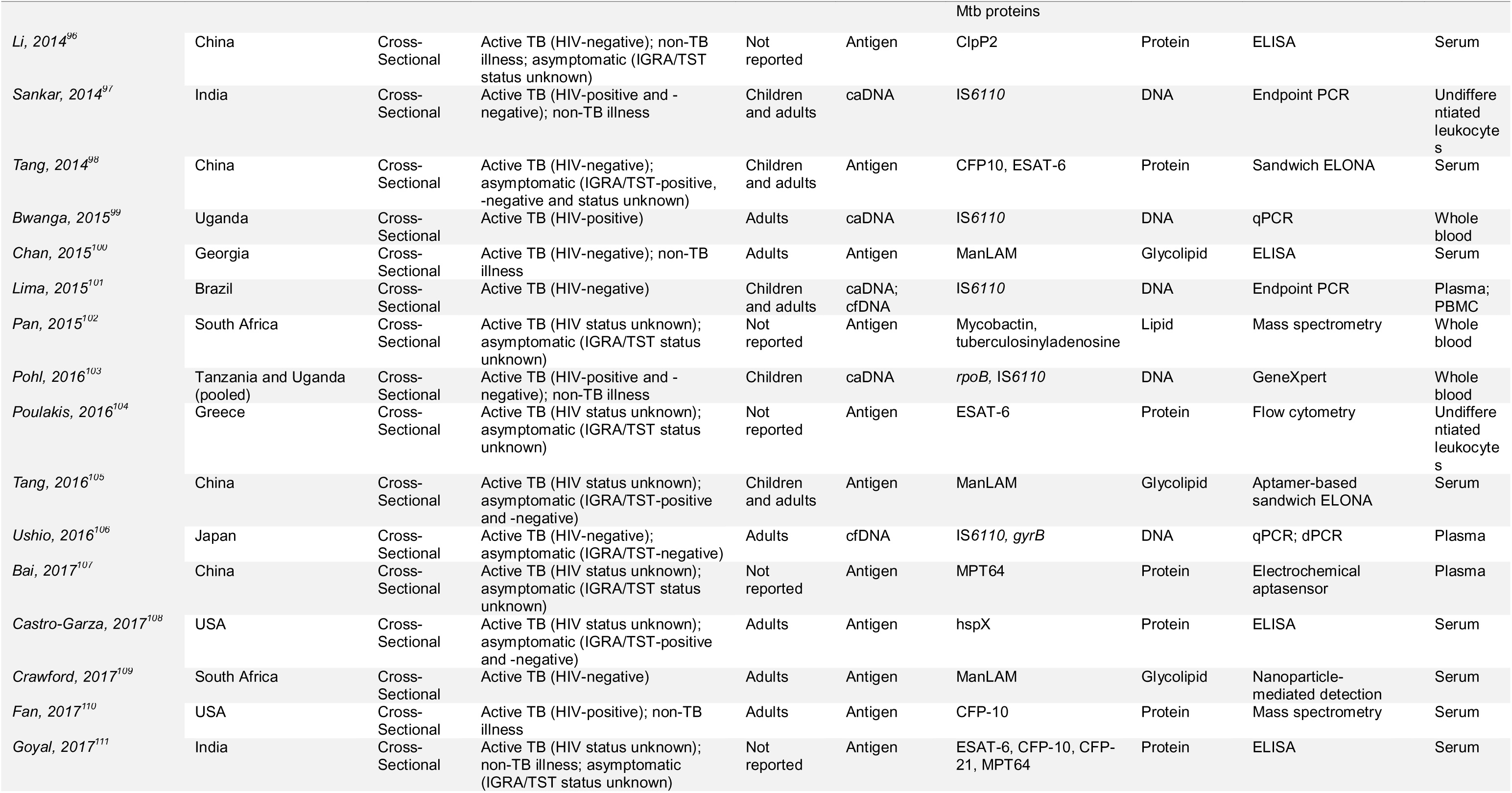
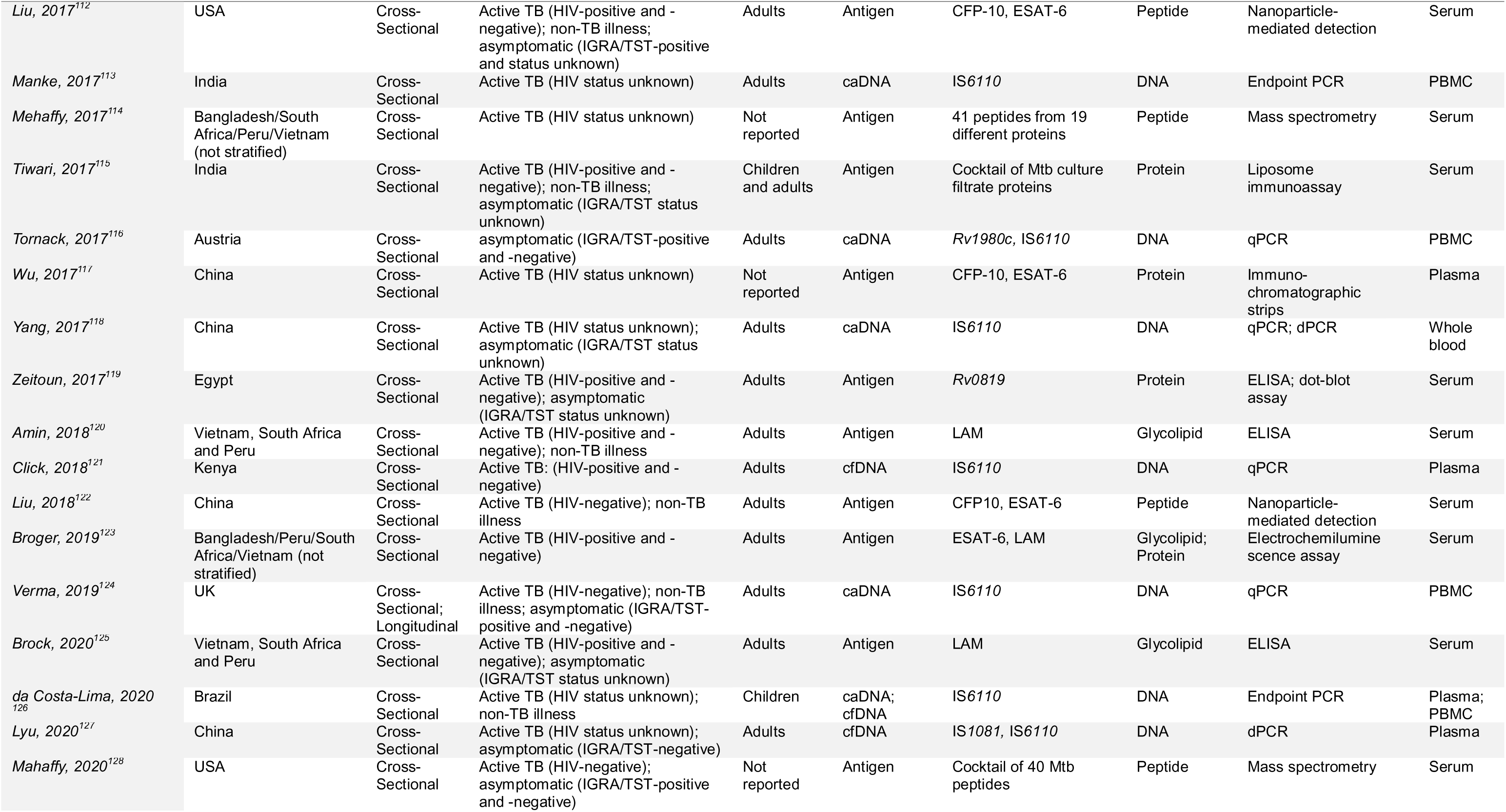
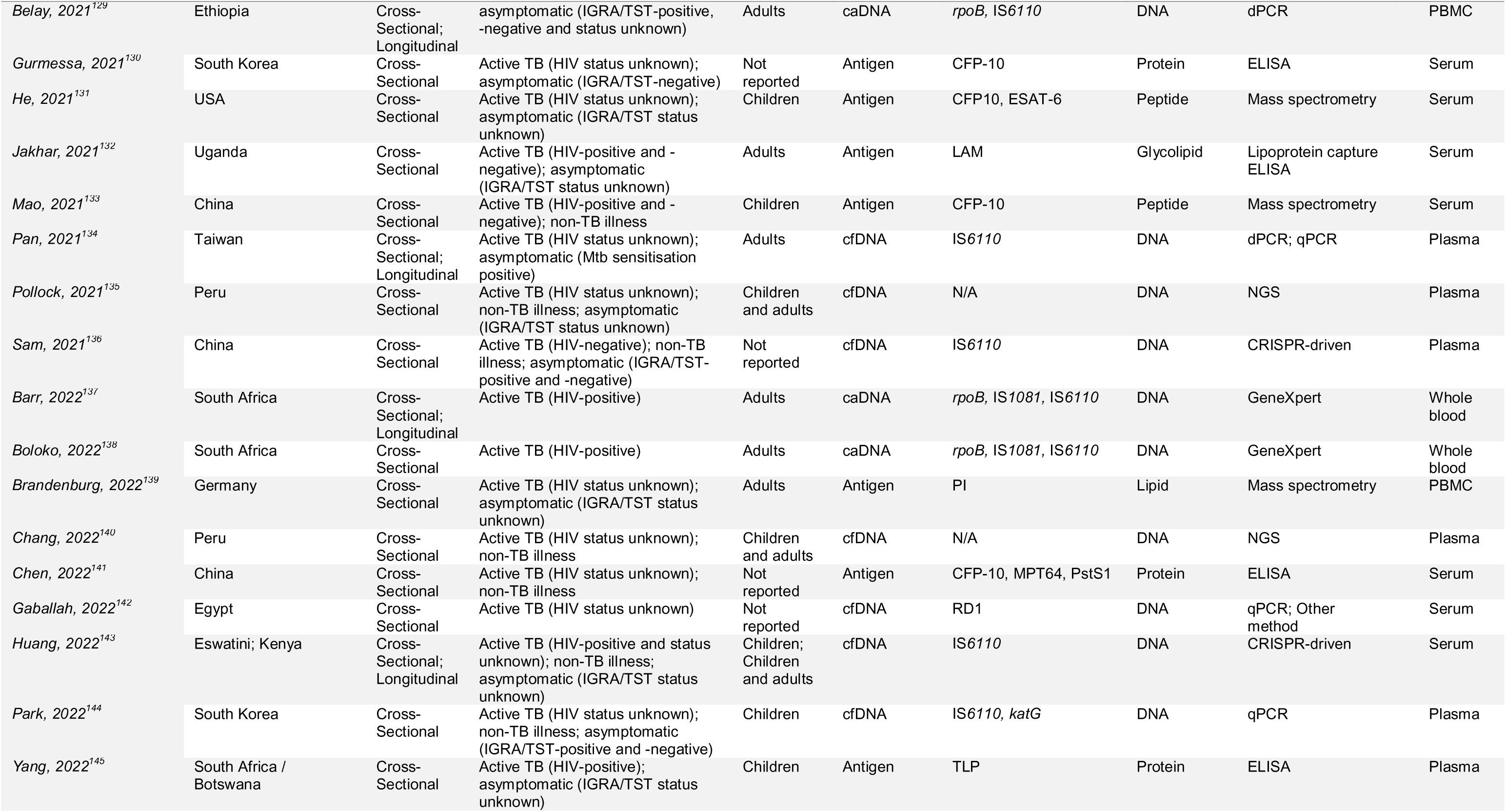
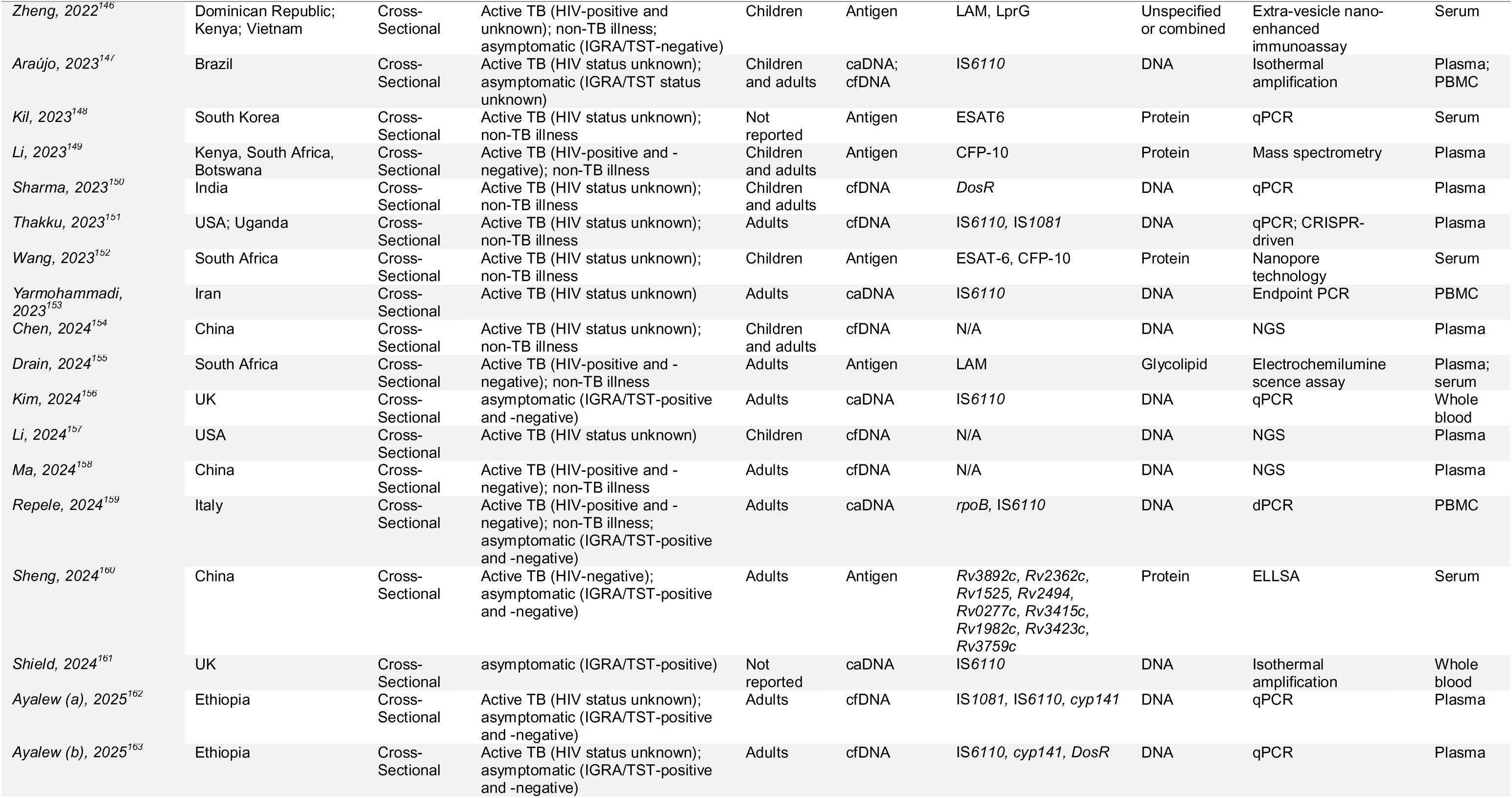
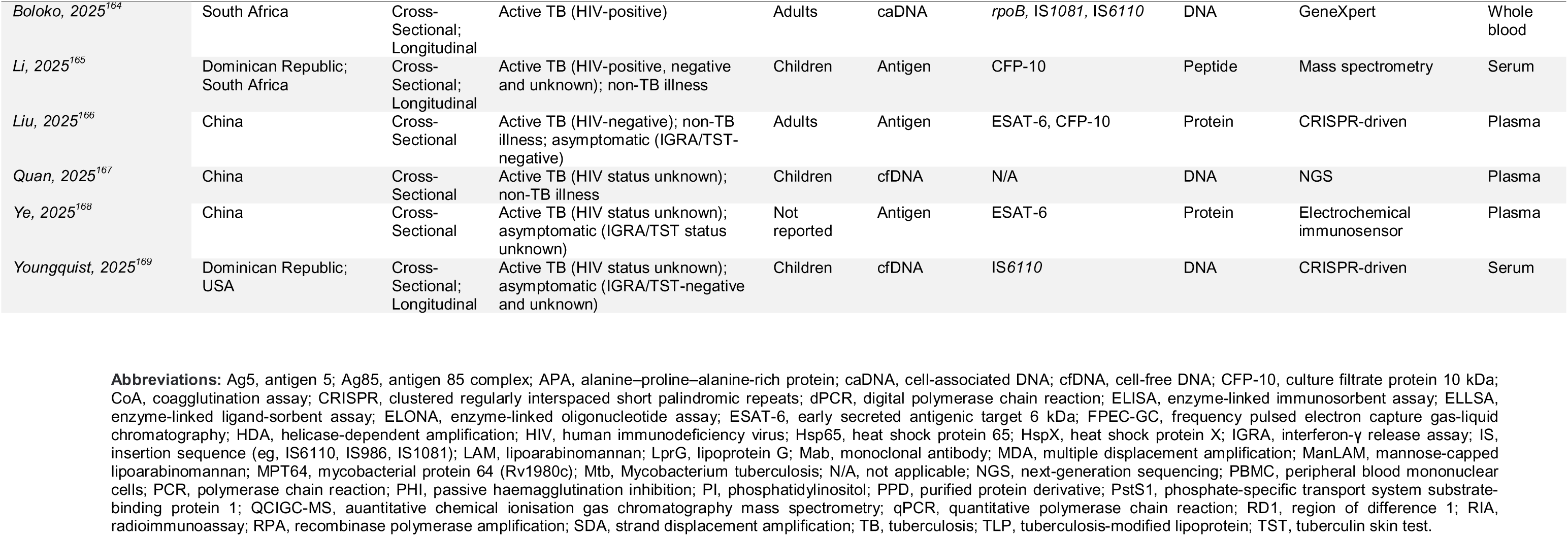
Characteristics of included studies.

A diverse range of targets were evaluated: 21 studies investigated cell-free Mtb DNA only, 39 studies investigated cell-associated Mtb DNA only, 45 studies investigated protein/peptide antigens only, 16 studies investigated lipid/glycolipid antigens only, and 10 studies investigated unspecified or combined antigens only. Six studies investigated two or more classes of microbial analyte. These targets were detected using a wide range of methods. Earlier studies in the field typically used endpoint PCR to detect cell-associated multicopy Mtb DNA targets (e.g. IS*6110*, IS*1081*) in whole blood or PBMC, or ELISA to detect glycolipid antigens (e.g. lipoarabinomannan [LAM], mannose-capped LAM [ManLAM]) protein/peptide antigens (e.g. ESAT-6, CFP-10) or unspecified / undifferentiated Mtb antigens in serum or plasma. As the field evolved, qPCR, digital PCR, CRISPR-based technologies and next generation sequencing were increasingly employed to detect Mtb DNA targets, and the greater sensitivity of some of these methods has allowed detection of cell-free Mtb DNA targets, predominantly in plasma. Isothermal amplification methods have also been investigated, with a view to developing point-of-care tests that can detect Mtb DNA targets in lower income settings where more costly technologies are not readily available. Methods for detection of Mtb antigens also evolved over time, with mass spectrometry, electrochemiluminescence immunoassays (ECLIA) and nanoparticle-mediated detection being increasingly used to detect Mtb antigens in plasma and serum.

### Risk of bias and applicability

Assessments of the risk of bias and applicability of study findings are presented in Table S2, Supplementary Material. Most studies were classified as being at high risk of bias (23/25 [92.0%] cell-free Mtb DNA studies; 35/44 [79.5%] cell-associated Mtb DNA studies; 45/46 [97.8%] protein/peptide antigen studies; 17/18 [94.4%] lipid/glycolipid antigen studies; and 9/10 [90.0%] studies of unspecified or combined antigens). Most frequently, this judgement arose from the ‘participants’ domain, reflecting the fact that many studies had a multi-gate design, in which cases and controls were recruited separately and according to different eligibility criteria.

Most studies were also classified as giving cause for high concern re applicability (23/25 [92.0%] cell-free Mtb DNA studies; 30/44 [68.2%] cell-associated Mtb DNA studies; 44/46 [95.7%] protein/peptide antigen studies; 16/18 [88.9%] lipid/glycolipid studies antigen and 8/10 [80.0%] studies of unspecified or combined antigens). Again, this judgement most frequently arose from the ‘participants’ domain, reflecting concerns that participant populations enrolled in many studies (often those with case-control design) did not match the intended populations for a tuberculosis detection biomarker.

### Estimating prevalence of biomarker positivity at baseline across clinical sub-groups

A total of 137 studies, yielding 200 unique study / setting / biomarker combinations (termed ‘investigations’ below), contributed data to analyses estimating prevalence of biomarker positivity at baseline across different clinical groups. Table 2 presents pooled estimates of biomarker positivity for five classes of microbial biomarker (cell-free Mtb DNA, cell-associated Mtb DNA, protein/peptide antigens, lipid/glycolipid antigens and unspecified or combined antigens) among study participants with and without active tuberculosis, overall and by sub-group. Point estimates for pooled prevalence of biomarker positivity among individuals with active tuberculosis ranged from 0.46 to 0.77 overall; from 0.33 to 0.77 among HIV-uninfected individuals; from 0.45 to 0.84 among HIV-infected individuals; and from 0.64 to 0.75 among individuals with unknown HIV status. In comparison, point estimates for pooled prevalence of biomarker positivity among individuals without vs. with active tuberculosis tended to be numerically lower, ranging from 0.05 to 0.15 overall; from 0.03 to 0.27 for those with non-tuberculosis illness; from 0.07 to 0.60 for asymptomatic IGRA/TST-positive individuals; from 0.06 to 0.13 for asymptomatic IGRA/TST-negative individuals; and from 0.04 to 0.08 for asymptomatic individuals with unknown IGRA/TST status. Within each clinical group, point estimates for prevalence of biomarker positivity were broadly similar across different classes of analyte investigated, except for asymptomatic IGRA/TST-positive individuals in whom the point estimates for prevalence of biomarker positivity were numerically lower for cell-free Mtb DNA (0.07) than for cell-associated Mtb DNA (0.42) and protein/peptide antigens (0.60). A high degree of heterogeneity was observed (*I*^2^ >50% in 36/40 analyses for which there were sufficient observations to meta-analyse). Forest plots displaying prevalence of biomarker positivity at baseline for individual studies and estimates of pooled prevalence for each biomarker class and clinical group for which sufficient studies were available to meta-analyse are presented in Figures S1 to S40, Supplementary Material; the Supplementary Figure number of the forest plot corresponding to each analysis is indicated in each cell of Table 2.

**Table 2.**
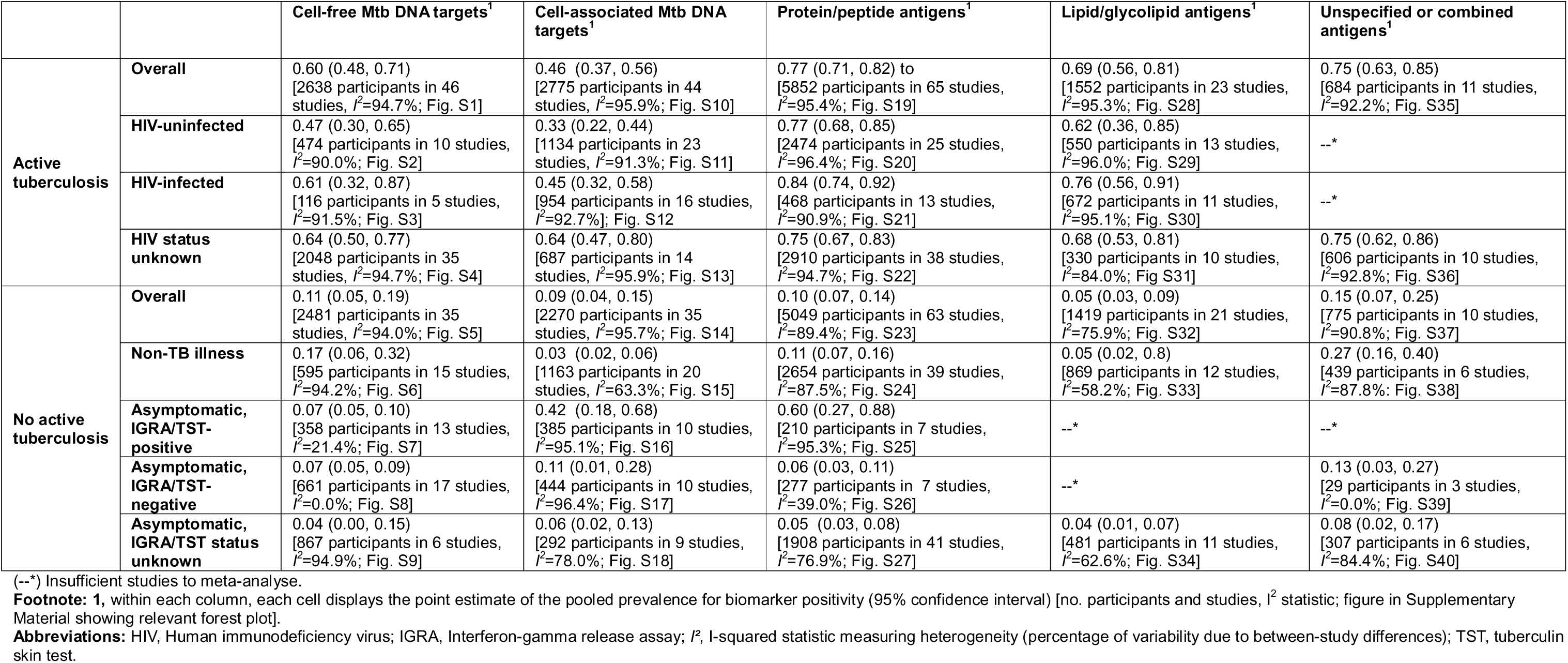
Pooled estimates for prevalence of biomarker positivity in individuals with and without active tuberculosis, overall and by sub-group.

### Bivariate analyses to estimate sensitivity and specificity for diagnosis of active tuberculosis

A total of 109 studies, reporting results of 159 investigations, contributed data to bivariate analyses estimating sensitivity and specificity of different microbial biomarker classes for the diagnosis of active tuberculosis. ROC curves showing diagnostic accuracy for each biomarker class are presented in Figure 2, and values for sensitivity, specificity and *I*^2^ statistics are presented in Table S3, Supplementary Material. For cell-free Mtb DNA targets, the AUC was 0.87 (95% CI 0.84 to 0.89; 4,878 samples in 34 investigations), with sensitivity 61.5% (95% CI 51.0% to 71.0%; *I*^2^=30%) and specificity 93.0% (95% CI 88.1% to 96.1%; *I*^2^=43%). For cell-associated Mtb DNA targets, the AUC was 0.93 (95% CI 0.90 to 0.95; 3,589 samples in 32 investigations), with sensitivity 43.9% (95% CI 29.4% to 59.4%; *I*^2^=47%) and specificity 97.1% (95% CI 94.5% to 98.5%; *I*^2^=36%). For protein/peptide targets, the AUC was 0.94 (95% CI 0.92 to 0.96; 10,260 samples in 61 investigations), with sensitivity 78.9% (95% CI 73.2% to 83.6%; *I*^2^=29%) and specificity 92.9% (95% CI 90.7% to 94.5%; *I*^2^=21%). For lipid/glycolipid targets, the AUC was 0.96 (95% CI 0.94 to 0.97; 3,287 samples in 22 investigations), with sensitivity 68.6% (95% CI 54.1% to 80.3%; *I*^2^=37%) and specificity 97.0% (95% CI 94.0% to 98.5; *I*^2^=32%). For unspecified or combined antigen targets, AUC was 0.91 (95% CI 0.88 to 0.93; 1,417 samples in 10 investigations), with sensitivity 80.2% (95% CI 67.1% to 88.9%) and specificity 87.9% (95% CI 76.0% to 94.3).

**Figure 2.**
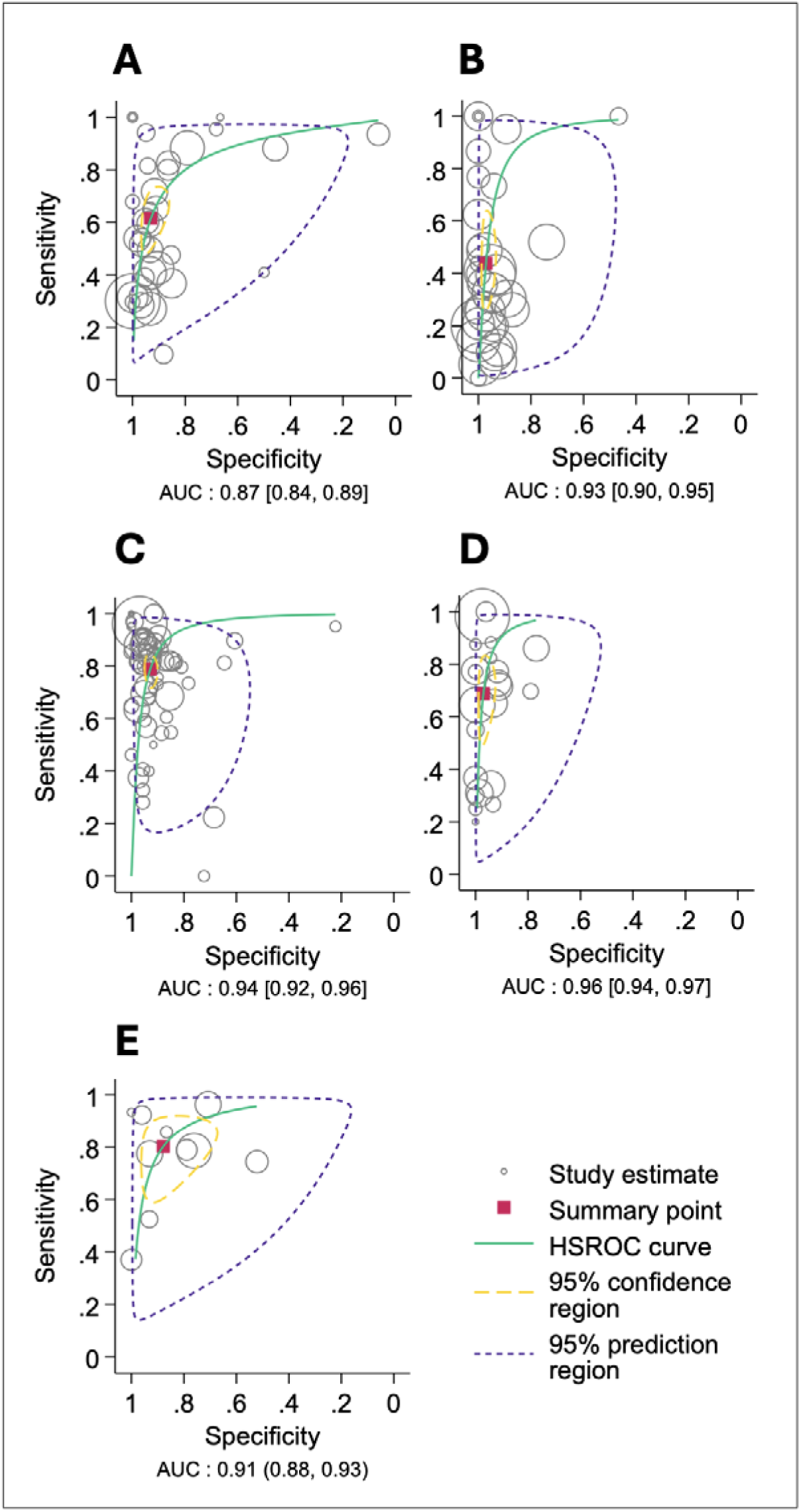
Receiver operating characteristic curves showing diagnostic accuracy of different microbial blood biomarker classes for active tuberculosis. A, cell-free Mtb DNA. B, cell-associated Mtb DNA. C, protein or peptide antigens. D, lipid or glycolipid antigens. E, unspecified or combined antigens. AUC, area under the curve. HSROC, Hierarchical Summary Receiver Operating Characteristic.

### Longitudinal analysis of data from studies evaluating response to antimicrobial therapy

A total of 13 studies, reporting results of 14 investigations, contributed data to risk difference analysis of longitudinal data from studies evaluating biomarker responses to antimicrobial therapy. The most common timepoint for follow-up was at 24 weeks after initiation of treatment, but some studies evaluated participants considerably sooner. These data are presented in Figure 3, along with forest plots displaying proportions of participants who were biomarker-positive before vs. after initiation of antimicrobial therapy for individual studies and estimates of pooled risk difference for each biomarker class. For cell-free Mtb DNA targets, the pooled risk difference was - 0.44 (95% CI -0.89 to 0.01; *I*^2^=97%; 68 samples in 5 investigations). For cell-associated Mtb DNA targets, the pooled risk difference was -0.46 (95% CI -0.88 to - 0.03; *I*^2^=98%; 89 samples in 5 investigations). For protein/peptide antigens, the pooled risk difference was -0.24 (95% CI -0.75 to 0.28); *I*^2^=98%; 160 samples in 4 investigations). Weighted linear regression revealed no relationship between risk difference and timing of post-treatment sampling (R^2^=0.00; Figure S42).

**Figure 3.**
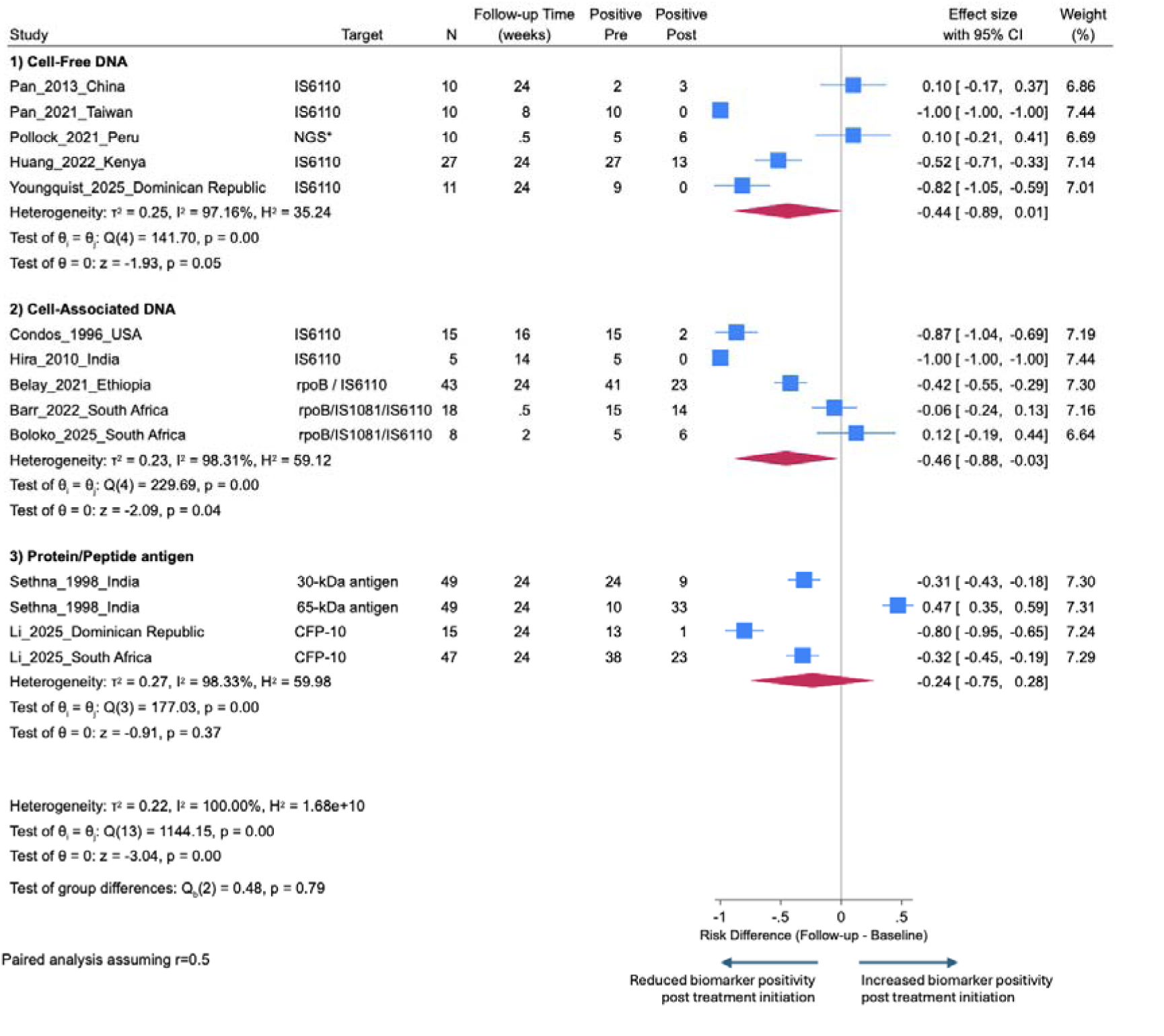
Forest plots of data from longitudinal studies showing risk differences for detection of microbial blood biomarkers after vs. before initiation of antimicrobial therapy.

### Longitudinal analysis of data from studies evaluating risk of progression to active tuberculosis

We identified two studies evaluating the predictive value of biomarker positivity in asymptomatic individuals for risk of progression to active tuberculosis. In one study,^26^ 2/3 asymptomatic participants who had cell-associated Mtb DNA detectable at baseline progressed to active tuberculosis by 7-month follow-up, compared with 0/15 individuals in whom cell-associated Mtb DNA was undetectable at baseline. In another study,^27^ 6/11 asymptomatic individuals who had cell-associated Mtb DNA detectable at baseline progressed to active tuberculosis by 12-month follow-up, compared with 0/8 individuals in whom cell-associated Mtb DNA was undetectable at baseline. Meta-analysis of these data was not attempted due to the small number of studies.

### Prediction of relapse among individuals who completed antimicrobial therapy

No studies evaluating the prognostic significance of end-treatment biomarker status for tuberculosis recurrence or relapse were identified.

### Subgroup analyses

As an extension of the primary analysis, we conducted bivariate meta-regression to investigate sources of between-study heterogeneity and identify potential subgroup factors which modified estimates of sensitivity and specificity for each group of analytes. Results are presented in Tables S4-S7, Supplementary Material. Overall, we observed moderate between-study heterogeneity across all four biomarker classes (*I*^2^ estimates for sensitivity and specificity mostly 25-50%), with the proportion attributable to test threshold effect being relatively low (2-32% for the different biomarker classes). Sample type significantly influenced test performance in protein/peptide antigen studies: for antigens investigated in plasma samples (360 samples in 6 investigations), sensitivity was 90.8% (95% CI 79.7% to 96.1%; *I*^2^=15%) and specificity was 96.7% (95% CI 90.3% to 98.9%; *I*^2^=10%), while for biomarkers investigated in serum samples (9,776 samples in 54 investigations), sensitivity was 76.9% (95%CI 70.7% to 82.2%) and specificity was 92.7% (95% CI 90.5% to 94.4%; P for interaction=0.045). WHO region significantly influenced test performance in cell-free DNA studies: for biomarkers investigated in WPRO region (1,924 samples in 14 investigations), sensitivity was 48.2% (37.9, 58.6; *I*^2^=15%) and specificity was 93.1 (89.0, 95.7; *I*^2^=11%), while for biomarkers investigated in AFRO region (1,807 samples in 13 investigations), sensitivity was 70.4% (57.3, 80.9; *I*^2^=22%) and specificity was 91.3 (84.0, 95.5; *I*^2^=22%) and for biomarkers investigated in AMRO region (1,062 samples in 6 investigations), sensitivity was 78.7 (49.3, 93.3; *I*^2^=41%) and specificity was 95.1 (46.6, 99.8; *I*^2^=80%); P for interaction=0.039. P values for interaction for all other subgroup factors investigated across all analyte classes were >0.05.

### Sensitivity analysis

For cell-associated Mtb DNA studies, we conducted a bivariate sensitivity analysis excluding those assessed to be at high risk of bias (35/44). For the 9 remaining investigations with 1,297 samples, the AUC was 0.94 (95% CI 0.91, 0.96), with sensitivity 31.6% (95% CI 16.0% to 52.8%; *I*^2^=34%) and specificity 96.7% (95% CI 94.1% to 98.2%; *I*^2^=10%); Table S3, Supplementary Material). There were insufficient studies (<4) assessed as being at low risk of bias to perform sensitivity analysis for other biomarker classes.

### Test of publication bias

A Deeks’ funnel plot for studies contributing to the bivariate analysis is presented in Figure S41, Supplementary Material. This shows significant asymmetry (P<0.01) with studies concentrated on the high, right side of the regression line (high performance; smaller studies).

## DISCUSSION

To our knowledge, this study represents the most comprehensive systematic review and meta-analysis of data from studies of microbial blood biomarkers of Mtb infection and tuberculosis disease conducted to date, and the first such meta-analysis to synthesise data from studies investigating detection of cell-associated Mtb DNA in blood. Within each clinical group investigated, point estimates for prevalence of microbial blood biomarker positivity were broadly similar for molecular vs. biochemical targets, tending to be numerically higher for symptomatic individuals with tuberculosis disease vs. non-tuberculosis illness, and for IGRA/TST-positive vs. -negative asymptomatic individuals. Bivariate analysis revealed promising AUC values, but Cell-associated Mtb DNA biomarkers exhibited a statistically significant response to antimicrobial therapy, with similar trends observed for cell-free Mtb DNA and protein/peptide antigens. However, most primary studies were assessed as being at high risk of bias. Insufficient studies investigating prognostic value of microbial blood biomarkers for disease progression and relapse were identified to allow meta-analysis.

AUC values relating to diagnostic accuracy of microbial blood biomarkers for tuberculosis disease reported here (0.87 to 0.96 for the four microbial biomarker classes investigated) compare favourably with those published for IGRA (0.73 to 0.93),^28,29^ whole blood transcriptional signatures (0.70 to 0.77)^30^ and serological tests (0.84 to 0.89),^31^ with the best-performing candidates fulfilling WHO TPP criteria for diagnostic tests for tuberculosis.^3^ However, estimates of sensitivity for detection of tuberculosis disease (ranging from 43.9% to 80.2%) were consistently lower than those of specificity (ranging from 92.9% to 97.1%). Limited sensitivity may reflect very low circulating concentrations of bacillary targets in some individuals with tuberculosis disease (e.g. those with a low Mtb load in tissue), falling below the lower limit of detection for some assays. Imperfect specificity might reflect confounding due to circulating antigens from non-tuberculous mycobacteria, undiagnosed asymptomatic Mtb infection in symptomatic individuals classified as having a non-tuberculosis illness, cross-contamination or (for molecular targets) non-specific PCR amplification.

Our study has some limitations. Most primary studies contributing data to this meta-analysis were assessed as being at high risk of bias, frequently reflecting their multi-gate or case-control design, which is likely to have introduced spectrum bias due to inclusion of extreme phenotypes. In anticipation of this limitation, we pre-specified sensitivity analyses excluding studies at high risk of bias. For studies investigating cell-associated Mtb DNA, this sensitivity analysis yielded a similar AUC value (0.94, 95% CI 0.91 to 0.96) to the primary analysis including all studies (0.93, 95% CI 0.90 to 0.95; Table S3, Supplementary Material). However, a paucity of studies assessed as being at low risk of bias precluded analogous sensitivity analyses for other analyte classes. We also did meta-regression to compare results of studies utilising single-gate vs. multi-gate designs, which might be expected to be at lower vs. higher risk of bias, respectively. These analyses did not reveal evidence of sub-group effects for any analyte class investigated (P values for interaction all _≥_0.05; Tables S4 to S7, Supplementary Material). Additionally, a moderate degree of heterogeneity was observed within analyses for different classes of microbial target: this likely reflects the wide range of different targets, detection methods and clinical phenotypes investigated for each target class. By contrast, results of meta-analyses were broadly similar across analyte classes, supporting the biological validity of this approach, whose inclusivity maximises statistical power. Our *a priori* decision to pool results from different members of a single analyte class is in keeping with that of previous studies in this field,^16,17^ and reflects that fact that targets within each analyte class are structurally similar and frequently detected using similar platforms (e.g. Mtb DNA targets were commonly detected by PCR, Mtb antigens were commonly detected by ELISA). We also highlight that the asymmetrical appearance of the Deeks’ funnel plot, consistent with a lack of larger studies reporting low diagnostic performance, might reflect publication bias.

Our study has several strengths. Our search was designed to be highly sensitive, and our eligibility criteria were broad, including four distinct classes of microbial analyte, two of which were molecular and two biochemical. Generalisability of our findings is maximised by inclusion of a wide range of studies in our initial estimates of prevalence of biomarker positivity, some of which were not included in bivariate analysis (which was necessarily restricted to studies that reported data on both cases and controls). We conducted meta-regression to explore reasons for heterogeneity, and complemented analysis of cross-sectional data with prospective analyses to evaluate potential use cases relating to therapeutic monitoring. Our findings also shed some light on the biology of Mtb infection and tuberculosis disease. IGRA/TST-positive asymptomatic participants were often found to be microbial biomarker-negative, consistent with the hypothesis that a significant proportion of IGRA/TST-positive asymptomatic individuals may have potentially resolved Mtb infection with retention of memory T cell responses.^8,9,32^ Reversion of microbial biomarker status from positive to negative following initiation of antimicrobial therapy was also a common occurrence: this contrasts with results from longitudinal studies of host response biomarkers in patients receiving antimicrobial therapy,^12,13^ and highlights the potential for use of microbial blood biomarkers to monitor therapeutic responses. The observation that microbial biomarker levels decline after initiation of antimicrobial therapy is also consistent with the hypothesis that detection of bacillary material in blood represents the presence of live organisms within the host.

In conclusion, this systematic review and meta-analysis of aggregate data highlights the potential value of microbial blood biomarkers of Mtb infection and tuberculosis disease for diagnosis of active tuberculosis and in monitoring response to antimicrobial therapy. However, high quality head-to-head studies utilising single-gate designs are required to assess accuracy and utility of the most promising candidate tests for the two use cases above. Future research should also address the lack of studies investigating the prognostic value of these biomarkers for progression to tuberculosis disease in Mtb-infected asymptomatic individuals, and for relapse in individuals who have completed antimicrobial therapy.

## Data Availability

The study dataset is available from

## Contributors

Conceptualisation: SC and ARM. Data extraction and curation: SC, ECC, DAJ and ARM; SC and ECC directly accessed and verified the underlying data reported in the manuscript. Formal analysis: DAJ. Funding acquisition: SC and ARM. Investigation: SC, ECC, DAJ, DT, DAB, GM, RKG, DGC, TCR and ARM. Supervision: ARM. Validation: RKG. Visualisation: DAJ. Writing of original draft: SC, ECC, DAJ and ARM. Writing – review and editing: SC, ECC, DAJ, DT, DAB, GM, RKG, DGC, TCR, ARM.

## Declaration of Interests

All authors have completed the ICMJE uniform disclosure form. TCR received salary support and travel cost reimbursement according to the terms of a service contract between FIND (a nonprofit organization) and his home institution, UC San Diego. TCR is a cofounder, board member, and unpaid shareholder of Verus Diagnostics Inc., a company that was founded with the intent of developing diagnostic assays. Verus Diagnostics was not involved in any way with data collection, analysis, or publication of the results of this manuscript. TCR has not received any financial support from Verus Diagnostics. No other author has had any financial relationship with any organisations that might have an interest in the submitted work in the previous three years. No other author has had any other relationship, or undertaken any activity, that could appear to have influenced the submitted work.

## Data Sharing

The study dataset is available from.

## Acknowledgements

We thank investigators from primary studies contributing data to this meta-analysis who provided us with re-formatted data. The study was funded by Barts Charity (ref G-002700). SC is supported by The Medical College of Saint Bartholomew’s Hospital Trust and a Wellcome HARP Doctoral Fellowship (ref 227532/Z/23/Z). ECC and DAJ are supported by Barts Charity (ref G-002700). ARM is supported by the United Kingdom Office for Students. GM is supported by the Wellcome Trust (214321/Z/18/Z, and 203135/Z/16/Z). The views expressed are those of the authors and not necessarily those of Barts Charity, The Medical College of Saint Bartholomew’s Hospital Trust, the Wellcome Trust or the Office for Students. For the purpose of open access, the authors have applied a CC BY public copyright licence to any Author Accepted Manuscript version arising from this submission.

## Transparency Declaration

SC and ARM are the manuscript’s guarantors, and they affirm that this is an honest, accurate, and transparent account of the study being reported and that no important aspects of the study have been omitted.

## Notes

### Funding Statement

The study was funded by Barts Charity (ref G-002700) The Medical College of Saint Bartholomews Hospital Trust and a Wellcome HARP Doctoral Fellowship (ref 227532/Z/23/Z). No funder had any role in writing the manuscript or in the decision to submit it for publication. The authors were not paid to write this article by a pharmaceutical company or other agency. The authors were not precluded from accessing data in the study, and they accept responsibility to submit for publication.

