## Supplementary Material for "Microbial biomarkers of tuberculosis infection and disease in blood: systematic review and meta-analysis"

### Supplementary Methods

#### Search terms

The following search terms were used to identify potentially eligible publications:

**PUBMED/MEDLINE**

("tuberculosis"[TIAB] OR "Mycobacterium tuberculosis"[TIAB] OR "M. tuberculosis"[TIAB] OR "Mtb"[TIAB] OR "tuberculostearic"[TIAB]) AND ("detection"[TIAB] OR "diagnosis"[TIAB] OR "diagnostic"[TIAB] OR "biomarker"[TIAB] OR "biomarkers"[TIAB] OR "diagnose"[TIAB] OR "identification"[TIAB]) AND ("blood"[TIAB] OR "plasma"[TIAB] OR "serum"[TIAB] OR "PBMC"[TIAB] OR "peripheral blood"[TIAB] OR "mononuclear cells"[TIAB] OR "whole blood"[TIAB] OR "CD34"[TIAB] OR "hematopoietic stem"[TIAB] OR "bacteremia"[TIAB] OR "bacteraemia"[TIAB] OR "bloodstream"[TIAB] OR "non-sputum"[TIAB] OR "circulating"[TIAB] OR "exosome"[TIAB] OR "extracellular vesicle"[TIAB]) AND ("antigen"[TIAB] OR "antigens"[TIAB] OR "DNA"[TIAB] OR "cell-free Mtb DNA"[TIAB] OR "cell-free"[TIAB] OR "LAM"[TIAB] OR "lipoarabinomannan"[TIAB] OR "ESAT-6"[TIAB] OR "CFP-10"[TIAB] OR "MPT64"[TIAB] OR "65 kD"[TIAB] OR "30 kD"[TIAB] OR "55 kDa"[TIAB] OR "55-kDa"[TIAB] OR "20 kDa"[TIAB] OR "IS6110"[TIAB] OR "rpoB"[TIAB] OR "mycothiol"[TIAB] OR "glycolipid"[TIAB] OR "mycobactin"[TIAB] OR "tuberculosinyladenosine"[TIAB] OR "MPT51"[TIAB] OR "MPT70"[TIAB] OR "TSTC"[TIAB] OR "Rv0819"[TIAB] OR "MAP-TB"[TIAB] OR "CRISPR"[TIAB] OR "Xpert"[TIAB] OR "GeneXpert"[TIAB] OR "Xpert Ultra"[TIAB] OR "bacteriophage"[TIAB] OR "phage"[TIAB] OR "Actiphage"[TIAB] OR "DMN-Tre"[TIAB] OR "trehalose"[TIAB] OR "modified lipoprotein"[TIAB] OR "lipoprotein"[TIAB] OR "TLP"[TIAB] OR (("exosome"[TIAB] OR "extracellular vesicle"[TIAB]) AND ("mass spectrometry”[TIAB] OR "MRM"[TIAB] OR "peptide"[TIAB])) OR "nucleic acid amplification"[TIAB] OR "NAAT"[TIAB]) AND (1990:2025[DP]) AND (Humans[Mesh])

**EMBASE**

("tuberculosis":ti,ab OR "Mycobacterium tuberculosis":ti,ab OR "M. tuberculosis":ti,ab OR "Mtb":ti,ab OR "tuberculostearic":ti,ab) AND ("detection":ti,ab OR "diagnosis":ti,ab OR "diagnostic":ti,ab OR "biomarker":ti,ab OR "biomarkers":ti,ab OR "diagnose":ti,ab OR "identification":ti,ab) AND ("blood":ti,ab OR "plasma":ti,ab OR "serum":ti,ab OR "PBMC":ti,ab OR "peripheral blood":ti,ab OR "mononuclear cells":ti,ab OR "whole blood":ti,ab OR "CD34":ti,ab OR "hematopoietic stem":ti,ab OR "bacteremia":ti,ab OR "bacteraemia":ti,ab OR "bloodstream":ti,ab OR "non-sputum":ti,ab OR "circulating":ti,ab OR "exosome":ti,ab OR "extracellular vesicle":ti,ab) AND ("antigen":ti,ab OR "antigens":ti,ab OR "DNA":ti,ab OR "cell-free Mtb DNA":ti,ab OR "cell-free":ti,ab OR "LAM":ti,ab OR "lipoarabinomannan":ti,ab OR "ESAT-6":ti,ab OR "CFP-10":ti,ab OR "MPT64":ti,ab OR "65 kD":ti,ab OR "30 kD":ti,ab OR "55 kDa":ti,ab OR "55-kDa":ti,ab OR "20 kDa":ti,ab OR "IS6110":ti,ab OR "rpoB":ti,ab OR "mycothiol":ti,ab OR "glycolipid":ti,ab OR "mycobactin":ti,ab OR "tuberculosinyladenosine":ti,ab OR "MPT51":ti,ab OR "MPT70":ti,ab OR "TSTC":ti,ab OR "Rv0819":ti,ab OR "MAP-TB":ti,ab OR "CRISPR":ti,ab OR "Xpert":ti,ab OR "GeneXpert":ti,ab OR "Xpert Ultra":ti,ab OR "bacteriophage":ti,ab OR "phage":ti,ab OR "Actiphage":ti,ab OR "DMN-Tre":ti,ab OR "trehalose":ti,ab OR "modified lipoprotein":ti,ab OR "lipoprotein":ti,ab OR "TLP":ti,ab OR (("exosome":ti,ab OR "extracellular vesicle":ti,ab) AND ("mass spectrometry”:ti,ab OR "MRM":ti,ab OR "peptide":ti,ab)) OR "nucleic acid amplification":ti,ab OR "NAAT":ti,ab) AND [humans]/lim AND [1990-2025]/py

**SCOPUS**

TITLE-ABS-KEY ( "tuberculosis" OR "Mycobacterium tuberculosis" OR "M. tuberculosis" OR "Mtb" OR "tuberculostearic" ) AND TITLE-ABS-KEY ( "detection" OR "diagnosis" OR "diagnostic" OR "biomarker" OR "biomarkers" OR "diagnose" OR "identification" ) AND TITLE-ABS-KEY ( "blood" OR "plasma" OR "serum" OR "PBMC" OR "peripheral blood" OR "mononuclear cells" OR "whole blood" OR "CD34" OR "hematopoietic stem" OR "bacteremia" OR "bacteraemia" OR "bloodstream" OR "non-sputum" OR "circulating" OR "exosome" OR "extracellular vesicle" ) AND TITLE-ABS-KEY ( "antigen" OR "antigens" OR "DNA" OR "cell-free Mtb DNA" OR "cell-free" OR "LAM" OR "lipoarabinomannan" OR "ESAT-6" OR "CFP-10" OR "MPT64" OR "65 kD" OR "30 kD" OR "55 kDa" OR "55-kDa" OR "20 kDa" OR "IS6110" OR "rpoB" OR "mycothiol" OR "glycolipid" OR "mycobactin" OR "tuberculosinyladenosine" OR "MPT51" OR "MPT70" OR "TSTC" OR "Rv0819" OR "MAP-TB" OR "CRISPR" OR "Xpert" OR "GeneXpert" OR "Xpert Ultra" OR "bacteriophage" OR "phage" OR "Actiphage" OR "DMN-Tre" OR "trehalose" OR "modified lipoprotein" OR "lipoprotein" OR "TLP" OR ( ( "exosome" OR "extracellular vesicle" ) AND ( "mass spectrometry" OR "MRM" OR "peptide" ) ) OR "nucleic acid amplification" OR "NAAT" ) AND PUBYEAR > 1989 AND PUBYEAR < 2026 AND TITLE-ABS-KEY ( "humans" ).

#### Assessments of risk of bias and applicability

Risk of bias and applicability were assessed using modified QUADAS-3 criteria^1^ as detailed below. Answers to signalling questions about risk of bias across four domains (participants, index test, target condition and analysis) were classified as ‘Yes’, ‘Probably Yes’, ‘Probably No’, ‘No’ or ‘No Information’. The risk of bias for each domain was judged as low if responses to all signalling questions were ‘Yes’ or ‘Probably Yes’; as high if responses to any signalling question were ‘No’ or ‘Probably No’; and as ‘insufficient information’ if insufficient data were available for the reviewer to make a judgement. A global judgement about each study was then made: if any of the four domains was assessed as being at high risk of bias then the study was classified as being at high risk of bias overall; all other studies were classified as low risk of bias overall.

Answers to signalling questions about applicability across three domains (participants, index test and target condition) were classified as causing ‘Low Concern’, ‘High Concern’ or ‘Insufficient Information’. A global judgement about each study was then made: if any of the three domains was assessed as giving cause for high concern then the study was classified as giving cause for high concern re applicability overall; ; all other studies were classified as giving cause for low risk of concern re applicability overall.

**DOMAIN 1: PARTICIPANTS**

**A. Risk of bias:**

1.1	Was a single-gate study design used?

1.2	Were participants enrolled prospectively?

1.3	Were participants enrolled consecutively or randomly?

1.4	Did the actual population of enrolled participants match the target population that the study aimed to recruit?

**B. Applicability concerns:**

Is there concern that the participant population in the study does not match the intended population for a tuberculosis detection biomarker?

**DOMAIN 2: INDEX TEST**

**A. Risk of bias:**

2.1	Was the index test performed in the same way on all participants?

2.2	If a threshold was used for the index test, was the method used to derive the cutoff standard or pre-specified?

2.3	Were the index test results interpreted without knowledge of the reference standard results (i.e. blinded)?

**B. Applicability concerns:**

Is there concern that the type, use or interpretation of the index test in the study does not match the intended type, use or interpretation of a tuberculosis detection biomarker?

**DOMAIN 3: TARGET CONDITION**

**A. Risk of bias:**

3.1	Was an appropriate reference standard used to correctly identify individuals with or without the target condition specified by the study?

3.2	Was the target condition assessed in all participants?

3.3	Was the target condition assessed in the same way in all individuals?

3.4	Did the reference standard avoid incorporating the index test?

3.5	Was there an appropriate time interval between index testing and reference standard assessment?

**B. Applicability concerns:**

Is there concern that the target condition in the study does not match the intended target condition for a tuberculosis detection biomarker?

**DOMAIN 4: ANALYSIS**

**Risk of bias:**

4.1 Were all participants included in the analysis?
4.2 Were missing data handled appropriately?

### Supplementary Results

### Table S1: Excluded studies (n=30)

| **Study** | **Target type** | **Reason for exclusion** |
| --- | --- | --- |
| Adhikhary_2025^2^ | Antigen | Unsuitable reporting of outcome |
| Aziz_2004^3^ | Cell-associated DNA | Unsuitable reporting of outcome |
| Cho_2011^4^ | Cell-associated DNA | Unsuitable reporting of outcome |
| Dai_2023 ^5^ | Antigen | Unsuitable reporting of outcome |
| de_Lyra_2014 ^6^ | Cell-associated DNA | Unsuitable reporting of outcome |
| Elsohaby_2020 ^7^ | Cell-associated DNA | Unsuitable reporting of outcome |
| Hanif_2012 ^8^ | Cell-associated DNA | Unsuitable reporting of outcome |
| Jiang_2025 ^9^ | Antigen | Unsuitable reporting of outcome |
| Kansal_1992 ^10^ | Antigen | Unsuitable reporting of outcome |
| Kox_1994 ^11^ | Cell-associated DNA | Cases series <3 eligible participants |
| Li_2022 ^12^ | Antigen | Unsuitable reporting of outcome |
| Li_2025 ^13^ | Cell-associated DNA | Unsuitable reporting of outcome |
| Majumdar_2010 ^14^ | Antigen | Detection of host response |
| Mao_2020 ^15^ | Antigen | Duplication |
| Montenegro_2013 ^16^ | Cell-associated DNA | Unsuitable reporting of outcome |
| Mourembou_2015 ^17^ | Cell-associated DNA | Does not report on Mtb detection |
| Nair_2000 ^18^ | Antigen | Detection of host response |
| Nandagopal_2010 ^19^ | Cell-associated DNA | Unsuitable reporting of outcome |
| Raza_2009 ^20^ | Cell-associated DNA | Unsuitable reporting of outcome |
| Rebollo_2006 ^21^ | Cell-associated DNA | Unsuitable reporting of outcome |
| Ritis_2000 ^22^ | Cell-associated DNA | Cases series <3 eligible participants |
| Rossetti_1997 ^23^ | Cell-associated DNA | Unsuitable reporting of outcome |
| Salazar_2024 ^24^ | Cell-associated DNA | Unsuitable reporting of outcome |
| Santos_2018 ^25^ | Cell-associated DNA | Unsuitable reporting of outcome |
| Shende_2005 ^26^ | Antigen | Unsuitable reporting of outcome |
| Shende_2008 ^27^ | Antibody | Detection of host response only |
| Stavri_1990b^28^ | Antigen | Duplication |
| Udaykumar_1991^29^ | Antigen | Unsuitable reporting of outcome |
| Waghmare_2012^30^ | Antigen | Unsuitable reporting of outcome |
| Wang_2025^31^ | Cell-free Mtb DNA | Assay development |

### Table S2: Assessments of risk of bias and applicability

| **First Author, Year** | **Analyte Class** | **Risk of Bias** | | | | **Applicability Concerns** | | |
| --- | --- | --- | --- | --- | --- | --- | --- | --- |
|  |  | Participants | Index Test | Target Condition | Analysis | Participants | Index Test | Target Condition |
| Sada, 1990 | Unspecified or combined antigen | - | - | + | + | - | + | + |
| Stravi, 1990 | Unspecified or combined antigen | - | - | + | + | - | + | + |
| Sood, 1991 | Protein | - | - | - | + | - | + | + |
| Ashtekar, 1992 | Unspecified or combined antigen | - | - | ? | - | - | + | + |
| Kolk, 1992 | Cell-associated Mtb DNA | - | + | - | + | - | + | + |
| Radhakrishnan, 1992 | Protein | - | - | - | + | - | + | - |
| Sada, 1992 | Glycolipid | - | - | ? | + | - | + | + |
| Rattan, 1993 | Protein | + | - | - | - | + | + | + |
| Schluger, 1994 | Cell-associated Mtb DNA / Cell-free Mtb DNA | - | ? | - | + | - | - | + |
| Patil, 1995 | Glycolipid | - | - | - | + | - | - | + |
| Rolfs, 1995 | Cell-associated Mtb DNA | + | + | + | + | - | - | + |
| Condos, 1996 | Cell-associated Mtb DNA | + | + | + | + | + | + | + |
| Folgueira, 1996 | Cell-associated Mtb DNA | - | ? | - | + | - | + | + |
| Khomenko, 1996 | Glycolipid | - | - | - | - | - | + | + |
| Richter, 1996 | Cell-associated Mtb DNA | + | + | + | + | + | + | + |
| Del Prete, 1997 | Cell-associated Mtb DNA | - | + | + | + | - | + | + |
| Ahmed, 1998 | Cell-associated Mtb DNA | - | ? | - | + | - | + | + |
| Brooks, 1998 | Lipid | - | - | ? | - | - | - | + |
| Gupta, 1998 | Unspecified or combined antigen | - | - | + | - | - | + | + |
| Lodam, 1998 | Unspecified or combined antigen | - | - | - | + | - | + | + |
| Seethalakshmi, 1998 | Cell-associated Mtb DNA | ? | ? | - | - | + | + | + |
| Sethna, 1998 | Protein | - | - | + | + | - | + | + |
| van Staden, 1998 | Cell-associated Mtb DNA | - | ? | - | + | - | - | + |
| Srivastava, 1999 | Unspecified or combined antigen | - | - | + | + | - | + | + |
| Almeda, 2000 | Cell-associated Mtb DNA | - | + | + | + | - | + | + |
| Chanteau, 2000 | Protein | - | - | + | - | - | + | + |
| Chaturvedi, 2001 | Unspecified or combined antigen | - | - | + | + | - | + | + |
| Honore, 2001 | Cell-associated Mtb DNA | - | ? | - | + | - | + | + |
| Landowski, 2001 | Protein | - | - | - | + | - | + | + |
| Attallah, 2003 | Protein | - | + | - | + | - | + | + |
| Mirza, 2003 | Cell-associated Mtb DNA | - | ? | - | + | - | + | + |
| Taci, 2003 | Cell-associated Mtb DNA | - | ? | - | + | - | + | + |
| Verettas, 2003 | Cell-associated Mtb DNA | - | - | + | + | - | + | + |
| Attallah, 2005 | Protein | - | + | - | + | - | + | + |
| Bera, 2006 | Protein | - | ? | - | + | - | + | + |
| Harinath, 2006 | Protein | - | - | - | + | - | + | + |
| Katti, 2006 | Unspecified or combined antigen | - | - | ? | + | - | + | + |
| Khan, 2006 | Cell-associated Mtb DNA | ? | ? | + | + | + | + | + |
| Kashyap, 2007 | Protein | - | - | + | + | - | + | + |
| Rajan, 2007 | Protein | - | - | - | + | - | + | + |
| Sharafeldin, 2007 | Cell-associated Mtb DNA | + | ? | - | - | + | + | + |
| Tiwari, 2007 | Glycolipid | - | + | + | + | - | + | + |
| Upadhye, 2007 | Protein | - | + | + | + | - | + | + |
| El-Masry, 2008 | Protein | - | - | - | + | - | + | + |
| Majumdar, 2008 | Protein | - | - | + | + | - | + | + |
| Shende, 2008 | Protein | - | ? | + | + | - | + | + |
| da Costa Lima, 2009 | Cell-associated Mtb DNA | - | + | + | + | - | + | + |
| Hira, 2009 | Cell-associated Mtb DNA | - | ? | - | + | - | + | + |
| Zhu, 2009 | Cell-associated Mtb DNA | - | ? | + | + | - | + | + |
| Kumar, 2010 | Cell-associated Mtb DNA | ? | ? | + | + | + | + | + |
| Crump, 2012 | Cell-associated Mtb DNA | ? | + | + | + | + | + | + |
| Mukundan 2012 | Protein | - | - | + | + | - | + | + |
| Zhu, 2012 | Protein | - | - | - | + | - | + | + |
| Dubey, 2013 | Cell-associated Mtb DNA | - | ? | + | + | - | + | + |
| Feasey, 2013 | Cell-associated Mtb DNA | + | + | - | - | + | + | + |
| Kashyap, 2013 | Cell-associated Mtb DNA / protein | - | ? | - | + | - | + | + |
| Nath, 2013 | Unspecified or combined antigen | - | ? | + | + | + | + | + |
| Pan, 2013 | Cell-free Mtb DNA | - | ? | - | + | - | + | + |
| Sakamuri, 2013 | Glycolipid | - | - | + | + | - | + | + |
| Shenai, 2013 | Cell-associated Mtb DNA | - | + | + | + | - | + | + |
| Frediani, 2014 | Lipid | - | - | - | + | - | - | + |
| Hajiabdolbaghi, 2014 | Cell-associated Mtb DNA | - | ? | - | + | - | + | + |
| Kruh-Garcia, 2014 | Peptide | - | - | + | + | - | - | - |
| Li, 2014 | Protein | - | - | - | + | - | + | + |
| Sankar, 2014 | Cell-associated Mtb DNA | + | ? | - | + | + | + | - |
| Tang, 2014 | Protein | - | - | + | + | - | + | + |
| Bwanga, 2015 | Cell-associated Mtb DNA | ? | - | + | + | + | + | + |
| Chan, 2015 | Glycolipid | - | - | + | - | - | + | + |
| da Costa-Lima, 2015 | Cell-associated Mtb DNA / Cell-free Mtb DNA | - | + | + | - | - | + | + |
| Pan, 2015 | Lipid | - | - | + | + | - | - | + |
| Pohl, 2016 | Cell-associated Mtb DNA | + | + | + | + | + | + | + |
| Poulakis, 2016 | Protein | - | - | - | + | - | - | + |
| Tang, 2016 | Glycolipid | - | - | + | + | - | + | + |
| Ushio, 2016 | Cell-free Mtb DNA | - | ? | - | + | - | + | + |
| Bai, 2017 | Protein | - | - | - | + | - | - | + |
| Castro-Garza, 2017 | Protein | - | - | - | + | - | + | + |
| Crawford, 2017 | Glycolipid | - | - | ? | + | - | - | + |
| Fan, 2017 | Protein | - | - | - | - | + | - | + |
| Goyal, 2017 | Protein | - | - | - | + | - | + | + |
| Liu, 2017 | Peptide | - | - | + | + | - | - | + |
| Manke, 2017 | Cell-associated Mtb DNA | - | ? | - | - | + | + | + |
| Mehaffy, 2017 | Peptide | - | - | ? | + | - | - | + |
| Tiwari, 2017 | Protein | - | - | + | + | - | + | + |
| Tornack 2017 | Cell-associated Mtb DNA | - | - | - | + | - | - | - |
| Wu, 2017 | Protein | - | ? | - | + | - | + | + |
| Yang, 2017 | Cell-associated Mtb DNA | - | - | - | + | - | + | + |
| Zeitoun, 2017 | Protein | - | - | + | + | - | + | + |
| Amin, 2018 | Glycolipid | - | - | + | - | - | - | + |
| Click, 2018 | Cell-free Mtb DNA | - | ? | + | + | - | + | + |
| Liu, 2018 | Peptide | + | + | + | - | + | - | + |
| Broger, 2019 | Glycolipid/protein | - | + | + | + | - | - | + |
| Verma, 2019 | Cell-associated Mtb DNA | - | + | - | + | - | + | + |
| Brock, 2020 | Glycolipid | - | - | + | + | + | - | + |
| da Costa Lima, 2020 | Cell-associated Mtb DNA / Cell-free Mtb DNA | ? | + | + | + | + | + | + |
| Lyu, 2020 | Cell-free Mtb DNA | - | - | - | + | - | + | + |
| Mehaffy, 2020 | Peptide | - | - | + | + | - | - | - |
| Belay, 2021 | Cell-associated Mtb DNA | ? | ? | - | - | + | - | + |
| Gurmessa, 2021 | Protein | - | - | - | ? | - | + | - |
| He, 2021 | Peptide | - | + | + | - | - | - | + |
| Jakhar, 2021 | Glycolipid | - | - | + | - | - | + | + |
| Mao, 2021 | Peptide | - | + | - | - | + | - | + |
| Pan, 2021 | Cell-free Mtb DNA | - | - | + | + | - | + | + |
| Pollock, 2021 | Cell-free Mtb DNA | - | ? | - | + | - | - | + |
| Sam, 2021 | Cell-free Mtb DNA | - | - | - | - | - | + | + |
| Barr, 2022 | Cell-associated Mtb DNA | - | + | + | + | - | + | + |
| Boloko, 2022 | Cell-associated Mtb DNA | + | + | + | + | + | + | + |
| Brandenburg, 2022 | Lipid | - | - | - | + | - | - | + |
| Chang, 2022 | Cell-free Mtb DNA | - | - | - | + | - | - | + |
| Chen, 2022 | Glycoprotein | - | - | - | + | + | + | + |
| Gaballah, 2022 | Cell-free Mtb DNA | ? | ? | - | - | + | + | + |
| Huang, 2022 | Cell-free Mtb DNA | - | - | - | + | - | - | + |
| Park, 2022 | Cell-free Mtb DNA | + | ? | + | + | - | + | + |
| Yang, 2022 | Protein | - | - | - | + | - | + | - |
| Zheng, 2022 | Unspecified or combined antigen | + | + | + | + | + | + | + |
| Araújo, 2023 | Cell-associated Mtb DNA / Cell-free Mtb DNA | - | ? | + | + | - | + | + |
| Kil, 2023 | Protein | - | - | + | + | - | - | - |
| Li, 2023 | Protein | - | + | + | + | - | - | + |
| Sharma, 2023 | Cell-free Mtb DNA | - | - | + | + | - | + | + |
| Thakku, 2023 | Cell-free Mtb DNA | - | + | - | + | - | - | + |
| Wang, 2023 | Protein | ? | - | + | + | + | + | + |
| Yarmohammadi, 2023 | Cell-associated Mtb DNA | - | - | + | + | - | + | + |
| Chen, 2024 | Cell-free Mtb DNA | - | - | - | + | - | - | + |
| Drain, 2024 | Glycolipid | + | + | + | + | + | + | + |
| Kim, 2024 | Cell-associated Mtb DNA | - | + | + | + | + | + | + |
| Li, 2024 | Cell-free Mtb DNA | - | - | - | - | - | - | + |
| Ma, 2024 | Cell-free Mtb DNA | - | ? | - | + | - | - | + |
| Repele, 2024 | Cell-associated Mtb DNA | - | ? | - | + | - | - | + |
| Sheng, 2024 | Protein | - | - | + | - | - | + | + |
| Shield, 2024 | Cell-free Mtb DNA | - | ? | - | + | - | + | - |
| Ayalew (a), 2025 | Cell-free Mtb DNA | - | ? | - | + | - | + | + |
| Ayalew (b), 2025 | Cell-free Mtb DNA | - | ? | - | + | - | + | + |
| Boloko, 2025 | Cell-associated Mtb DNA | - | + | + | + | - | + | + |
| Li, 2025 | Peptide | + | + | + | + | + | - | + |
| Liu, 2025 | Protein | - | - | + | + | - | - | + |
| Quan, 2025 | Cell-free Mtb DNA | - | ? | + | + | - | - | + |
| Ye, 2025 | Protein | - | - | + | + | - | + | + |
| Youngquist, 2025 | Cell-free Mtb DNA | - | + | - | + | - | + | + |

| Low Risk (+) | Unclear (?) | High Risk (-) |
| --- | --- | --- |

### Table S3: Bivariate analyses estimating sensitivity and specificity of different microbial biomarker classes for the diagnosis of active tuberculosis: overall, and sensitivity analyses excluding studies assessed as being at high risk of bias

| **Target** | **N** | **Sensitivity**  **(95% CI)** | **Variation in sensitivity attributable to between-study heterogeneity,**  ***I^2^* (95% CI)** | **Specificity (95% CI)** | **Variation in specificity attributable to between-study heterogeneity,**  ***I^2^* (95% CI)** | **AUC**  **(95% CI)** | **Proportion of between-study heterogeneity due to threshold effect** |
| --- | --- | --- | --- | --- | --- | --- | --- |
| **Overall** | | | | | | | |
| Cell-free Mtb DNA | 34 | 61.5 (51.0, 71.0) | 30% (18, 42) | 93.0 (88.1, 96.1) | 43% (28, 58) | 0.87 (0.84, 0.89) | 34% |
| Cell-associated Mtb DNA | 32 | 43.9 (29.4, 59.4) | 47% (31, 63) | 97.1 (94.5, 98.5) | 36% (16, 56) | 0.93 (0.90, 0.95) | 5% |
| Protein/peptide/glycoprotein antigens | 61 | 78.9 (73.2, 83.6) | 29% (20, 38) | 92.9 (90.7, 94.5) | 21% (13, 29) | 0.94 (0.92, 0.96) | 2% |
| Lipid/glycolipid antigens | 22 | 68.6 (54.1, 80.3) | 37% (21, 54) | 97.0 (94.0, 98.5) | 32% (9, 54) | 0.96 (0.94, 0.97) | 8% |
| Unspecified or combined antigens | 10 | 80.2 (67.1, 88.9) | 24% (5, 42) | 87.9 (76.0, 94.3) | 28% (3, 53) | 0.91, 0.88, 0.93) | 19% |
| **Sensitivity analysis excluding studies at high risk of bias** | | | | | | | |
| Cell-free Mtb DNA | --* | --* |  | --* |  | --* | --* |
| Cell-associated Mtb DNA | 9 | 31.6 (16.0, 52.8) | 34% (10, 58) | 96.7 (94.1, 98.2) | 10% (0, 24) | 0.94 (0.91, 0.96) | 100% |
| Protein/peptide/glycoprotein antigens | --* | --* |  | --* |  | --* | --* |
| Lipid/glycolipid antigens | --* | --* |  | --* |  | --* | --* |
| Unspecified or combined antigens | --* | --* |  | --* |  | --* | --* |

--* insufficient studies (<4) to perform sensitivity analysis after excluding studies at high risk of bias

### Table S4: Meta-regression of Cell-free DNA studies

| **Factor^1^** | **N** | **Sensitivity**  **(95% CI)** | **Variation in sensitivity attributable to between-study heterogeneity,**  ***I^2^* (95% CI)** | **Specificity (95% CI)** | **Variation in specificity attributable to between-study heterogeneity,**  ***I^2^* (95% CI)** | **AUC**  **(95% CI)** | **Proportion of between-study heterogeneity due to threshold effect** | **Pairwise**  **P value** | **Global P value** |
| --- | --- | --- | --- | --- | --- | --- | --- | --- | --- |
| Overall | 34 | 61.5 (51.0, 71.0) | 30% (18, 42) | 93.0 (88.1, 96.1) | 43% (28, 58) | 0.87 (0.84, 0.89) | 34% | - | - |
| Age |  |  |  |  |  |  |  |  |  |
| Adults | 23 | 58.8 (48.6, 68.3) | 21% (9, 33) | 92.6 (86.6, 96.0) | 38% (21, 55) | 0.84 (0.80, 0.87) | 61% | Ref. **^2^** | - |
| Children | 4 | 62.4 (33.6, 84.5) | 29% (0, 60) | 90.8 (64.3, 98.2) | 43% (4, 81) | 0.85 (0.81, 0.88) | 85% | 0.96 |  |
| WHO Region |  |  |  |  |  |  |  |  |  |
| WPRO | 14 | 48.2 (37.9, 58.6) | 15% (4, 25) | 93.1 (89.0, 95.7) | 11% (0, 28) | 0.86 (0.83, 0.89) | 23% | Ref. | 0.039 |
| AFRO | 13 | 70.4 (57.3, 80.9) | 22% (6, 38) | 91.3 (84.0, 95.5) | 27% (9, 46) | 0.89 (0.86, 0.92) | 75% | 0.033 |  |
| AMRO | 6 | 78.7 (49.3, 93.3) | 41% (8, 73) | 95.1 (46.6, 99.8) | 80% (57, 100) | 0.91 (0.88, 0.93) | 55% | 0.34 |  |
| Study Design |  |  |  |  |  |  |  |  |  |
| Multi-gate | 30 | 62.0 (50.2, 72.4) | 32% (19, 46) | 93.1 (87.4, 96.3) | 46% (30, 62) | 0.87 (0.84, 0.90) | 34% | Ref. | - |
| Single-gate | 4 | 59.1 (41.4, 74.7) | 11% (0, 29) | 90.5 (84.3, 94.4) | 1% (0, 8) | 0.90 (0.87, 0.93) | 100% | 0.99 |  |
| Target |  |  |  |  |  |  |  |  |  |
| Multi-copy | 24 | 66.0 (54.6, 75.8) | 26% (12, 41) | 90.8 (86.4, 94.0) | 21% (8, 33) | 0.89 (0.86, 0.92) | 34% | Ref. | - |
| Single-copy | 10 | 49.9 (30.3, 69.5) | 34% (12, 56) | 96.8 (82.5, 99.5) | 69% (46, 93) | 0.81 (0.77, 0.84) | 37% | 0.32 |  |
| Detection Method |  |  |  |  |  |  |  |  |  |
| qPCR | 15 | 53.6 (42.8, 64.1) | 16% (4, 29) | 94.3 (91.5, 96.2) | 8% (0, 20) | 0.92 (0.89, 0.94) | 23% | Ref. | 0.21 |
| dPCR / ddPCR | 5 **^3^** |  |  |  |  |  |  |  |  |
| CRISPR-driven | 5 | 84.2 (58.5, 95.3) | 34% (0, 94) | 83.2 (61.1, 94.0) | 29% (0, 60) | 0.90 (0.88, 0.93) | 1% | 0.064 |  |
| Whole genome / metagenomic sequencing | 5 | 65.8 (35.3, 87.1) | 37% (5, 68) | 96.8 (46.2, 99.9) | 81% (60, 100) | 0.85 (0.82, 0.88) | 86% | 0.64 |  |
| Sample Type |  |  |  |  |  |  |  |  |  |
| Plasma | 29 | 54.6 (44.6, 64.2) | 25% (13, 36) | 93.6 (88.5, 96.5) | 43% (27, 59) | 0.83 (0.79, 0.86) | 41% |  | - |
| Serum | 5 **^3^** |  |  |  |  |  |  |  |  |

**(1)** Where the sum of studies in the subgroups is less than the overall number of studies, one or more subgroups were excluded because they did not meet the minimum requirement of 4 studies for bivariate modelling.
**(2)** Ref. indicates the reference category (most studies) used for pairwise Wald tests. Global P values indicate the significance of the factor across all subgroups using a joint Wald test; pairwise P values compare the specific subgroup against the reference.

**(3)** Estimates not reported due to failure of model to converge.

**Abbreviations:** **AFRO:** WHO African Region; **AMRO:** WHO Region of the Americas; **AUC:** Area Under the Curve; **CI:** Confidence Interval; **CRISPR:** Clustered Regularly Interspaced Short Palindromic Repeats; **ddPCR:** Droplet Digital Polymerase Chain Reaction; **dPCR:** Digital Polymerase Chain Reaction; **qPCR:**Quantitative Polymerase Chain Reaction; **WPRO:** WHO Western Pacific Region.

### Table S5: Meta-regression of Cell-Associated DNA studies

| **Factor ^1^** | **N** | **Sensitivity**  **(95% CI)** | **Variation in sensitivity attributable to between-study heterogeneity,**  ***I^2^* (95% CI)** | **Specificity (95% CI)** | **Variation in specificity attributable to between-study heterogeneity,**  ***I^2^* (95% CI)** | **AUC**  **(95% CI)** | **Proportion of between-study heterogeneity due to threshold effect** | **Pairwise**  **P value** | **Global P value** |
| --- | --- | --- | --- | --- | --- | --- | --- | --- | --- |
| Overall | 32 | 43.9 (29.4, 59.4) | 47% (31, 63) | 97.1 (94.5, 98.5) | 36% (16, 56) | 0.93 (0.90, 0.95) | 5% | - | - |
| Decade of publication |  |  |  |  |  |  |  |  |  |
| 1990s | 11 **^3^** |  |  |  |  |  |  |  | 0.32 |
| 2000s | 7 | 46.4 (27.3, 66.6) | 24% (0, 49) | 98.0 (80.2, 99.8) | 48% (0, 100) | 0.86 (0.82, 0.88) | 100% | 0.60 |  |
| 2010s | 8 | 42.8 (15.7, 75.0) | 53% (22, 84) | 98.8 (94.4, 99.8) | 49% (11, 87) | 0.96 (0.93, 0.97) | 2% | Ref. **^2^** |  |
| 2020s | 6 | 39.6 (6.7, 85.6) | 67% (30, 100) | 91.3 (80.0, 96.5) | 26% (0, 55) | 0.88 (0.85, 0.90) | 72% | 0.11 |  |
| WHO Region |  |  |  |  |  |  |  |  |  |
| EURO | 13 | 35.9 (21.6, 53.2) | 31% (10, 52) | 94.7 (92.0, 96.5) | 3% (0, 13) | 0.93 (0.91, 0.95) | 85% | Ref. | 0.12 |
| AFRO | 4 | 16.1 (8.3, 28.8) | 12% (0, 33) | 99.3 (97.1, 99.8) | 1% (0, 11) | 0.99 (0.98, 1.00) | 100% | 0.053 |  |
| AMRO | 5 | 58.4 (4.0, 98.0) | 81% (51, 100) | 93.6 (65.5, 99.1) | 53% (3, 100) | 0.93 (0.90, 0.95) | 73% | 0.40 |  |
| EMRO | 4 **^3^** |  |  |  |  |  |  |  |  |
| SEARO | 4 | 38.8 (26.1, 53.2) | 7% (0, 19) | 99.2 (70.0, 99.9) | 74% (29, 100) | 0.58 (0.54, 0.62) | 100% | 0.71 |  |
| Study Design |  |  |  |  |  |  |  |  |  |
| Multi-gate | 19 | 54.5 (30.7, 76.4) | 56% (34, 79) | 96.0 (90.6, 98.4) | 36% (08, 65) | 0.94 (0.91, 0.96) | 0% | Ref. | - |
| Single-gate | 13 | 31.8 (18.8, 48.5) | 32% (13, 51) | 97.9 (95.5, 99.0) | 22% (0, 46) | 0.93 (0.90, 0.95) | 56% | 0.11 |  |
| Target |  |  |  |  |  |  |  |  |  |
| Multi-copy | 27 | 45.8 (29.6, 62.9) | 49% (31, 66) | 97.5 (94.4, 98.9) | 44% (20, 67) | 0.93 (0.90, 0.95) | 7% | Ref. | - |
| Single-copy | 5 | 31.6 (12.4, 60.2) | 30% (0, 74) | 96.7 (92.2, 98.7) | 9% (0, 34) | 0.94 (0.91, 0.96) | 100% | 0.79 |  |
| Detection Method |  |  |  |  |  |  |  |  |  |
| Endpoint PCR | 21 | 44.2 (33.1, 55.9) | 24% (11, 38) | 97.3 (94.2, 98.8) | 33% (7, 59) | 0.88 (0.85, 0.91) | 1% | - | - |
| dPCR / ddPCR | 4 **^3^** |  |  |  |  |  |  |  |  |
| Sample Type |  |  |  |  |  |  |  |  |  |
| PBMC | 14 | 46.1 (22.9, 71.2) | 52% (26, 78) | 96.6 (90.2, 98.9) | 38% (3, 73) | 0.93 (0.90, 0.95) | 3% | Ref. | - |
| Whole blood | 14 | 47.0 (24.5, 70.8) | 50% (27, 74) | 96.5 (93.7, 98.0) | 14% (0, 31) | 0.95 (0.93, 0.97) | 8% | 0.97 |  |
| Buffy coat / undifferentiated leukocytes | 4 **^3^** |  |  |  |  |  |  |  |  |

**(1)** Where the sum of studies in the subgroups is less than the overall number of studies, one or more subgroups were excluded because they did not meet the minimum requirement of 4 studies for bivariate modelling.
**(2)** Ref. indicates the reference category (most studies) used for pairwise Wald tests. Global P values indicate the significance of the factor across all subgroups using a joint Wald test; pairwise P values compare the specific subgroup against the reference.

**(3)** Estimates not reported due to failure of model to converge.

**Abbreviations:** **AFRO:** WHO African Region; **AMRO:** WHO Region of the Americas; **AUC:** Area Under the Curve; **CI:** Confidence Interval; **ddPCR:** Droplet Digital Polymerase Chain Reaction; **dPCR:** Digital Polymerase Chain Reaction; **EMRO:** WHO Eastern Mediterranean Region; **EURO:** WHO European Region; **PBMC:** Peripheral Blood Mononuclear Cells; **SEARO:** WHO South-East Asian Region.

### Table S6: Meta-regression of Protein/Peptide/Glycoprotein Studies

| **Factor ^1^** | **N** | **Sensitivity**  **(95% CI)** | **Variation in sensitivity attributable to between-study heterogeneity,**  ***I^2^* (95% CI)** | **Specificity (95% CI)** | **Variation in specificity attributable to between-study heterogeneity,**  ***I^2^* (95% CI)** | **AUC**  **(95% CI)** | **Proportion of between-study heterogeneity due to threshold effect** | **Pairwise**  **P value** | **Global P value** |
| --- | --- | --- | --- | --- | --- | --- | --- | --- | --- |
| Overall | 61 | 78.9 (73.2, 83.6) | 29% (20, 38) | 92.9 (90.7, 94.5) | 21% (13, 29) | 0.94 (0.92, 0.96) | 2% | - | - |
| Age |  |  |  |  |  |  |  |  |  |
| Adults | 15 | 72.2 (57.0, 83.6) | 33% (13, 53) | 92.0 (84.7, 95.9) | 33% (11, 55) | 0.91 (0.88, 0.93) | 3% | Ref. **^2^** | - |
| Children | 6 | 82.2 (77.6, 86.1) | 0% (0, 1) | 94.5 (87.1, 97.7) | 20% (0, 53) | 0.85 (0.82, 0.88) | 100% | 0.35 |  |
| Decade of publication |  |  |  |  |  |  |  |  |  |
| 1990s | 5 | 54.0 (33.9, 73.0) | 20% (0, 41) | 88.5 (76.3, 94.8) | 18% (0, 41) | 0.82 (0.78, 0.85) | 53% | 0.075 | 0.17 |
| 2000s | 22 | 77.9 (69.4, 84.7) | 24% (12, 36) | 93.9 (91.9, 95.4) | 5% (0, 11) | 0.95 (0.93, 0.97) | 0% | Ref. |  |
| 2010s | 19 | 83.1 (69.6, 91.4) | 43% (22, 65) | 92.0 (85.2, 95.8) | 36% (17, 55) | 0.95 (0.92, 0.96) | 1% | 0.27 |  |
| 2020s | 15 | 81.4 (75.1, 86.5) | 10% (0, 21) | 93.2 (89.0, 95.9) | 14% (0, 32) | 0.94 (0.92, 0.96) | 89% | 0.76 |  |
| WHO Region |  |  |  |  |  |  |  |  |  |
| SEARO | 21 | 79.6 (71.1, 86.1) | 24% (12, 36) | 92.7 (89.2, 95.1) | 18% (7, 29) | 0.94 (0.92, 0.96) | 29% | Ref. | 0.36 |
| AFRO | 8 | 71.6 (47.3, 87.7) | 38% (12, 64) | 89.9 (73.2, 96.7) | 40% (10, 70) | 0.89 (0.86, 0.92) | 60% | 0.29 |  |
| AMRO | 9 | 66.6 (42.5, 84.3) | 39% (10, 68) | 93.7 (88.1, 96.7) | 18% (0, 39) | 0.93 (0.91, 0.95) | 36% | 0.33 |  |
| EMRO | 6 | 81.7 (70.4, 89.3) | 15% (0, 30) | 93.6 (90.0, 95.9) | 2% (0, 11) | 0.95 (0.93, 0.97) | 14% | 0.89 |  |
| WPRO | 15 | 85.8 (76.6, 91.8) | 24% (4, 44) | 93.1 (90.5, 95.1) | 5% (0, 12) | 0.96 (0.93, 0.97) | 38% | 0.53 |  |
| Study Design |  |  |  |  |  |  |  |  |  |
| Multi-gate | 50 | 79.4 (72.6, 84.9) | 33% (22, 44) | 93.1 (90.6, 95.0) | 24% (14, 34) | 0.95 (0.92, 0.96) | 2% | Ref. | - |
| Single-gate | 11 | 77.4 (70.0, 83.7) | 10% (0, 19) | 91.9 (88.0, 94.6) | 06% (0, 17) | 0.93 (0.91, 0.95) | 14% | 0.95 |  |
| Detection Method |  |  |  |  |  |  |  |  |  |
| ELISA | 34 | 71.3 (62.6, 78.7) | 28% (16, 39) | 91.7 (89.0, 93.8) | 14% (6, 22) | 0.92 (0.90, 0.94) | 20% | Ref. | - |
| Mass spectrometry | 7 | 85.6 (81.4, 88.9) | 1% (0, 3) | 93.9 (73.4, 98.8) | 55% (20, 91) | 0.87 (0.84, 0.90) | 100% | 0.055 |  |
| Nanoparticle mediated detection | 4 **^3^** |  |  |  |  |  |  |  |  |
| Sample Type |  |  |  |  |  |  |  |  |  |
| Serum | 54 | 76.9 (70.7, 82.2) | 29% (20, 38) | 92.7 (90.5, 94.4) | 19% (11, 28) | 0.94 (0.91, 0.96) | 2% | Ref. | - |
| Plasma | 6 | 90.8 (79.7, 96.1) | 15% (0, 45) | 96.7 (90.3, 98.9) | 10% (0, 45) | 0.98 (0.97, 0.99) | 100% | 0.045 |  |
| Antigen association |  |  |  |  |  |  |  |  |  |
| Free | 54 | 78.2 (71.9, 83.5) | 31% (21, 40) | 92.6 (90.3, 94.4) | 20% (11, 28) | 0.94 (0.92, 0.96) | 2% | Ref. | - |
| Immune complex | 6 | 81.4 (69.6, 89.4) | 14% (0, 32) | 94.3 (84.9, 98.0) | 31% (1, 60) | 0.94 (0.91, 0.95) | 4% | 0.81 |  |

**(1)** Where the sum of studies in the subgroups is less than the overall number of studies, one or more subgroups were excluded because they did not meet the minimum requirement of 4 studies for bivariate modelling.
**(2)** Ref. indicates the reference category (most studies) used for pairwise Wald tests. Global P values indicate the significance of the factor across all subgroups using a joint Wald test; pairwise P values compare the specific subgroup against the reference.

**(3)** Estimates not reported due to failure of model to converge.

**Abbreviations:** **AFRO:** WHO African Region; **AMRO:** WHO Region of the Americas; **AUC:** Area Under the Curve; **CI:** Confidence Interval; **ELISA:** Enzyme-Linked Immunosorbent Assay; **EMRO:** WHO Eastern Mediterranean Region; **SEARO:** WHO South-East Asian Region; **WPRO:** WHO Western Pacific Region.

### Table S7: Meta-regression of Lipid/Glycolipid Studies

| **Factor ^1^** | **N** | **Sensitivity**  **(95% CI)** | **Variation in sensitivity attributable to between-study heterogeneity,**  ***I^2^* (95% CI)** | **Specificity (95% CI)** | **Variation in specificity attributable to between-study heterogeneity,**  ***I^2^* (95% CI)** | **AUC**  **(95% CI)** | **Proportion of between-study heterogeneity due to threshold effect** | **Pairwise**  **P value** | **Global P value** |
| --- | --- | --- | --- | --- | --- | --- | --- | --- | --- |
| Overall | 22 | 68.6 (54.1, 80.3) | 37% (21, 54) | 97.0 (94.0, 98.5) | 32% (09, 54) | 0.96 (0.94, 0.97) | 8% | - | - |
| Decade of publication |  |  |  |  |  |  |  |  |  |
| 1990s | 4 | 66.1 (44.3, 82.7) | 18% (0, 44) | 98.7 (73.0, 100.0) | 59% (0, 100) | 0.90 (0.87, 0.92) | 46% | 0.39 | 0.33 |
| 2010s | 10 | 71.9 (47.4, 87.9) | 43% (15, 71) | 97.8 (92.3, 99.4) | 37% (3, 72) | 0.97 (0.95, 0.98) | 23% | 0.12 |  |
| 2020s | 7 | 56.0 (40.5, 70.4) | 16% (0, 32) | 94.1 (88.0, 97.2) | 17% (0, 40) | 0.89 (0.86, 0.91) | 100% | Ref. **^2^** |  |
| WHO Region |  |  |  |  |  |  |  |  |  |
| AFRO | 8 | 59.1 (43.0, 73.5) | 19% (0, 37) | 96.8 (92.6, 98.7) | 15% (0, 42) | 0.94 (0.92, 0.96) | 1% | Ref. | - |
| EURO | 5 | 70.5 (47.2, 86.4) | 23% (0, 50) | 89.5 (81.4, 94.3) | 3% (0, 17) | 0.91 (0.88, 0.93) | 7% | 0.096 |  |
| Study Design |  |  |  |  |  |  |  |  |  |
| Multi-gate | 18 | 72.5 (56.2, 84.5) | 40% (21, 59) | 97.9 (94.2, 99.2) | 42% (13, 71) | 0.97 (0.95, 0.98) | 14% | - | - |
| Single-gate | 4 **^3^** |  |  |  |  |  |  |  |  |
| Detection Method |  |  |  |  |  |  |  |  |  |
| ELISA | 5 | 73.6 (31.2, 94.5) | 54% (12, 96) | 94.5 (89.1, 97.3) | 4% (0, 23) | 0.95 (0.93, 0.97) | 6% | Ref. | 0.25 |
| ECLIA | 6 | 47.4 (33.5, 61.7) | 13% (0, 26) | 96.1 (92.2, 98.1) | 8% (0, 26) | 0.92 (0.89, 0.94) | 100% | 0.43 |  |
| Mass spectrometry | 4 | 74.2 (53.9, 87.6) | 11% (0, 52) | 87.6 (53.9, 87.6) | 5% (0, 29) | 0.90 (0.87, 0.92) | 100% | 0.31 |  |
| Sample Type |  |  |  |  |  |  |  |  |  |
| Serum | 16 | 70.0 (51.9, 83.4) | 41% (22, 61) | 98.1 (95.0, 99.3) | 38% (9, 66) | 0.97 (0.95, 0.98) | 14% | Ref. | - |
| Plasma | 4 | 69.9 (44.5, 87.0) | 22% (0, 51) | 92.6 (87.0, 95.9) | 0% (0, 4) | 0.93 (0.91, 0.95) | 100% | 0.38 |  |

**(1**) Where the sum of studies in the subgroups is less than the overall number of studies, one or more subgroups were excluded because they did not meet the minimum requirement of 4 studies for bivariate modelling.
**(2**) Ref. indicates the reference category (most studies) used for pairwise Wald tests. Global P values indicate the significance of the factor across all subgroups using a joint Wald test; pairwise P values compare the specific subgroup against the reference.

**(3)** Estimates not reported due to failure of model to converge.

**Abbreviations:** **AFRO:** WHO African Region; **AUC:** Area Under the Curve; **CI:** Confidence Interval; ECLIA, Electrochemiluminescence immunoassay; **ELISA:**Enzyme-Linked Immunosorbent Assay; **EURO:** WHO European Region.

### Figure S1: Forest plot showing prevalence of cell-free Mtb DNA detection among all individuals with active tuberculosis

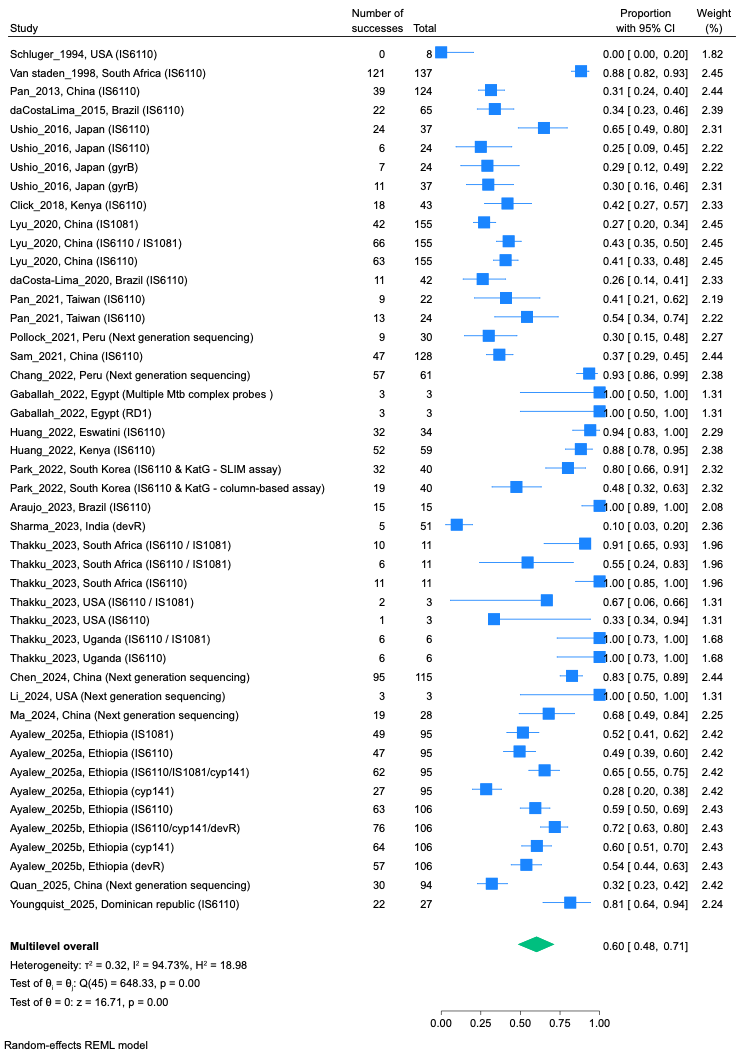

### Figure S2: Forest plot showing prevalence of cell-free Mtb DNA detection among HIV-uninfected individuals with active tuberculosis

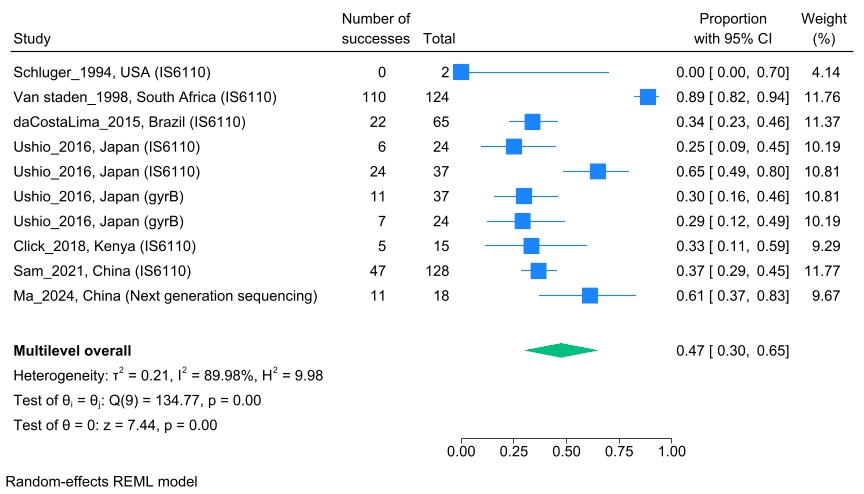

### Figure S3: Forest plot showing prevalence of cell-free Mtb DNA detection among HIV-infected individuals with active tuberculosis

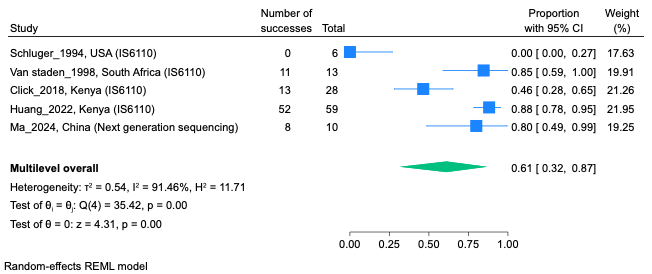

### Figure S4: Forest plot showing prevalence of cell-free Mtb DNA detection among individuals with active tuberculosis with unknown HIV status

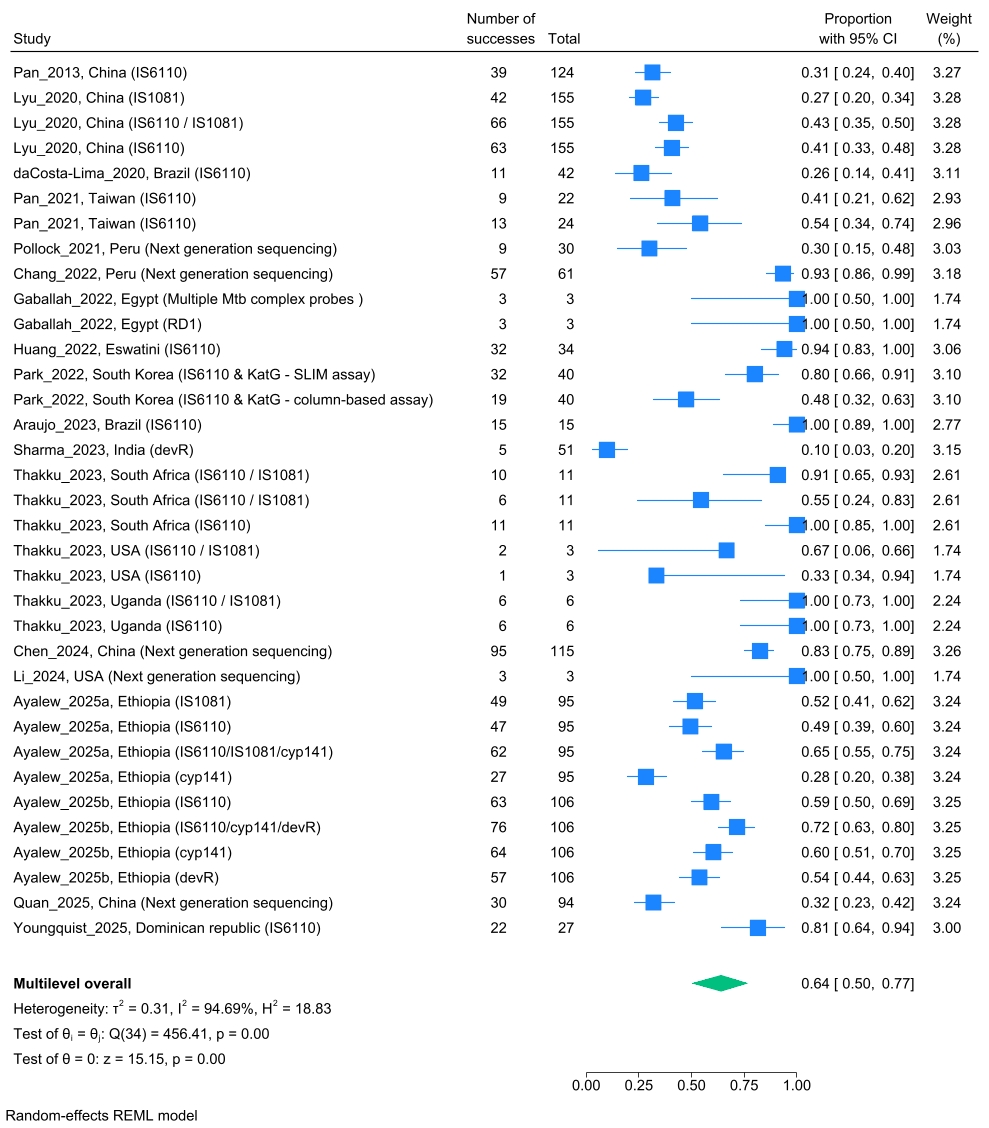

### Figure S5: Forest plot showing prevalence of cell-free Mtb DNA detection among all individuals without active tuberculosis

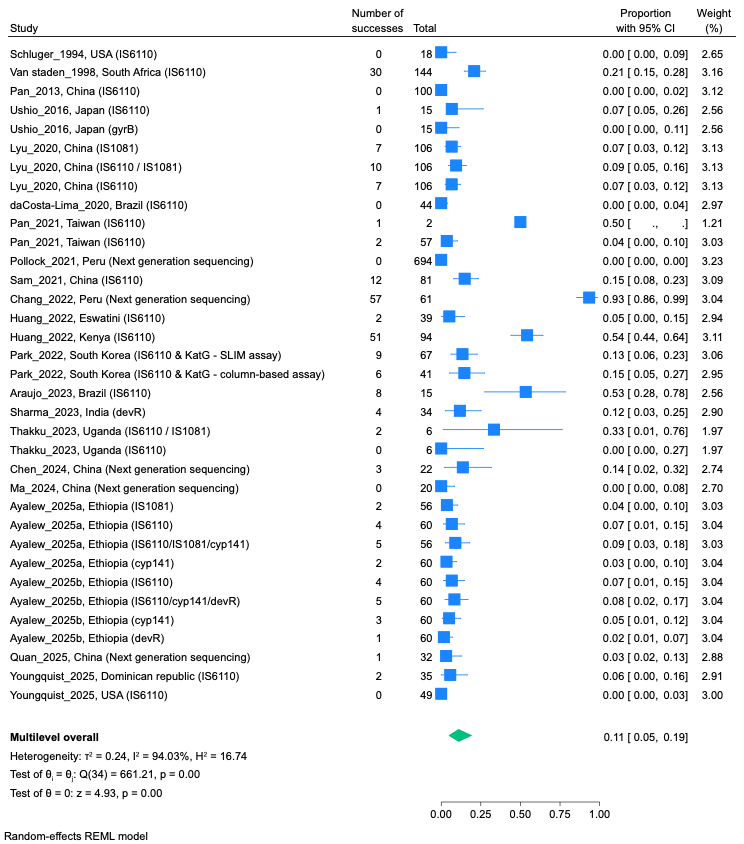

### Figure S6: Forest plot showing prevalence of cell-free Mtb DNA detection among individuals with non-tuberculosis illness

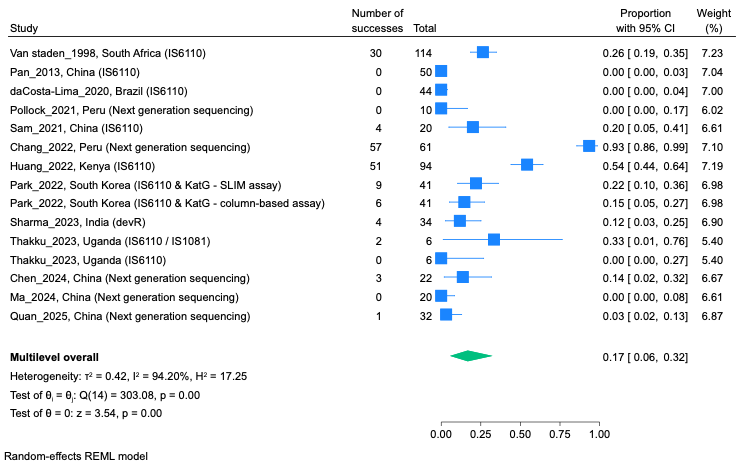

### Figure S7: Forest plot showing prevalence of cell-free Mtb DNA detection among asymptomatic IGRA/TST-positive individuals

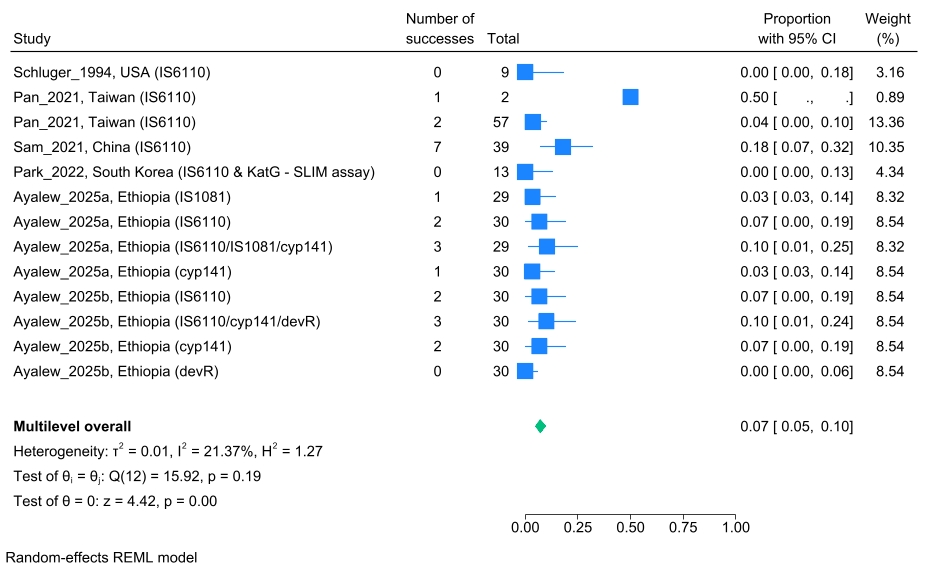

### Figure S8: Forest plot showing prevalence of cell-free Mtb DNA detection among asymptomatic IGRA/TST/TST-negative individuals

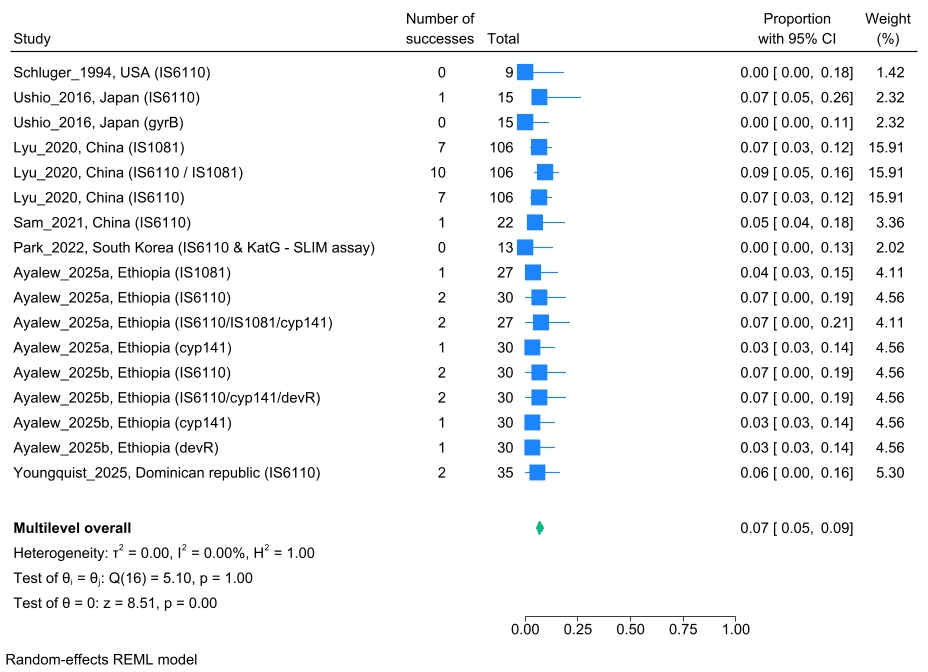

### Figure S9: Forest plot showing prevalence of cell-free Mtb DNA detection among asymptomatic individuals with unknown IGRA/TST/TST status

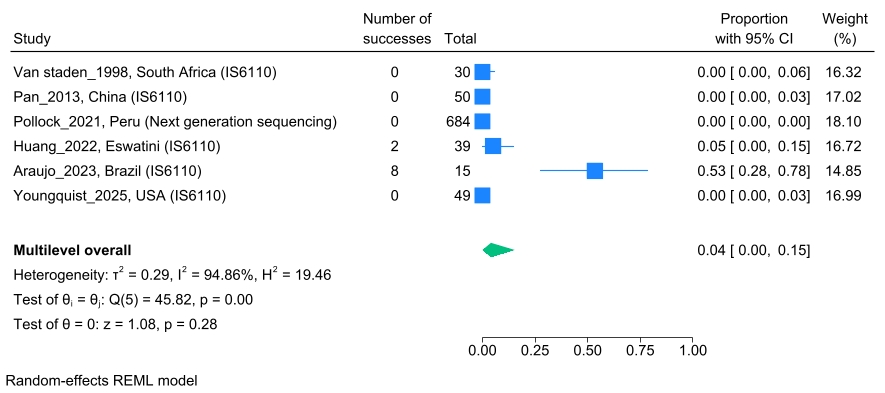

### Figure S10: Forest plot showing prevalence of cell-associated Mtb DNA detection among all individuals with active tuberculosis

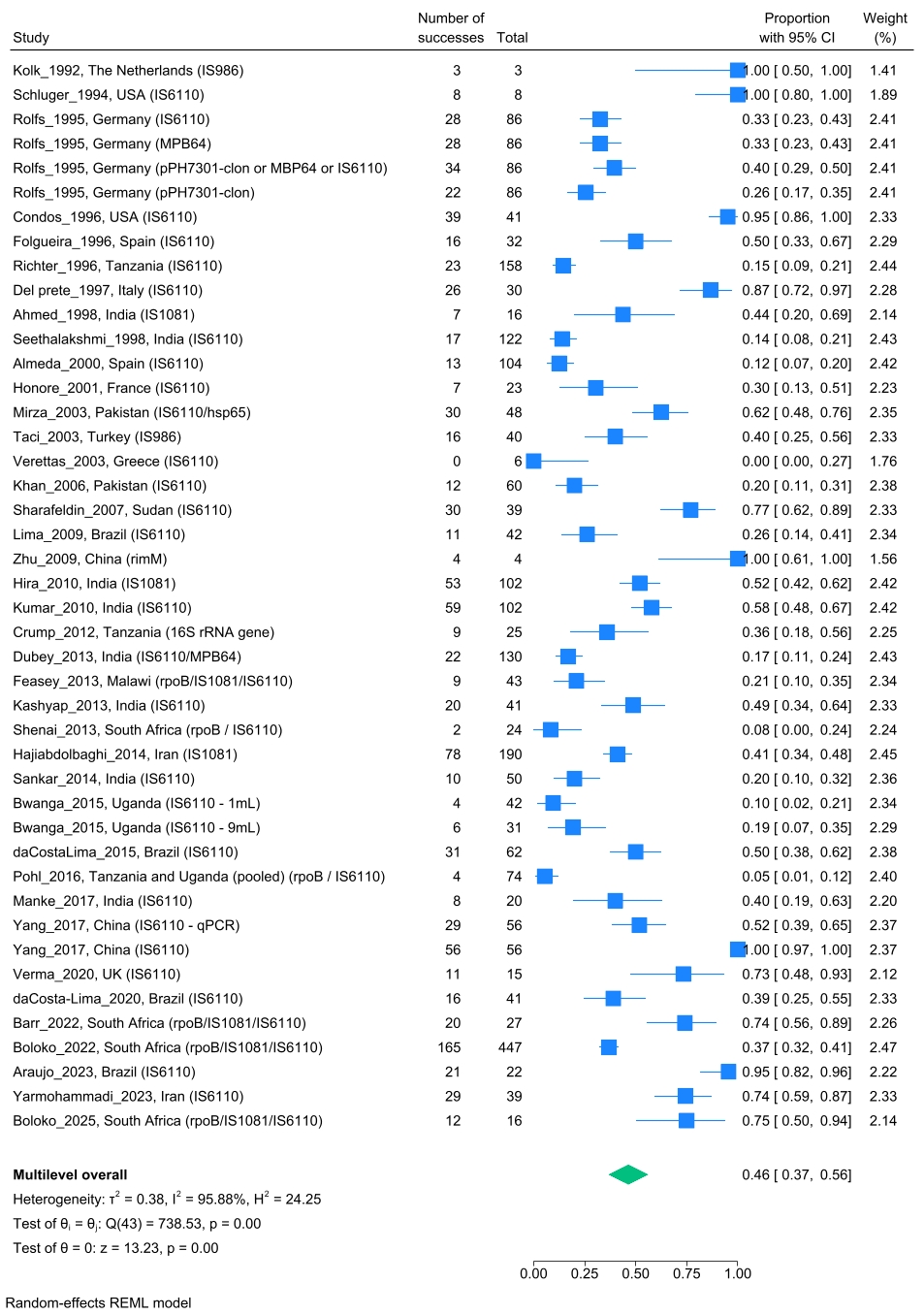

### Figure S11: Forest plot showing prevalence of cell-associated Mtb DNA detection among HIV-uninfected individuals with active tuberculosis

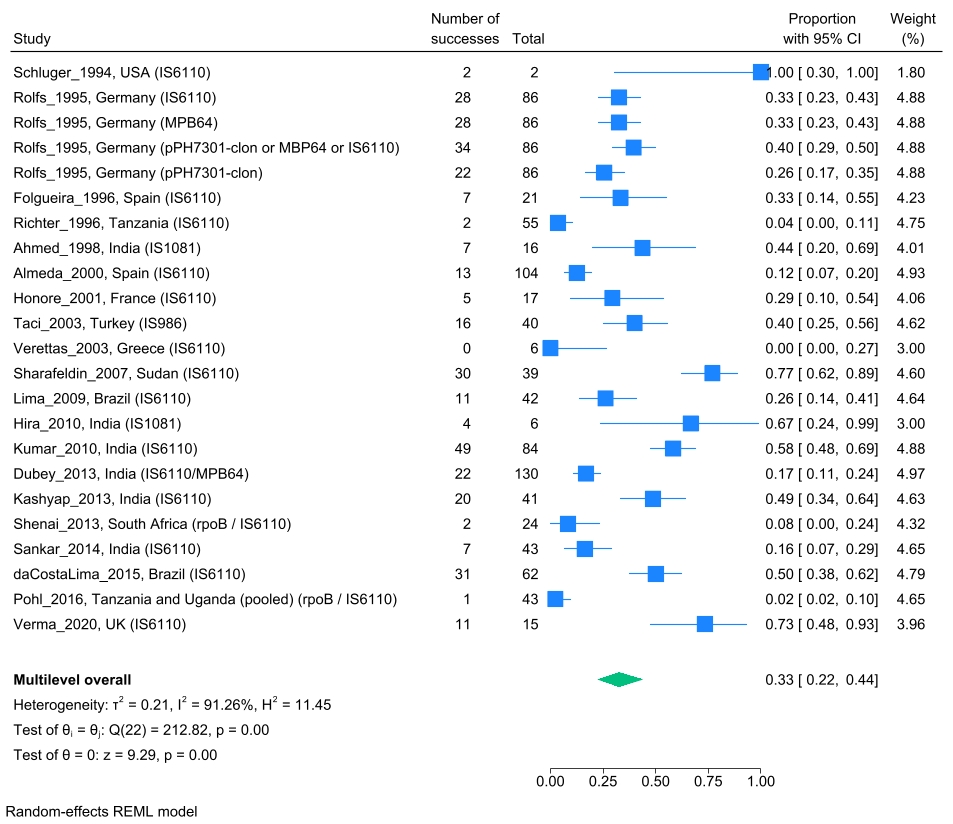

### Figure S12: Forest plot showing prevalence of cell-associated Mtb DNA detection among HIV-infected individuals with active tuberculosis

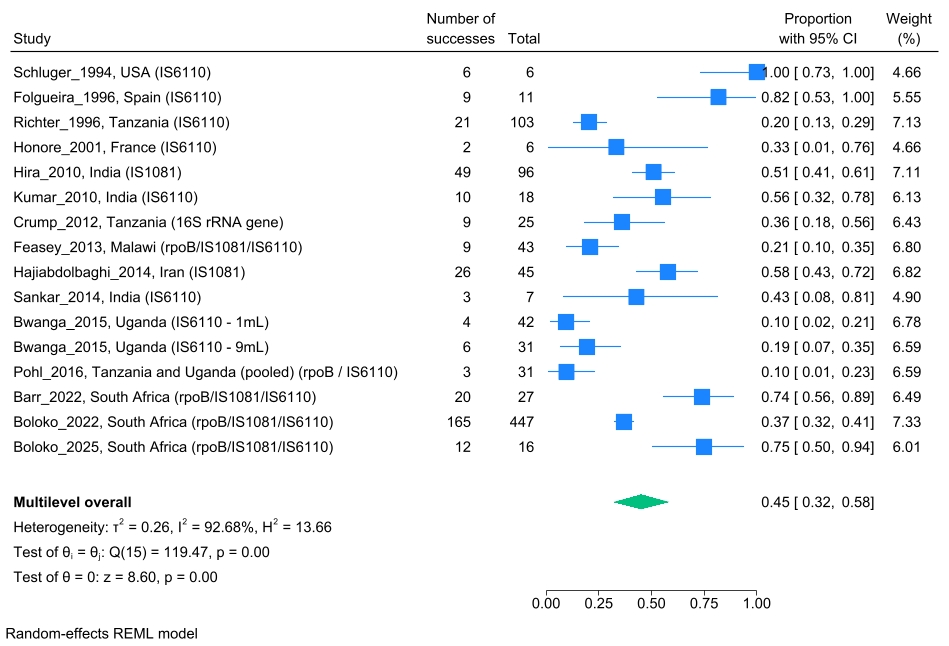

### Figure S13: Forest plot showing prevalence of cell-associated Mtb DNA detection among individuals with active tuberculosis and HIV status unknown

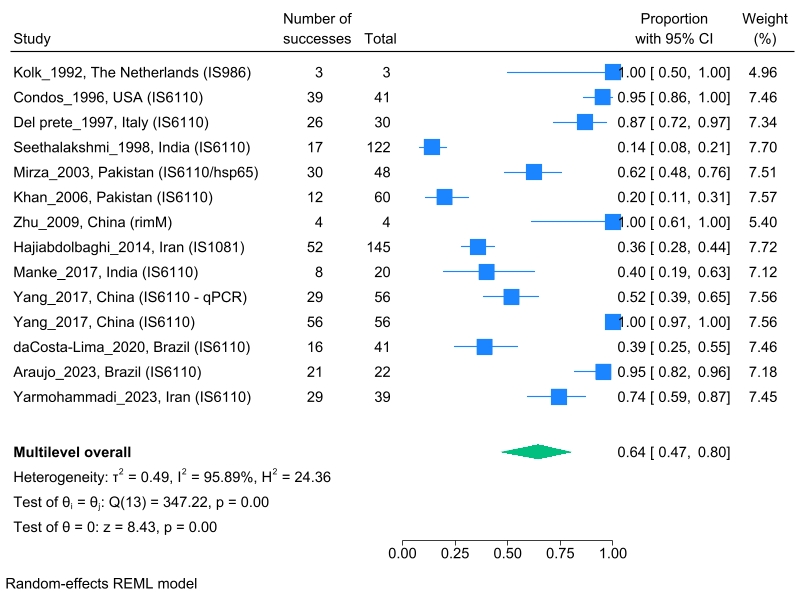

### Figure S14: Forest plot showing prevalence of cell-associated Mtb DNA detection among all individuals without active tuberculosis

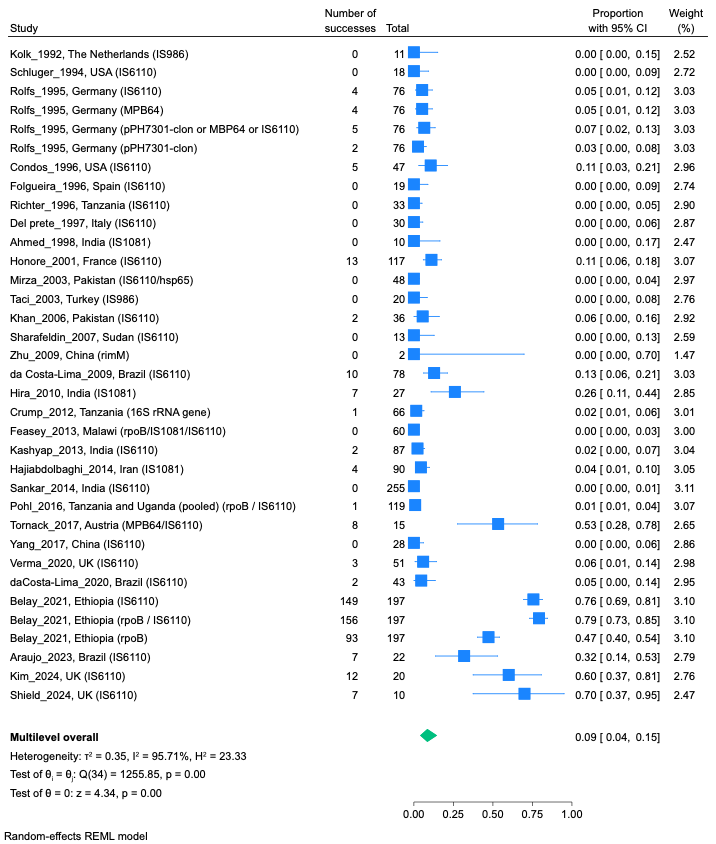

### Figure S15: Forest plot showing prevalence of cell-associated Mtb DNA detection among individuals with non-tuberculosis illness

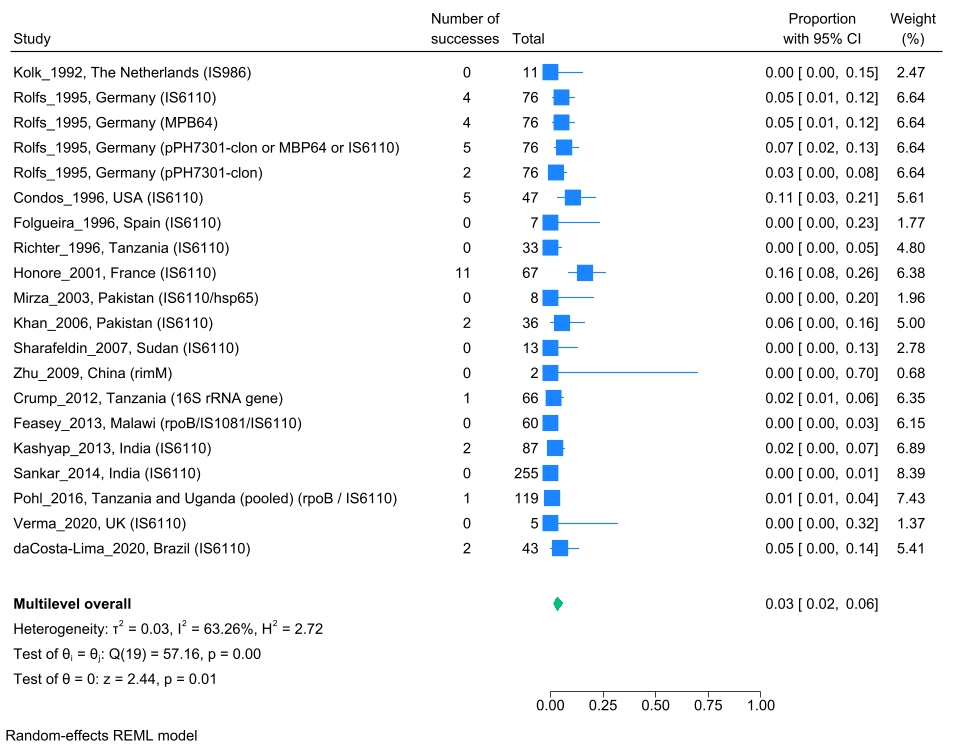

### Figure S16: Forest plot showing prevalence of cell-associated Mtb DNA detection among asymptomatic IGRA/TST/TST-positive individuals

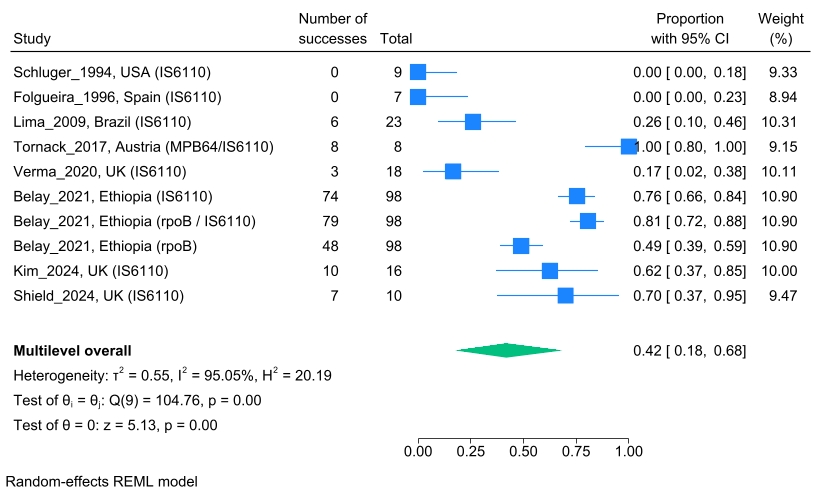

### Figure S17: Forest plot showing prevalence of cell-associated Mtb DNA detection among asymptomatic IGRA/TST/TST-negative individuals

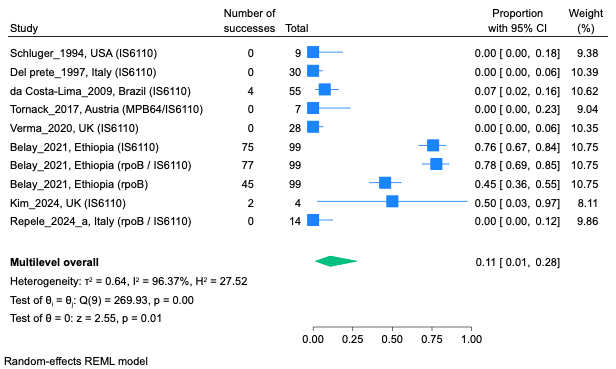

### Figure S18: Forest plot showing prevalence of cell-associated Mtb DNA detection among asymptomatic individuals with unknown IGRA/TST/TST status

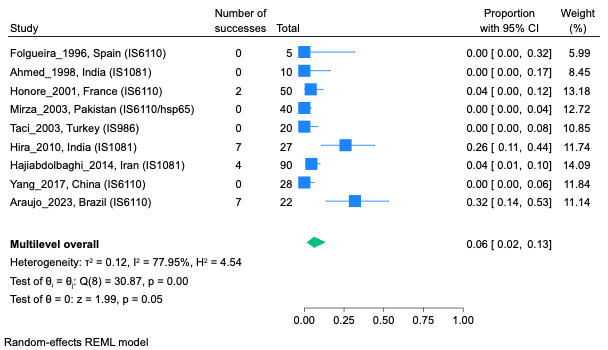

### Figure S19: Forest plot showing prevalence of protein/peptide antigen detection among all individuals with active tuberculosis

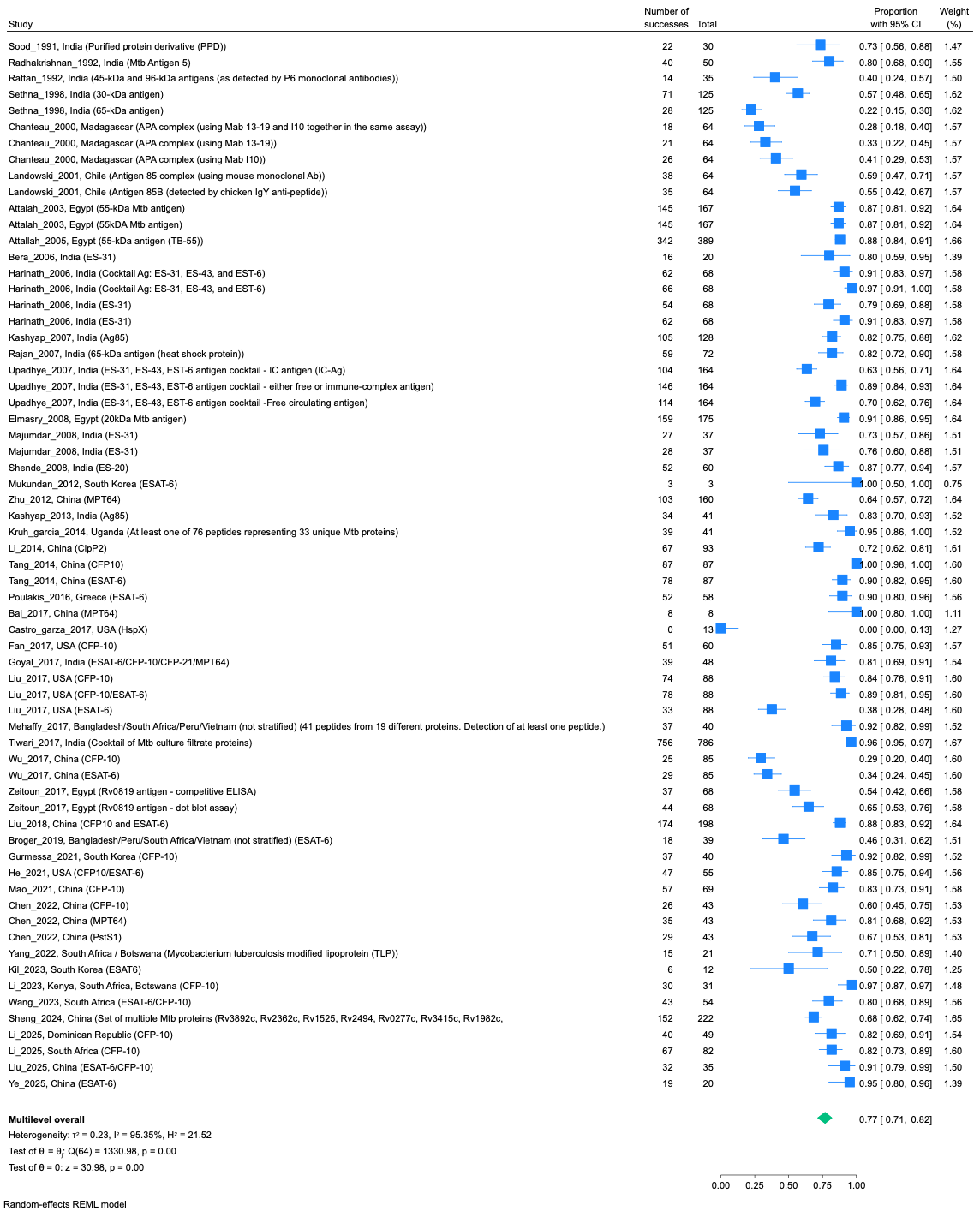

### Figure S20: Forest plot showing prevalence of protein/peptide antigen detection among HIV-uninfected individuals with active tuberculosis

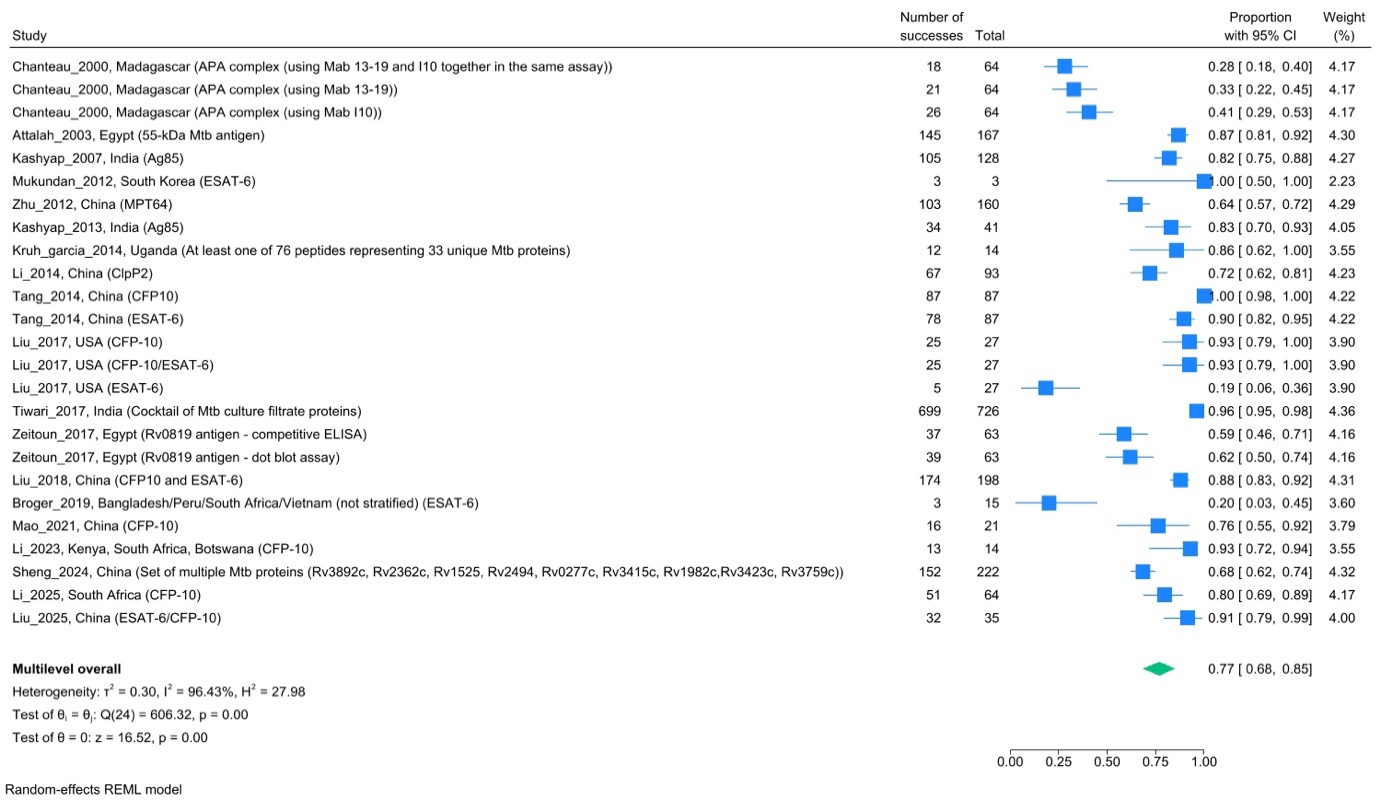

### Figure S21: Forest plot showing prevalence of protein/peptide antigen detection among HIV-infected individuals with active tuberculosis

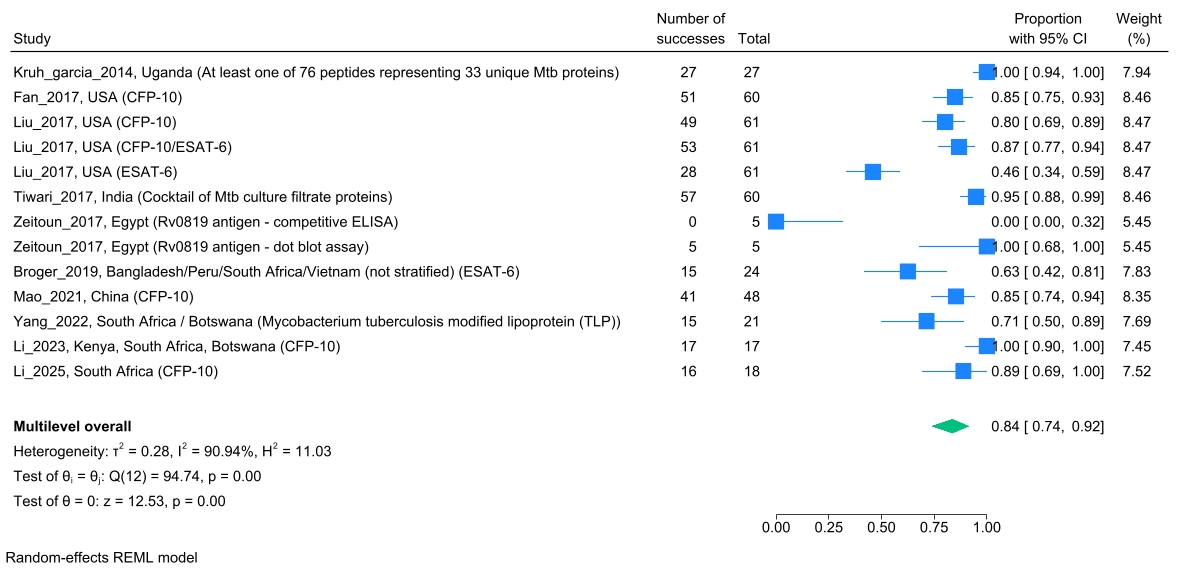

### Figure S22: Forest plot showing prevalence of protein/peptide antigen detection among individuals with active tuberculosis and unknown HIV status

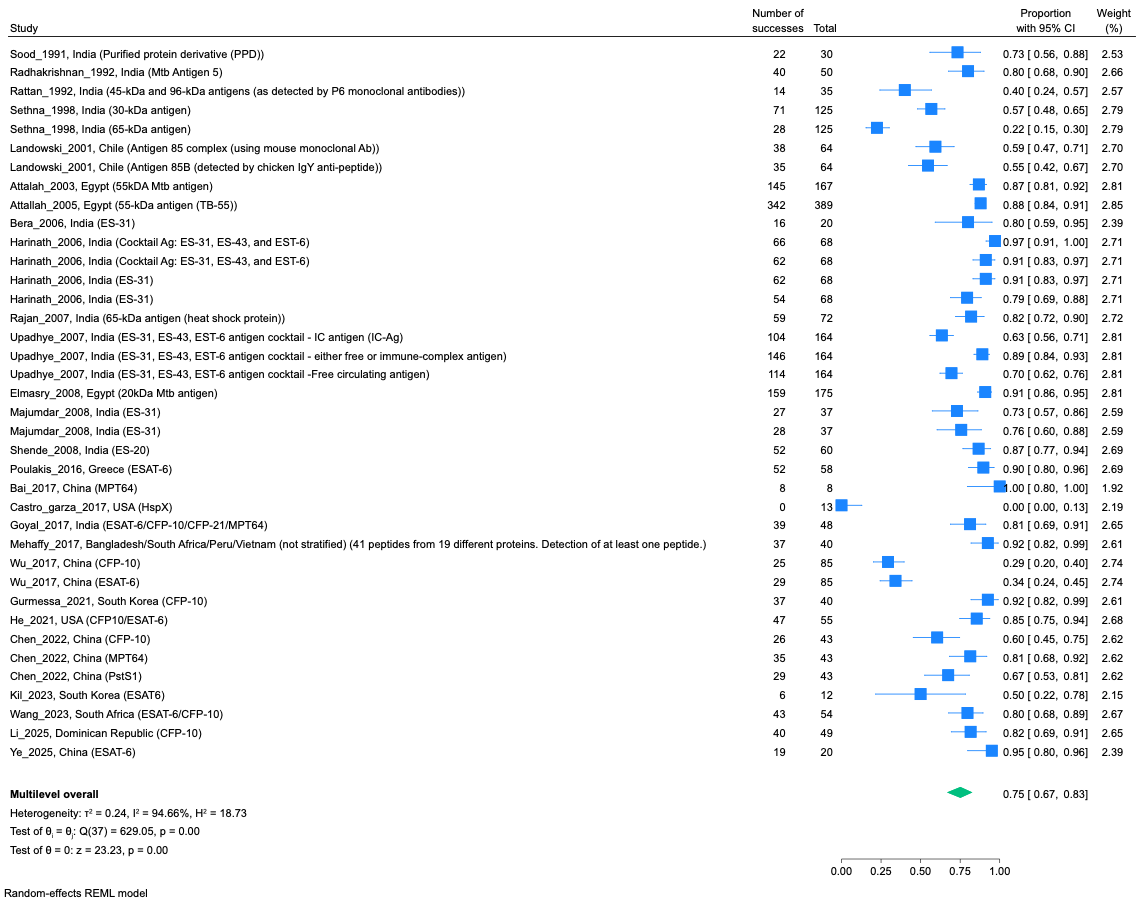

### Figure S23: Forest plot showing prevalence of protein/peptide antigen detection among all individuals without active tuberculosis

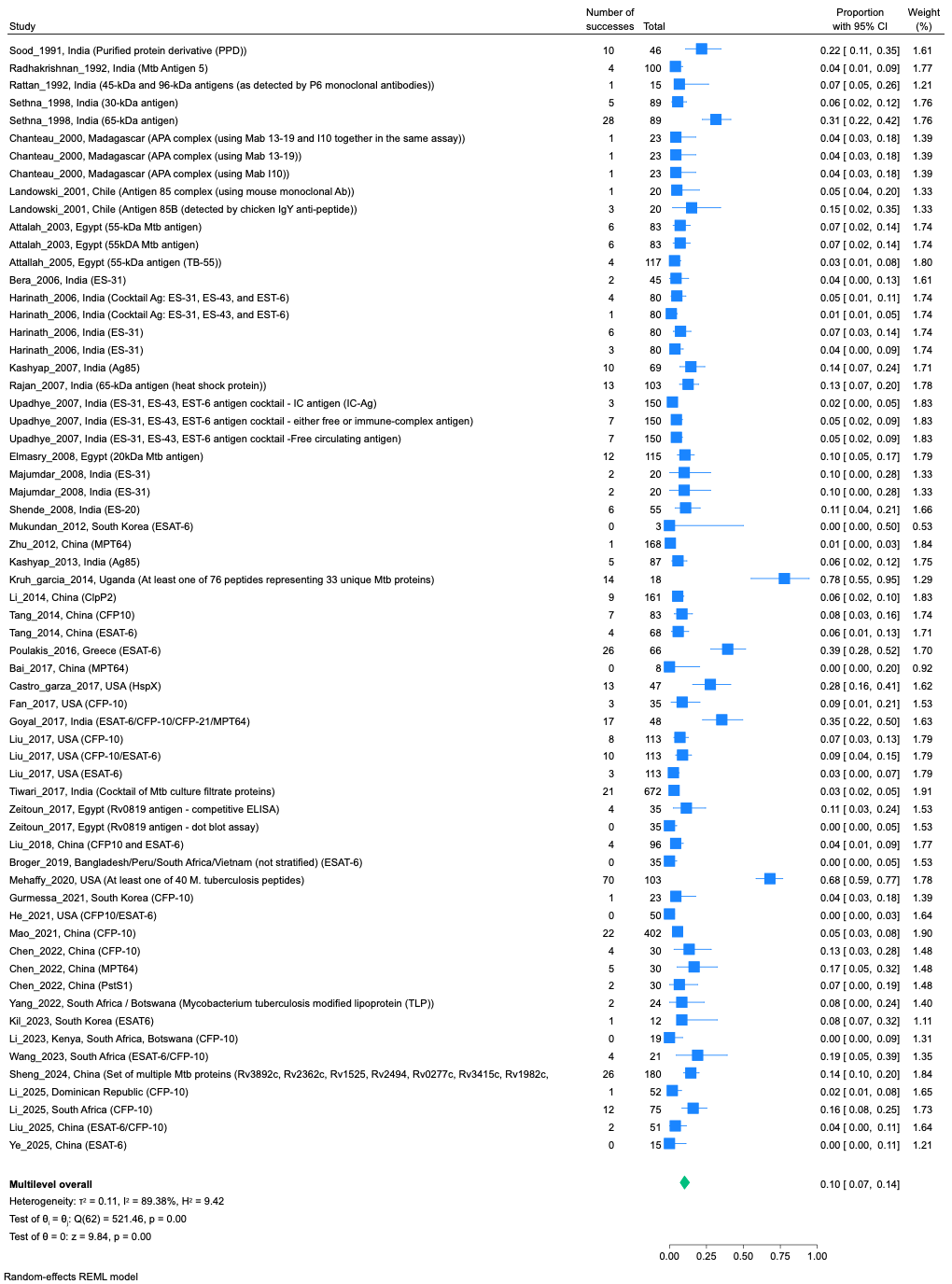

### Figure S24: Forest plot showing prevalence of protein/peptide antigen detection among individuals with non-tuberculosis illness

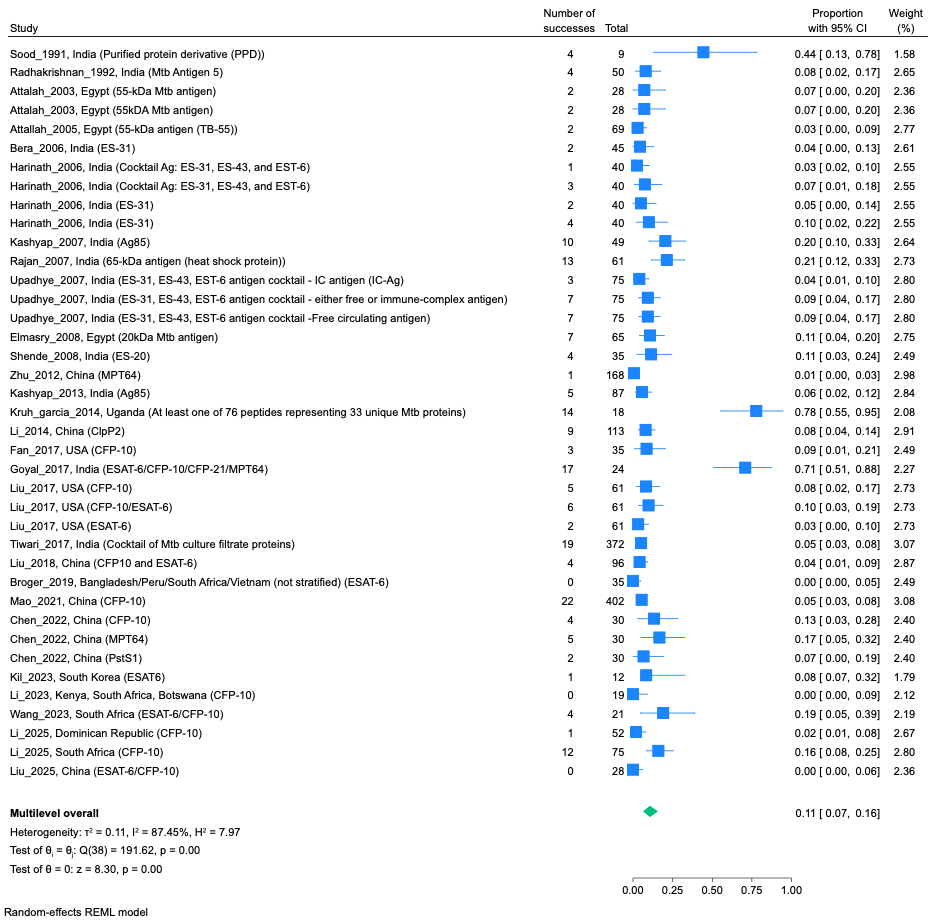

### Figure S25: Forest plot showing prevalence of protein/peptide antigen detection among asymptomatic IGRA/TST-positive individuals

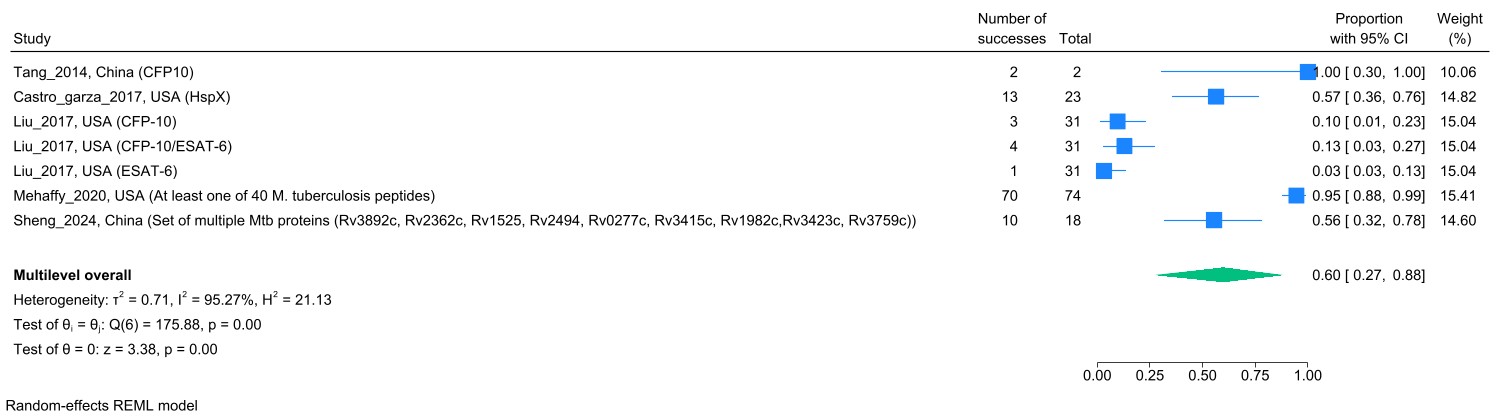

### Figure S26: Forest plot showing prevalence of protein/peptide antigen detection among asymptomatic IGRA/TST-negative individuals

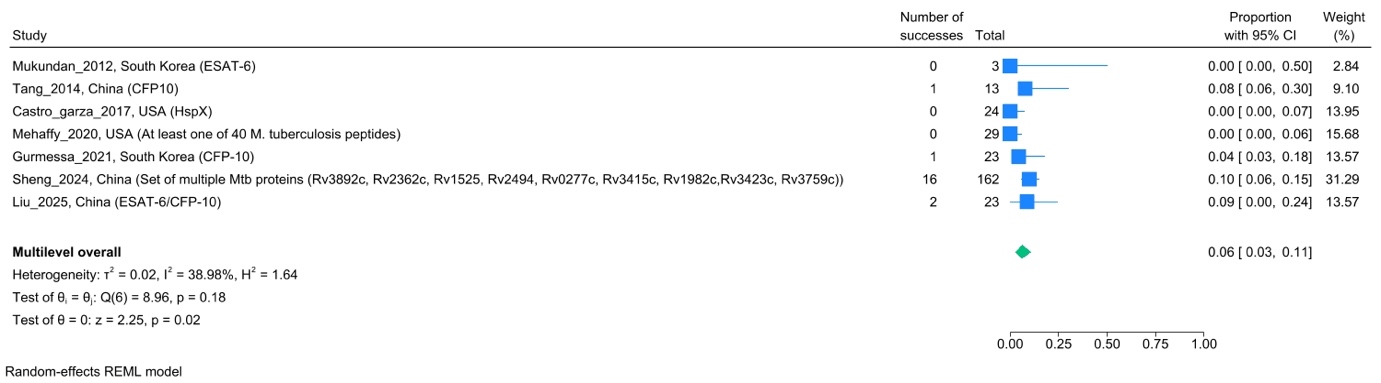

### Figure S27: Forest plot showing prevalence of protein/peptide antigen detection among asymptomatic individuals with unknown IGRA/TST status

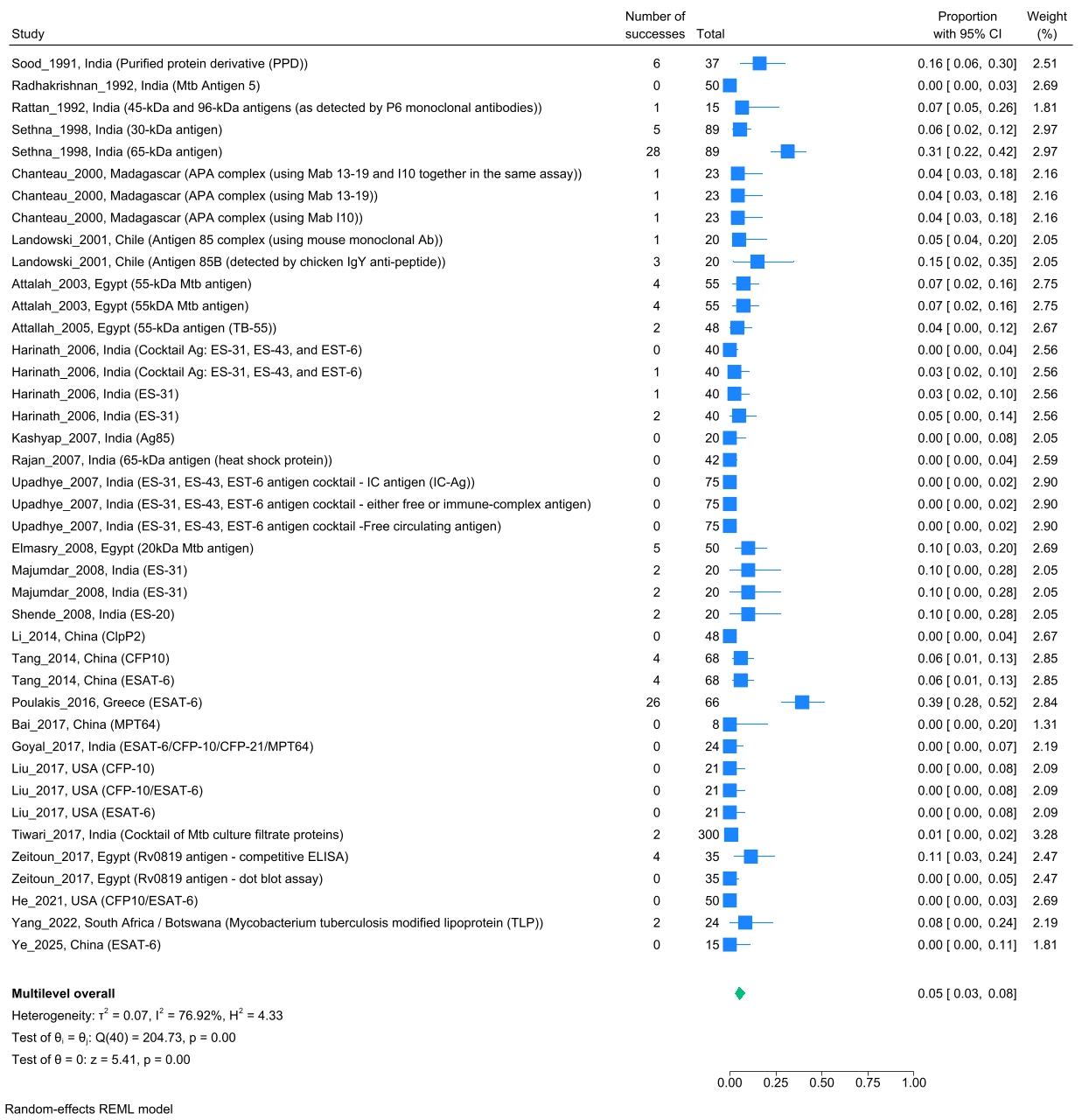

### Figure S28: Forest plot showing prevalence of lipid/glycolipid antigen detection among all individuals with active tuberculosis

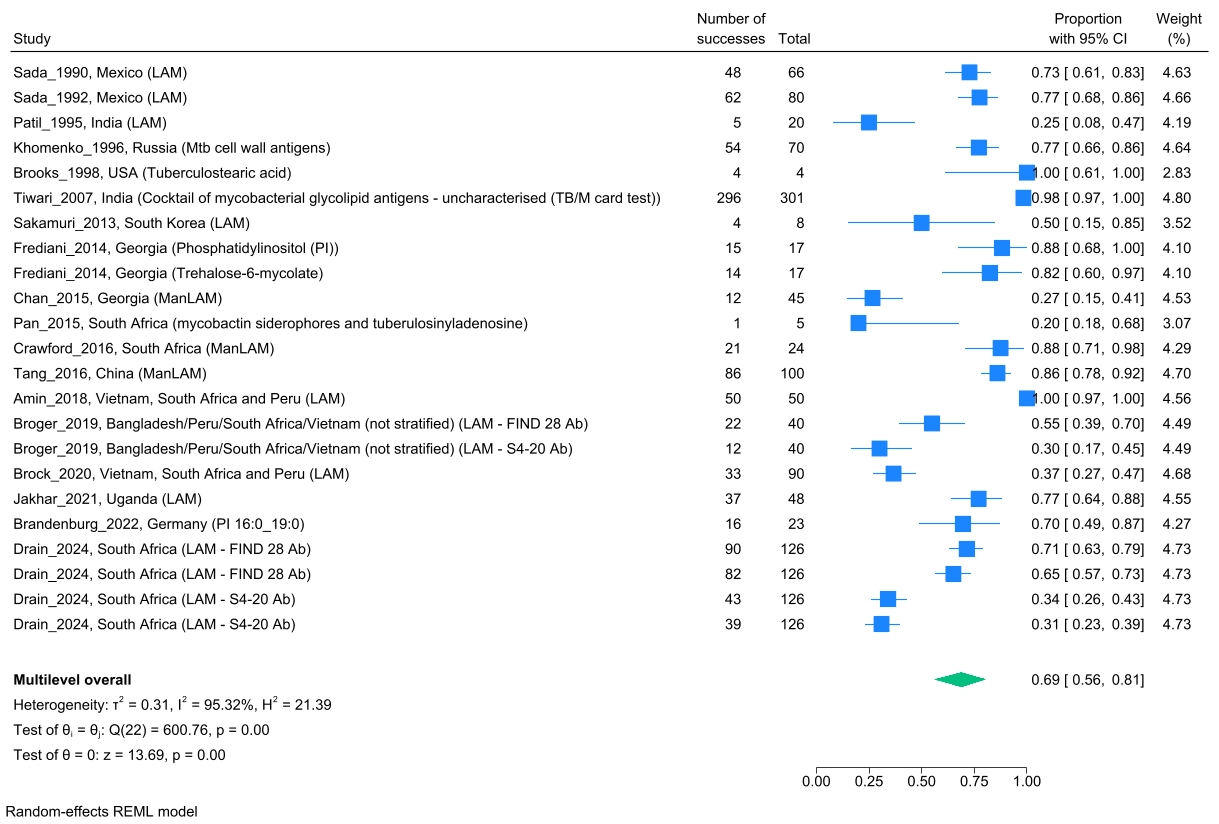

### Figure S29: Forest plot showing prevalence of lipid/glycolipid antigen detection among HIV-uninfected individuals with active tuberculosis

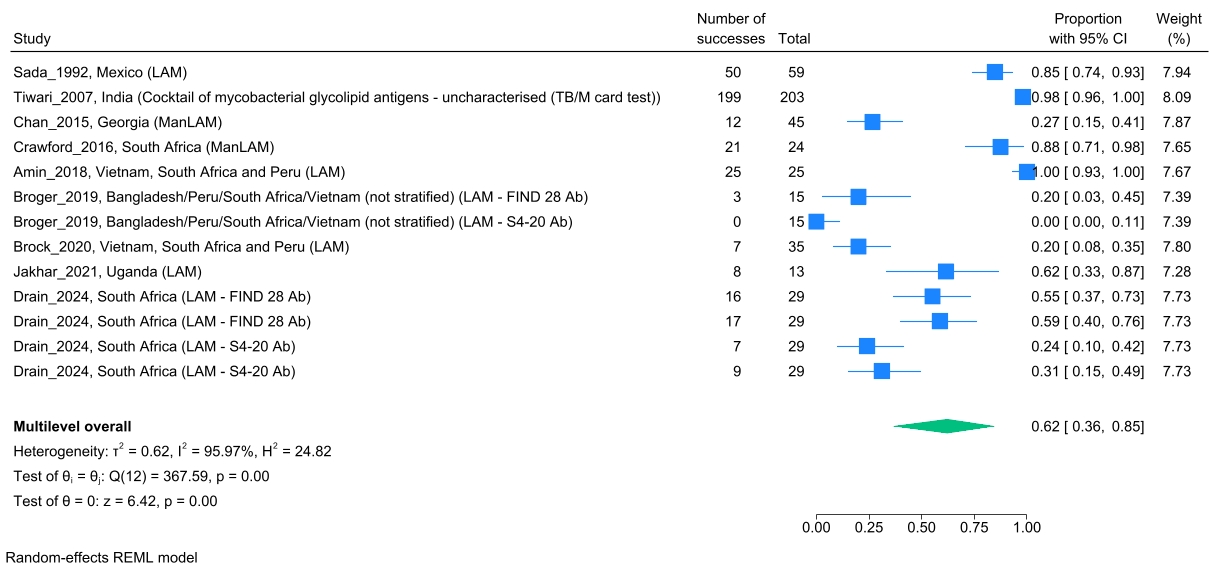

### Figure S30: Forest plot showing prevalence of lipid/glycolipid antigen detection among HIV-infected individuals with active tuberculosis

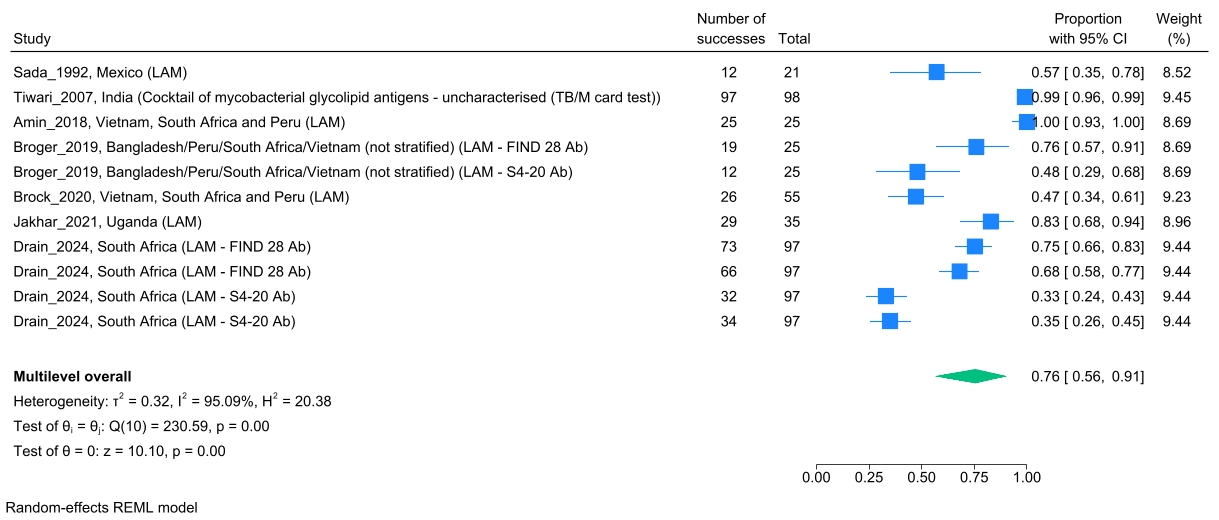

### Figure S31: Forest plot showing prevalence of lipid/glycolipid antigen detection among individuals with active tuberculosis and unknown HIV status

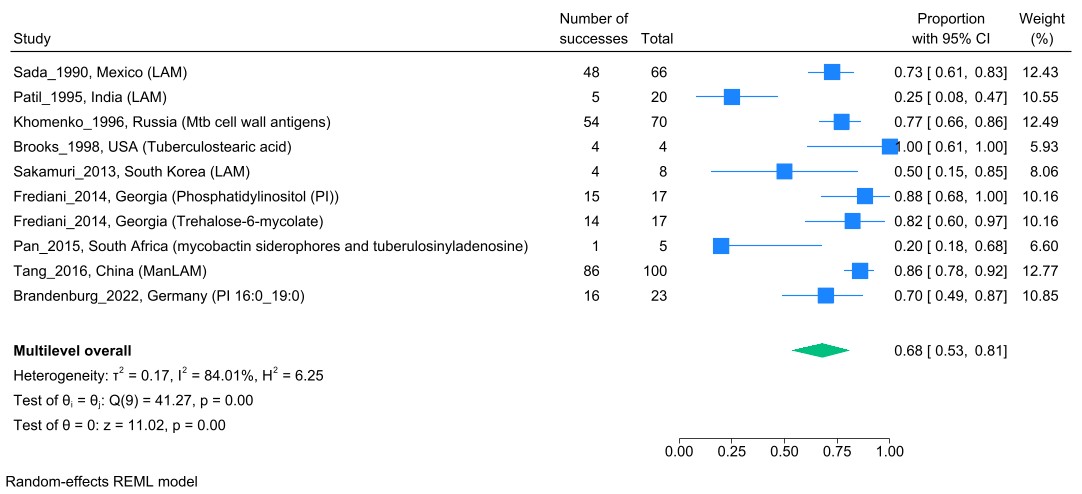

### Figure S32: Forest plot showing prevalence of lipid/glycolipid antigen detection among all individuals without active tuberculosis

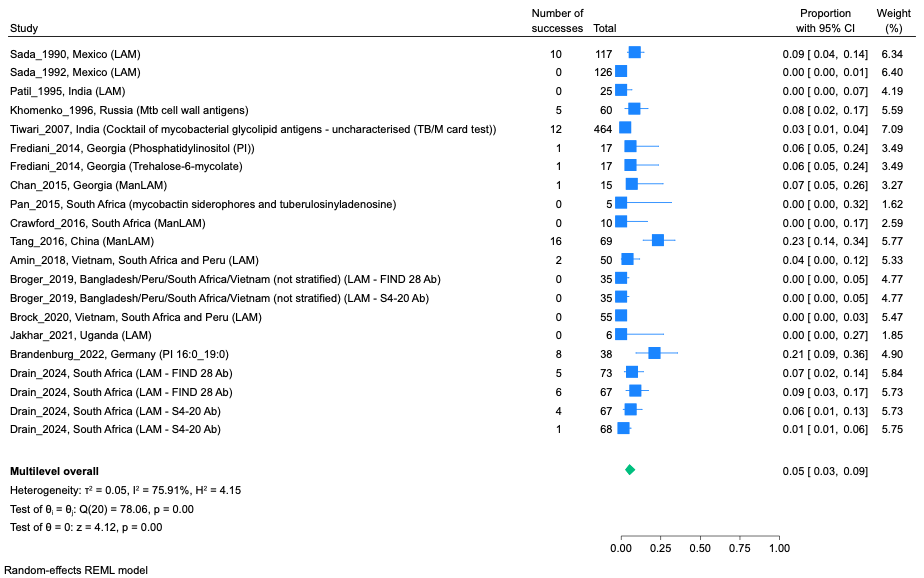

### Figure S33: Forest plot showing prevalence of lipid/glycolipid antigen detection among individuals with non-tuberculosis illness

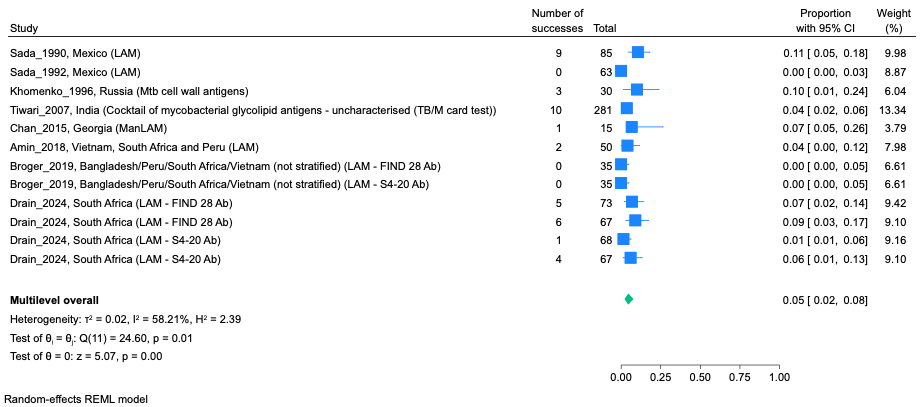

### Figure S34: Forest plot showing prevalence of lipid/glycolipid antigen detection among asymptomatic individuals with unknown IGRA/TST status

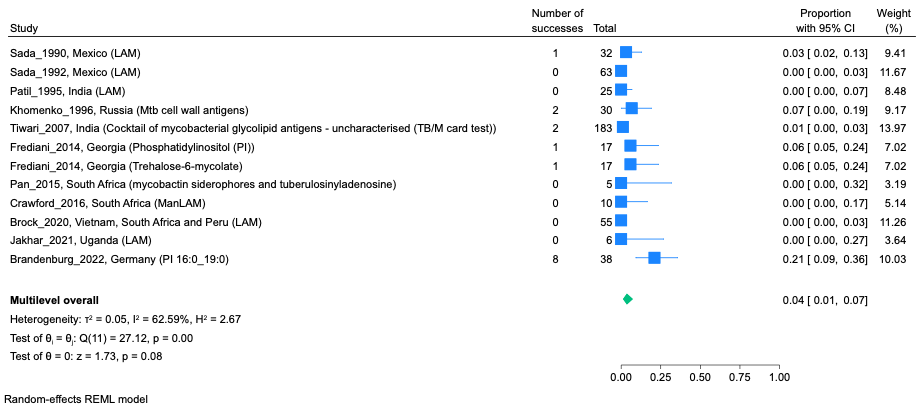

### Figure S35: Forest plot showing prevalence of unspecified/combined antigen detection among all individuals with active tuberculosis

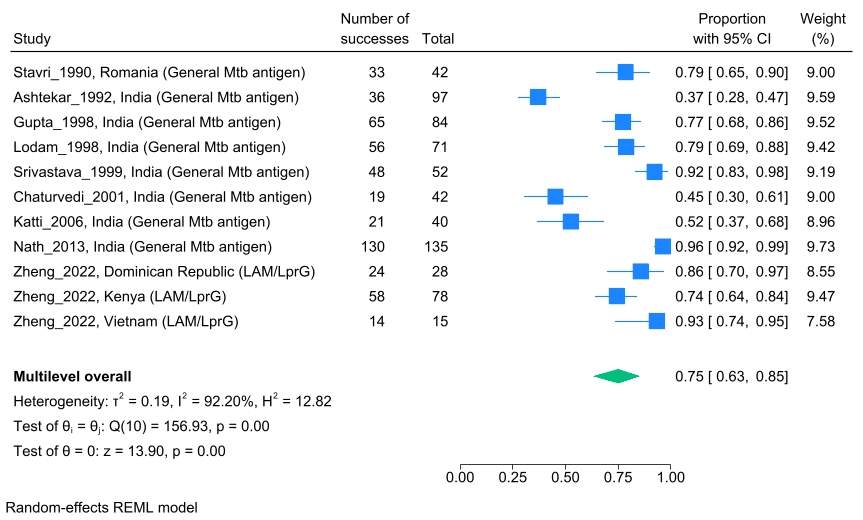

### Figure S36: Forest plot showing prevalence of unspecified/combined antigen detection among individuals with active tuberculosis and unknown HIV status

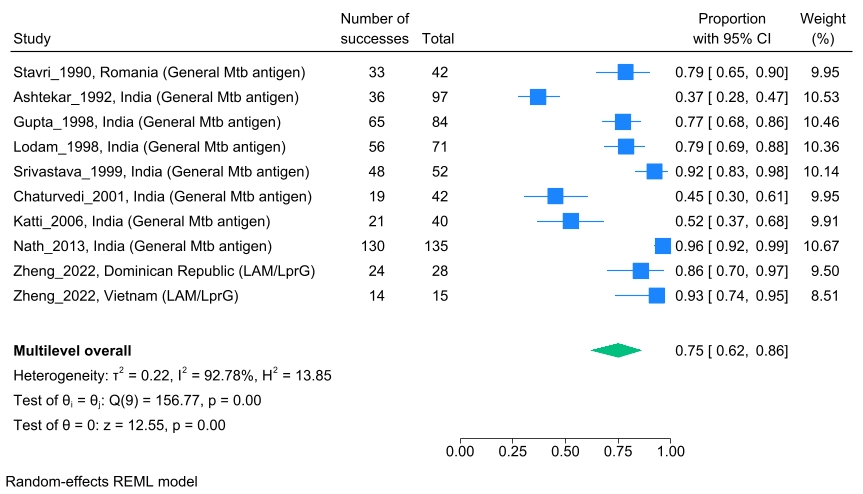

### Figure S37: Forest plot showing prevalence of unspecified/combined antigen detection among all individuals without active tuberculosis

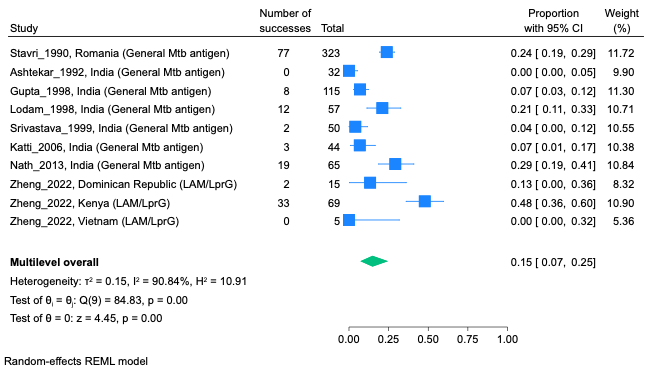

### Figure S38: Forest plot showing prevalence of unspecified/combined antigen detection among individuals with non-tuberculosis illness

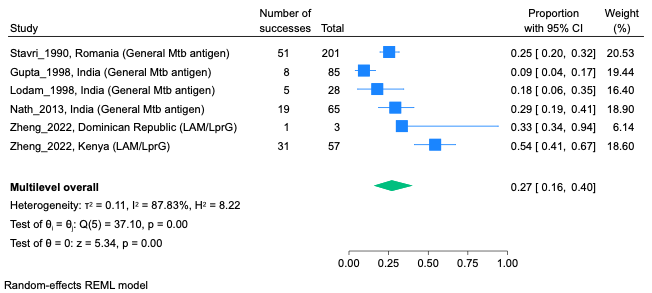

### Figure S39: Forest plot showing prevalence of unspecified/combined antigen detection among asymptomatic IGRA/TST-negative individuals

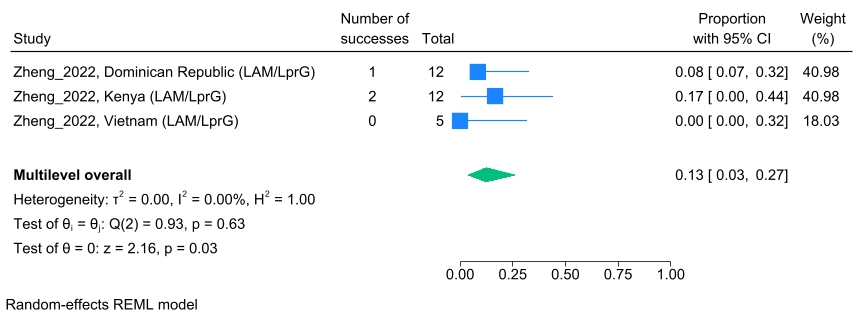

### Figure S40: Forest plot showing prevalence of unspecified/combined antigen detection among asymptomatic individuals with unknown IGRA/TST status

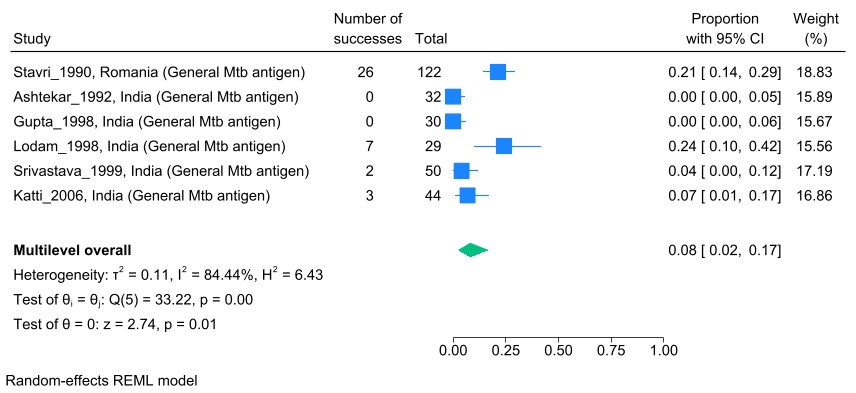

### Figure S41: Deeks’ Funnel plot of bivariate analysis studies

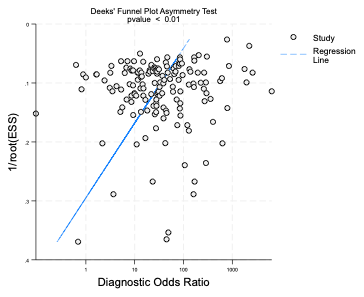

### Figure S42: Bubble plot of correlation between longitudinal study risk difference and follow-up sampling timepoint.

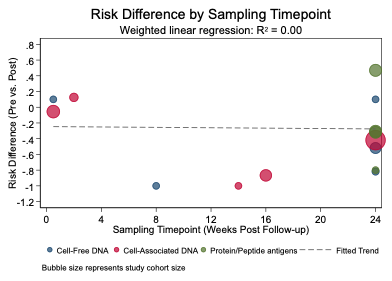
